# Large-scale analysis of structural brain asymmetries in schizophrenia via the ENIGMA consortium

**DOI:** 10.1101/2022.03.01.22271652

**Authors:** Dick Schijven, Merel C. Postema, Masaki Fukunaga, Junya Matsumoto, Kenichiro Miura, Sonja M.C. de Zwarte, Neeltje E.M. van Haren, Wiepke Cahn, Hilleke E. Hulshoff Pol, René S. Kahn, Rosa Ayesa-Arriola, Víctor Ortiz-García de la Foz, Diana Tordesillas-Gutierrez, Javier Vázquez-Bourgon, Benedicto Crespo-Facorro, Dag Alnæs, Andreas Dahl, Lars T. Westlye, Ingrid Agartz, Ole A. Andreassen, Erik G. Jönsson, Peter Kochunov, Jason M. Bruggemann, Stanley V. Catts, Patricia T. Michie, Bryan J. Mowry, Yann Quidé, Paul E. Rasser, Ulrich Schall, Rodney J. Scott, Vaughan J. Carr, Melissa J. Green, Frans A. Henskens, Carmel M. Loughland, Christos Pantelis, Cynthia Shannon Weickert, Thomas W. Weickert, Lieuwe de Haan, Katharina Brosch, Julia-Katharina Pfarr, Kai G. Ringwald, Frederike Stein, Andreas Jansen, Tilo T.J. Kircher, Igor Nenadic, Bernd Krämer, Oliver Gruber, Theodore D. Satterthwaite, Juan Bustillo, Daniel H. Mathalon, Adrian Preda, Vince D. Calhoun, Judith M. Ford, Steven G. Potkin, Jingxu Chen, Yunlong Tan, Zhiren Wang, Hong Xiang, Fengmei Fan, Fabio Bernardoni, Stefan Ehrlich, Paola Fuentes-Claramonte, Maria Angeles Garcia-Leon, Amalia Guerrero-Pedraza, Raymond Salvador, Salvador Sarró, Edith Pomarol-Clotet, Valentina Ciullo, Fabrizio Piras, Daniela Vecchio, Nerisa Banaj, Gianfranco Spalletta, Stijn Michielse, Therese van Amelsvoort, Erin W. Dickie, Aristotle N. Voineskos, Kang Sim, Simone Ciufolini, Paola Dazzan, Robin M. Murray, Woo-Sung Kim, Young-Chul Chung, Christina Andreou, André Schmidt, Stefan Borgwardt, Andrew M. McIntosh, Heather C. Whalley, Stephen M. Lawrie, Stefan du Plessis, Hilmar K. Luckhoff, Freda Scheffler, Robin Emsley, Dominik Grotegerd, Rebekka Lencer, Udo Dannlowski, Jesse T. Edmond, Kelly Rootes-Murdy, Julia M. Stephen, Andrew R. Mayer, Linda A. Antonucci, Leonardo Fazio, Giulio Pergola, Alessandro Bertolino, Covadonga M. Díaz-Caneja, Joost Janssen, Noemi G. Lois, Celso Arango, Alexander S. Tomyshev, Irina Lebedeva, Simon Cervenka, Carl M. Sellgren, Foivos Georgiadis, Matthias Kirschner, Stefan Kaiser, Tomas Hajek, Antonin Skoch, Filip Spaniel, Minah Kim, Yoo Bin Kwak, Sanghoon Oh, Jun Soo Kwon, Anthony James, Geor Bakker, Christian Knöchel, Michael Stäblein, Viola Oertel, Anne Uhlmann, Fleur M. Howells, Dan J. Stein, Henk S. Temmingh, Ana M. Diaz-Zuluaga, Julian A. Pineda-Zapata, Carlos López-Jaramillo, Stephanie Homan, Ellen Ji, Werner Surbeck, Philipp Homan, Simon E. Fisher, Barbara Franke, David C. Glahn, Ruben C. Gur, Ryota Hashimoto, Neda Jahanshad, Eileen Luders, Sarah E. Medland, Paul M. Thompson, Jessica A. Turner, Theo G.M. van Erp, Clyde Francks

## Abstract

Left-right asymmetry is an important organizing feature of the healthy brain that may be altered in schizophrenia, but most studies have used relatively small samples and heterogeneous approaches, resulting in equivocal findings. We carried out the largest case-control study of structural brain asymmetries in schizophrenia, using MRI data from 5,080 affected individuals and 6,015 controls across 46 datasets in the ENIGMA consortium, using a single image analysis protocol. Asymmetry indexes were calculated for global and regional cortical thickness, surface area, and subcortical volume measures. Differences of asymmetry were calculated between affected individuals and controls per dataset, and effect sizes were meta-analyzed across datasets. Small average case-control differences were observed for thickness asymmetries of the rostral anterior cingulate and the middle temporal gyrus, both driven by thinner left-hemispheric cortices in schizophrenia. Analyses of these asymmetries with respect to the use of antipsychotic medication and other clinical variables did not show any significant associations. Assessment of age- and sex-specific effects revealed a stronger average leftward asymmetry of pallidum volume between older cases and controls. Case-control differences in a multivariate context were assessed in a subset of the data (N = 2,029), which revealed that 7% of the variance across all structural asymmetries was explained by case-control status. Subtle case-control differences of brain macro-structural asymmetry may reflect differences at the molecular, cytoarchitectonic or circuit levels that have functional relevance for the disorder. Reduced left middle temporal cortical thickness is consistent with altered left-hemisphere language network organization in schizophrenia.

## Introduction

Schizophrenia is a serious mental illness characterized by various combinations of symptoms that may include delusions, hallucinations, disorganized speech, affective flattening, avolition, and executive function deficits (1). Left-right asymmetry is an important feature of human brain organization for diverse cognitive functions – for example, roughly 90% of people present with a left-hemisphere dominance for language and right-handedness (2-5). A possible role of altered structural and functional brain asymmetry in schizophrenia has been studied for several decades (6-10). Theoretical work has especially focused on disrupted laterality for language in relation to disorganized speech perception and production – the former may sometimes result in auditory verbal hallucinations which are a relatively prevalent symptom (11-14). Individuals with schizophrenia have been reported to show decreased left-lateralized language dominance (15, 16), as well as an absence or even reversal of structural asymmetries of language-related regions around the Sylvian fissure (which divides the temporal lobe from the frontal and parietal lobes) (13, 17-19). Language disturbances such as idiosyncratic semantic associations or reduced grammatical complexity are also commonly reported (20). Furthermore, the rate of non-right-handedness in schizophrenia is elevated compared to the general population (13, 21-25). Interestingly, some genomic loci that influence aspects of structural brain asymmetry or hand preference overlap with those associated with schizophrenia (26-29). Thus, there might be an etiological link between altered brain asymmetry and schizophrenia.

However, alterations in structural asymmetry of the cerebral cortex in schizophrenia have so far only been reported in studies with relatively small samples (13, 17-19, 30-36); to our knowledge, the largest case-control sample consisted of 167 affected individuals and 159 controls (33). Many of the existing findings are inconsistent and/or remain unreplicated, which is possibly due to low statistical power which limits the sensitivity to detect true effects, and also increases the risk of overestimating effect sizes (37-39). The reproducibility of findings may be further affected by the heterogeneity of clinical and demographic characteristics across studies. Moreover, varying approaches to process and analyze magnetic resonance imaging (MRI) data limit the possibility to reproduce results and/or to perform meta-analyses. For example, in studies targeting specific regions of interest, regions have been inconsistently defined, while studies that involved cortex-wide mapping used different image analysis protocols. Studies of subcortical volumetric asymmetries in schizophrenia have generally suffered from similar issues (40-42), with the notable exception of a study in 884 affected individuals and 1,680 controls that used a single image analysis pipeline (43). This study found an increased leftward asymmetry of the pallidum in schizophrenia (driven by a larger pallidum volume in the left hemisphere) compared to controls, which was also detectable in adolescents with subclinical psychotic experiences (43, 44).

The Enhancing Neuro Imaging Genetics through Meta-Analysis (ENIGMA, http://enigma.ini.usc.edu) consortium aims to perform large-scale analyses by combining imaging data from research groups across the world, processed with standardized protocols (45, 46). Previously, this consortium reported large-scale cortical thinning, smaller surface area, and altered subcortical volume in individuals with schizophrenia compared to controls (47, 48). However, asymmetry was not measured in these previous ENIGMA studies, and no tests were performed to assess whether case-control effects were different in the two hemispheres. The ENIGMA consortium has investigated structural brain asymmetries in other disorders (49): major depressive disorder (MDD) (50), autism spectrum disorder (ASD) (51), obsessive compulsive disorder (OCD) (52), and attention deficit/hyperactivity disorder (ADHD) (53). Case-control group-level effects were small for all of these disorders, with ASD showing the most widespread asymmetry differences – mostly involving regional cortical thickness measures – with a maximum Cohen’s *d* of 0.13 (51). Similar effect sizes may be anticipated for schizophrenia. Therefore, a large sample size is likely required to detect and accurately measure any effects. Although small group-average differences of brain macro-anatomy are unlikely to have clinical uses by themselves, they may help to identify brain regions and networks that have clinically relevant disruptions at other neurobiological levels – for example molecular or cytoarchitectonic – which can be investigated in future studies. Of note, the ENIGMA consortium has recently reported on asymmetry alterations with respect to subcortical *shape* (2,833 individuals with schizophrenia versus 3,929 controls), based on an automated approach quantifying local concave versus convex surface curvature (54), but that study did not address subcortical *volume* asymmetries, and omitted the cerebral cortex.

For the current study, we were able to measure both cortical and subcortical structural asymmetries in schizophrenia using by far the largest sample to date: 5,080 affected individuals and 6,015 controls, from 46 separate datasets. The datasets were collected originally as distinct studies over approximately 25 years, using different recruitment schemes, MRI scanning equipment and parameters. Importantly, for the current study, all primary MRI data were processed through a single pipeline for cortical atlas-based segmentation/subcortical parcellation and quality control.

Given previous theoretical and empirical work linking schizophrenia to reduced language laterality and function (see above), we had a particular interest in whether typical structural asymmetries of the core cerebral cortical language network might be reduced in schizophrenia – this includes asymmetries of lateral temporal cortex and inferior frontal cortex (55). However, linguistic tasks can also recruit various other brain regions (56), while disrupted cognition in schizophrenia affects multiple domains beyond language (1). Our primary aim was therefore to map potentially altered structural asymmetry in schizophrenia across all cortical and subcortical regions, for a thorough and unconstrained mapping of brain asymmetry in schizophrenia, supported by our unprecedented sample size. We achieved this through separate region-by-region testing of case-control group average differences in asymmetry (followed by false discovery rate correction), where the testing was two-tailed, i.e. we allowed for either reductions, increases or even reversals of asymmetry in affected individuals compared to controls. Due to restrictions on sharing individual-level data for many of the primary datasets, case-control differences were first tested for each regional asymmetry index (AI) separately within each dataset, and effects were then combined across datasets using meta-analysis methodology.

We also performed various secondary/exploratory analyses of the data. We explored possible associations of structural brain asymmetries with medication use and other disorder-specific measures: age at onset, duration of illness, as well as total, positive, and negative symptom scores. In addition, we tested age-and sex-specific asymmetry differences. Finally, for 14 datasets for which individual-level data were available, we tested for a multivariate association of case-control status simultaneously with regional AIs across the brain.

Together, these analyses aimed to provide novel insights into the extent and mapping of structural brain asymmetry alterations in schizophrenia, and how they relate to key clinical variables.

## Methods and materials

### Datasets

Structural MRI data were derived from 46 separate datasets (45 case-control and one case-only) via researcher participation in the ENIGMA schizophrenia working group, totaling 11,095 individuals. Of these, 5,080 were affected with schizophrenia and 6,015 were unaffected controls (Table 1, Table S1A). The datasets came from various countries around the world and were collected over the last roughly 25 years (Fig. 1). For each of the datasets, all relevant local ethical regulations were complied with, and appropriate informed consent was obtained for all individuals. The present study was carried out under approval from the Ethics Committee Faculty of Social Sciences of Radboud University Nijmegen. Sample size-weighted mean age across datasets was 33.3 (range 16.2-44.0) years for individuals with schizophrenia and 33.0 (11.8-43.6) years for controls. Affected individuals and controls were 67% and 52% male, respectively. Diagnostic interviews were conducted by registered clinical research staff using different diagnostic criteria (either the Diagnostic and Statistical Manual of Mental Disorders [DSM]-III, DSM-IV, DSM-5 or International Classification of Diseases [ICD]-10), and hand preference was obtained through assessment scales (mainly the Edinburgh Handedness Inventory and Annett Handedness Scale) or self-report (Table S2). No controls had present or past indications of schizophrenia.

**Table 1.** Overview of the ENIGMA-Schizophrenia datasets.

| Dataset | Country | N Total | Individuals with schizophrenia |  | Unaffected Controls |  |
| --- | --- | --- | --- | --- | --- | --- |
|  |  |  | N M/F | Mean age (years) | N M/F | Mean age (years) |
| AMC | Netherlands | 405 | 180 / 26 | 22.2 | 130 / 69 | 23.5 |
| ASRB* | Australia | 429 | 177 / 86 | 38.6 | 79 / 87 | 39.3 |
| CAMH | Canada | 264 | 70 / 48 | 44.0 | 77 / 69 | 43.6 |
| CASSI* | Australia | 116 | 35 / 18 | 35.2 | 33 / 30 | 30.5 |
| CIAM | South Africa | 51 | 13 / 8 | 31.1 | 16 / 14 | 26.6 |
| CLING | Germany | 371 | 35 / 13 | 32.4 | 132 / 191 | 25.2 |
| COBRE* | United States | 143 | 60 / 13 | 37.4 | 50 / 20 | 35.7 |
| EdinburghEHRS | United Kingdom | 67 | 19 / 12 | 21.8 | 17 / 19 | 21.2 |
| EdinburghFunc | United Kingdom | 60 | 11 / 14 | 37.2 | 18 / 17 | 37.5 |
| EdinburghSFMH | United Kingdom | 76 | 23 / 12 | 37.5 | 23 / 18 | 38.2 |
| EONCKS* | South Africa | 200 | 74 / 34 | 34.2 | 51 / 41 | 31.9 |
| ESO* | Czech Republic | 80 | 20 / 20 | 29.5 | 20 / 20 | 29.1 |
| FBIRN | United States | 359 | 139 / 46 | 39.0 | 124 / 50 | 37.5 |
| FIDMAG | Spain | 283 | 124 / 36 | 39.6 | 54 / 69 | 37.5 |
| FOR2107 Marburg* | Germany | 403 | 23 / 14 | 37.2 | 143 / 223 | 34.0 |
| FOR2107 Muenster* | Germany | 163 | 4 / 4 | 33.4 | 60 / 95 | 27.0 |
| Frankfurt | Germany | 59 | 20 / 9 | 38.1 | 13 / 17 | 35.2 |
| GAP | United Kingdom | 209 | 85 / 37 | 27.5 | 32 / 55 | 25.9 |
| GIPSI | Colombia | 43 | 35 / 8 | 33.5 | - | - |
| GROUP | Netherlands | 271 | 59 / 29 | 28.2 | 83 / 100 | 30.1 |
| HMS | Germany | 101 | 32 / 14 | 28.4 | 28 / 27 | 35.4 |
| HUBIN | Sweden | 196 | 70 / 24 | 41.7 | 69 / 33 | 42.0 |
| Huilong | China | 333 | 133 / 112 | 25.5 | 49 / 39 | 27.7 |
| IGP* | Australia | 138 | 40 / 28 | 41.7 | 38 / 32 | 36.0 |
| IMH* | Singapore | 227 | 105 / 46 | 33.1 | 47 / 29 | 31.8 |
| JBNU | South Korea | 208 | 57 / 37 | 39.3 | 48 / 66 | 41.4 |
| KaSP | Sweden | 88 | 34 / 22 | 30.3 | 15 / 17 | 27.5 |
| Madrid | Spain | 105 | 17 / 4 | 16.2 | 59 / 25 | 11.8 |
| MCIC | United States | 311 | 113 / 35 | 32.9 | 101 / 62 | 31.4 |
| MPRC* | United States | 437 | 128 / 78 | 35.4 | 96 / 135 | 37.1 |
| OLIN* | United States | 868 | 174 / 138 | 37.7 | 310 / 246 | 37.6 |
| Osaka | Japan | 855 | 118 / 98 | 36.1 | 318 / 321 | 34.1 |
| Oxford | United Kingdom | 74 | 24 / 17 | 16.3 | 15 / 18 | 16.1 |
| PAFIP | Spain | 556 | 214 / 138 | 29.9 | 127 / 77 | 29.2 |
| RomeSL | Italy | 280 | 110 / 54 | 39.4 | 73 / 43 | 37.5 |
| RSCZ | Russia | 98 | 46 / 0 | 22.2 | 52 / 0 | 22.3 |
| SCORE | Switzerland | 205 | 117 / 44 | 25.5 | 17 / 27 | 25.5 |
| SNUH | South Korea | 80 | 18 / 22 | 22.9 | 20 / 20 | 22.6 |
| SWIFT | Switzerland | 37 | 17 / 7 | 34.2 | 5 / 8 | 29.3 |
| TOP | Norway | 522 | 130 / 89 | 32.0 | 159 / 144 | 35.4 |
| UCISZ* | United States | 57 | 22 / 5 | 42.9 | 23 / 7 | 41.4 |
| UMCU | Netherlands | 600 | 236 / 79 | 30.9 | 165 / 120 | 32.9 |
| UNIBA* | Italy | 143 | 54 / 19 | 33.5 | 28 / 42 | 26.6 |
| UNIMAAS | Netherlands | 66 | 21 / 10 | 28.3 | 24 / 11 | 28.1 |
| UPENN | United States | 370 | 105 / 72 | 38.9 | 90 / 103 | 36.4 |
| Zurich* | Switzerland | 88 | 45 / 15 | 30.5 | 18 / 10 | 32.5 |
| **Total/Mean** |  | 11,095 | 3,386 / 1,694 | 33.3 | 3,149 / 2,866 | 33.0 |

**Figure 1.**
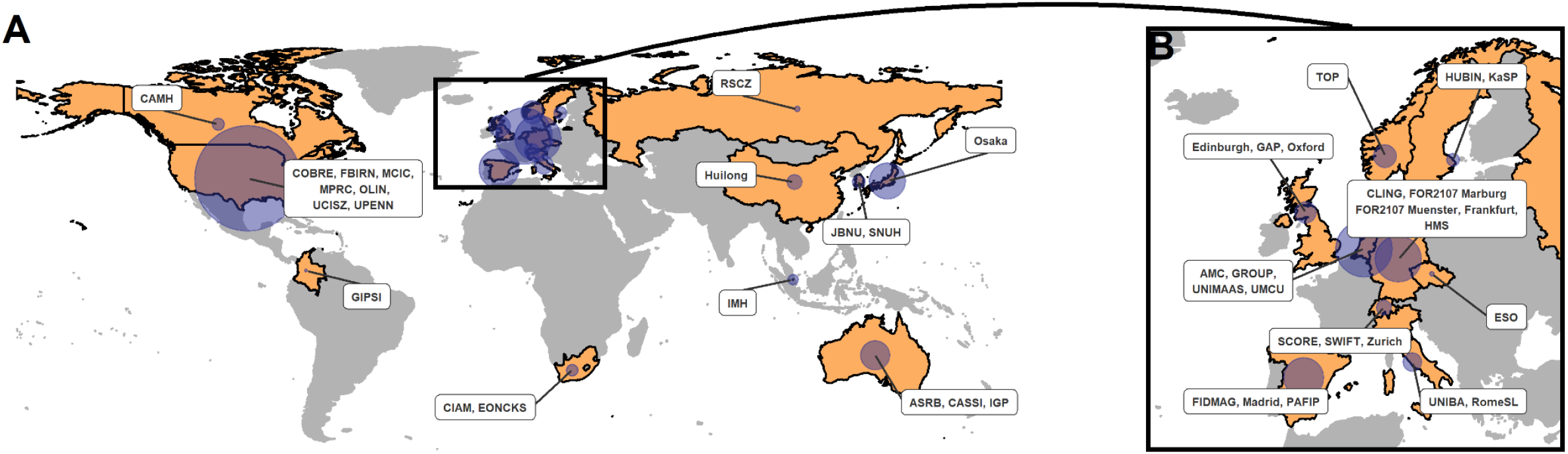
Geographic origin of included datasets. **A)** Countries from which one or more datasets originate are highlighted in the world map, with dataset names included in labels. The relative sample size of datasets per country is indicated by blue circles. **B)** Zoomed map of Europe. For more details, see Table 1. Figure generated in R using packages *ggplot2* (105), *rnaturalearth* (106), *sf* (107) and *ggrepel* (108).

### Image acquisition, processing and quality control

T1-weighted structural brain MRI scans were acquired at each study site. Dataset-specific scanner information, field strengths (1 T, 1.5 T, and 3 T), and image acquisition parameters are provided in Table S2. For data from all sites, image processing and segmentation were performed using FreeSurfer (see Table S2 for software versions) (57). For each individual, using the ‘recon-all’ pipeline, cerebral cortical thickness and surface area measures were derived for 34 bilaterally paired Desikan-Killiany (DK) atlas regions, as well as whole hemisphere-level average cortical thickness and surface area measures (58). Volumes for 8 bilaterally paired regions from a neuroanatomical atlas of brain subcortical structures (59) were derived using the ‘aseg’ segmentation command in FreeSurfer. A standardized ENIGMA quality control procedure was applied at each participating site (described in full here: http://enigma.ini.usc.edu/protocols/imaging-protocols/). Briefly, this included outlier detection in the derived cortical and subcortical measures and visual inspection of segmentations projected onto the T1-weighted image of each individual. For cortical measures, predefined guidelines for visual inspection were followed. Measurements from regions with poor segmentation were excluded, as well as individuals whose data failed overall quality checks. Data sharing limitations did not allow the central analysis group to have access to individual-level data for the majority of participating study sites. For further processing and analyses of the data, a script running in R software (R Foundation for Statistical Computing, Vienna, Austria, www.R-project.org) (60) was prepared and distributed among participating sites, to ensure coordinated collection of descriptive and summary statistics for subsequent meta-analysis by the central analysis team.

### Asymmetry index calculation

For each bilaterally paired brain regional measure, we used the left (*L*) and right (*R*) hemispheric measurements to calculate 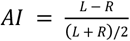, where the denominator corrects for automatic scaling of the index with the magnitude of the bilateral measure. This formula for AI calculation has been widely used (2, 52, 61-63). A negative value of the AI reflects a larger right hemispheric measurement (*R* > *L*), and a positive value a larger left hemispheric measurement (*L* > *R*). Distributions of AIs were plotted using histograms to allow for visual inspection. Left or right measurements equal to 0 were set to missing, as these most likely reflected data entry errors. Furthermore, when a left or right measurement was missing, the corresponding measurement in the opposite hemisphere was also set to missing. The standardized pipeline from raw image data through FreeSurfer does not introduce left-right flipping errors, but to ensure that such errors were not introduced during processing of raw imaging data by non-standard processes (e.g. during the conversion of DICOM to NIFTI files with bespoke scripts), we compared mean regional asymmetries for all datasets against grand sample-size adjusted means. If we noticed a large proportion of reversed average asymmetries for a dataset, we contacted the relevant site to re-check and correct their process (Table S3).

### Asymmetry differences between individuals with schizophrenia and unaffected controls

Group differences were examined separately for each brain regional AI and each case-control dataset, using univariate linear regression implemented in R. Our primary analysis model included diagnosis (case-control status) as the main binary predictor, and sex and age as covariates (model 1 in Supporting Information 1). For ten datasets where more than one scanner had been used (Table S2), we added *n*-1 binary dummy covariates (where *n* is the number of scanners in a given dataset), to statistically control for scanner effects. Sex was not included as a covariate for the RSCZ dataset, as this dataset included only males. Collinearity between predictor variables was assessed using the R-package *usdm* (v1-1.18) (64), and high collinearity (variance inflation factor > 5) was not found for any dataset. Linear regression analysis for any structural AI was not performed if the total sample size of a given dataset was lower than ten plus the number of scanner covariates, or if one of the diagnostic groups had a sample size lower than five. For each brain regional AI and each case-control dataset, we extracted the *t*-statistic for the ‘diagnosis’ term to calculate its corresponding Cohen’s *d* effect size, standard error and 95% confidence interval, using 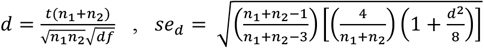, and 95% *CI* = [*d* -1.96\**se*_*d*_, *d* + 1.96\**se*_*d*_](65). In these equations, *d* is the Cohen’s *d* effect size, *t* is the *t*-statistic, *se* is the standard error, *n*_1_ is the number of unaffected controls, *n*_2_ is the number of individuals with schizophrenia, and *df* the degrees of freedom in the linear model.

### Random-effects meta-analysis

For each brain regional AI (Fig. S1-S3), effect sizes for ‘diagnosis’ from each case-control dataset were meta-analyzed in a random-effects model fitted with a restricted maximum likelihood (REML) estimator, using the function ‘rma’ in the R package *metafor* (v3.0-2) (66). Meta-analyzed effect sizes were projected on 3D meshes of inflated cortical or subcortical models from Brainder (www.brainder.org/research/brain-for-blender/), using Matlab R2020a (version 9.8.0.1323502; MathWorks, Natick, MA, USA). We calculated false discovery rate (FDR) corrected p-values using the Benjamini-Hochberg method to account for multiple tests (67) (i.e., separately for testing 35 cortical thickness AIs, 35 cortical surface area AIs, and eight subcortical volume AIs). Effects with *p*_FDR_ < 0.05 were considered statistically significant. For AIs that showed significant group differences between cases and controls, the group differences for the corresponding left and right measurements separately were also assessed *post hoc* (again using linear modelling with diagnosis, age and sex as predictors), to help describe the asymmetry differences.

### Sensitivity analyses

For any AI that showed a significant case-control group difference in the primary meta-analysis, we carried out three types of sensitivity analyses:

First, we identified datasets within which the 95% CI of the diagnosis effect did not overlap with the 95% CI of the meta-analyzed effect – using the ‘find.outliers’ function in the R package *dmetar* (v0.0.9) (68) – and then repeated the meta-analysis after excluding such outlier datasets.

Second, to assess whether between-dataset heterogeneity in effect sizes could be partly explained by known aspects of technical, diagnostic or geographic variability between datasets, we applied meta-regression and the Cochran’s Q test. As possible moderators we tested scanner strength, scanner manufacturer, use of a single scanner versus multiple scanners, image slice orientation, FreeSurfer version, diagnostic tool, and geographic origin of datasets (ethnicity was not recorded). See Table S2 for more information on these possible moderators.

Third, we applied models that included the same covariates as the primary analysis, but additionally included either handedness (right-handed vs. non-right-handed), intracranial volume (ICV), both handedness and ICV, or age^2^ (models 2-5 in Supporting Information 1).

### Medication group differences

For AIs that showed significant case-control group differences in the primary analysis, we explored associations with antipsychotic medication use at the time of scanning, through between-group comparisons of AIs of unmedicated individuals with schizophrenia, affected individuals taking only first-generation (typical) antipsychotics, affected individuals taking only second-generation (atypical) antipsychotics, and those taking both first-and second-generation antipsychotics. Sex and age were included as covariates (model 6 in Supporting Information 1) and derived Cohen’s *d* effect sizes were again meta-analyzed across datasets in a random-effects model. Applying a minimum group size threshold of 5 within any given dataset, sufficient data on the presence/absence of antipsychotic medication use for at least one comparison were available for 31 of the datasets (Table S1B), and the sample sizes for each between-group comparison are in Table S9. We calculated FDR corrected p-values to correct for all of the multiple subgroup comparisons and structural asymmetries tested.

### Correlations of asymmetries with clinical variables

For AIs that showed significant case-control group differences in the primary analysis, we assessed relationships between these AIs and clinical variables within affected individuals only: age at onset, duration of illness, chlorpromazine equivalent medication dose (at the time of scanning), as well as positive, negative, and total symptom severity scores from the Positive and Negative Symptom Scale (PANSS) (69), or the Scale for the Assessment of Positive Symptoms (SAPS) (70) and Scale for the Assessment of Negative Symptoms (SANS) (71) (separately depending on data availability, see Table S1A). Partial correlations between brain AIs and these quantitative measures were estimated using the ‘pcor.test’ function in the R package *ppcor* (v1.1) (72). Age and sex were included as covariates (model 7 in Supporting Information 1). The same minimum sample size requirement for dataset inclusion was applied as in the linear regression analyses (above). Correlation coefficients were meta-analyzed across datasets in a mixed-effects model including dataset as a random effect. We calculated FDR corrected p-values to control for all of the clinical variables and structural asymmetries tested. Sample sizes for each model are shown in Table S10.

### Secondary analysis of age-or sex-specific effects

For all AIs in all case-control datasets we applied models which were the same as the primary analysis but additionally included either diagnosis-by-age or diagnosis-by-sex interaction terms. We then carried out meta-analyses of the interaction effect estimates across datasets to assess possible AI differences between affected individuals and controls that were relatively specific to either males or females, or differed with age (models 8-9 in Supporting Information 1). In the same way as our primary analysis, we calculated FDR corrected p-values to account for multiple regional asymmetries tested.

### Multivariate analysis of case-control asymmetry differences

To examine case-control group differences across all brain regional AIs simultaneously in one model, we conducted a multivariate analysis based on 14 datasets for which individual-level data were available to the central analysis team. For this analysis, we only retained individuals with complete data for all bilateral measures of cortical and subcortical structures, which were 935 individuals affected with schizophrenia and 1,095 unaffected controls (Table S1C). We separately adjusted the left and right measurements using ComBat harmonization (an empirical Bayesian method) to remove dataset effects (73), where each dataset (and each scanner within multi-scanner datasets) was treated as a distinct ‘batch’. Diagnosis, age and sex were used as covariates when finding the data harmonization parameters in ComBat. After ComBat adjustment, one additional control individual was removed due to being assigned a negative corrected right hemisphere lateral ventricle volume (Fig. S4). AIs for cortical and subcortical measures were then calculated using the same formula as above, and collinearity between AIs was assessed by calculating a correlation matrix. AIs did not show higher pairwise correlations than 0.5 (Fig. S5-S6) A multivariate analysis of covariance (MANCOVA) using the ‘manova’ function in R was applied, testing all 76 regional structural brain AIs simultaneously against case-control status, with age and sex as covariates. We ran one million label-swapping permutations of case-control labels and calculated a permutation *p*-value by assessing the number of times the *F*-statistic of an analysis with permuted data was equal to or larger than the *F*-statistic of the analysis with real data, divided by the total number of permutations. When permuting case-control labels, we conserved case-control numbers within each dataset (and within scanner for multi-scanner datasets). To help interpret the MANCOVA results, we also derived univariate case-control association statistics for each separate structural AI from the multivariate association analysis output, using ANCOVA (‘summary.aov’ function in R).

## Results

### Asymmetry differences between individuals with schizophrenia and unaffected controls

In our primary analysis (model 1), total hemispheric average cortical thickness asymmetry (*d* = -0.053, *z* = -1.92, *p* = 0.055) and surface area asymmetry (*d* = 0.027, *z* = 1.23, *p* = 0.22) did not significantly differ between affected individuals and controls. At a regional level (Fig. 2, Table S4, Fig. S1-S3), there was a small but significant case-control difference in cortical thickness asymmetry of the rostral anterior cingulate cortex (*d* = -0.083, *z* = -3.21, *p* = 1.3 × 10^−3^, *p*_FDR_ = 0.047, reversal from leftward average asymmetry in controls to rightward average asymmetry in cases), and also in cortical thickness asymmetry of the middle temporal gyrus (*d* = -0.074, *z* = -2.99, *p* = 2.8 × 10^−3^, *p*_FDR_ = 0.048, increased average rightward asymmetry in cases) (Fig. 3, Fig. S7-S8, Table S5). *Post hoc* analysis of unilateral effects showed that both of these regional asymmetry differences were driven primarily by thinner left than right cortex in individuals with schizophrenia compared to controls (Table 2, Table S6). The middle temporal cortex is a core language network region (56), and left-hemisphere thinning is compatible with disrupted leftward laterality of brain organization for language in schizophrenia (10, 11). Nominally significant regional case-control associations (i.e. which did not survive multiple testing correction), were found for the AIs of inferior parietal cortex thickness, cuneus surface area, parahippocampal gyrus surface area, and nucleus accumbens volume (Fig. 3, Table S5).

**Figure 2.**
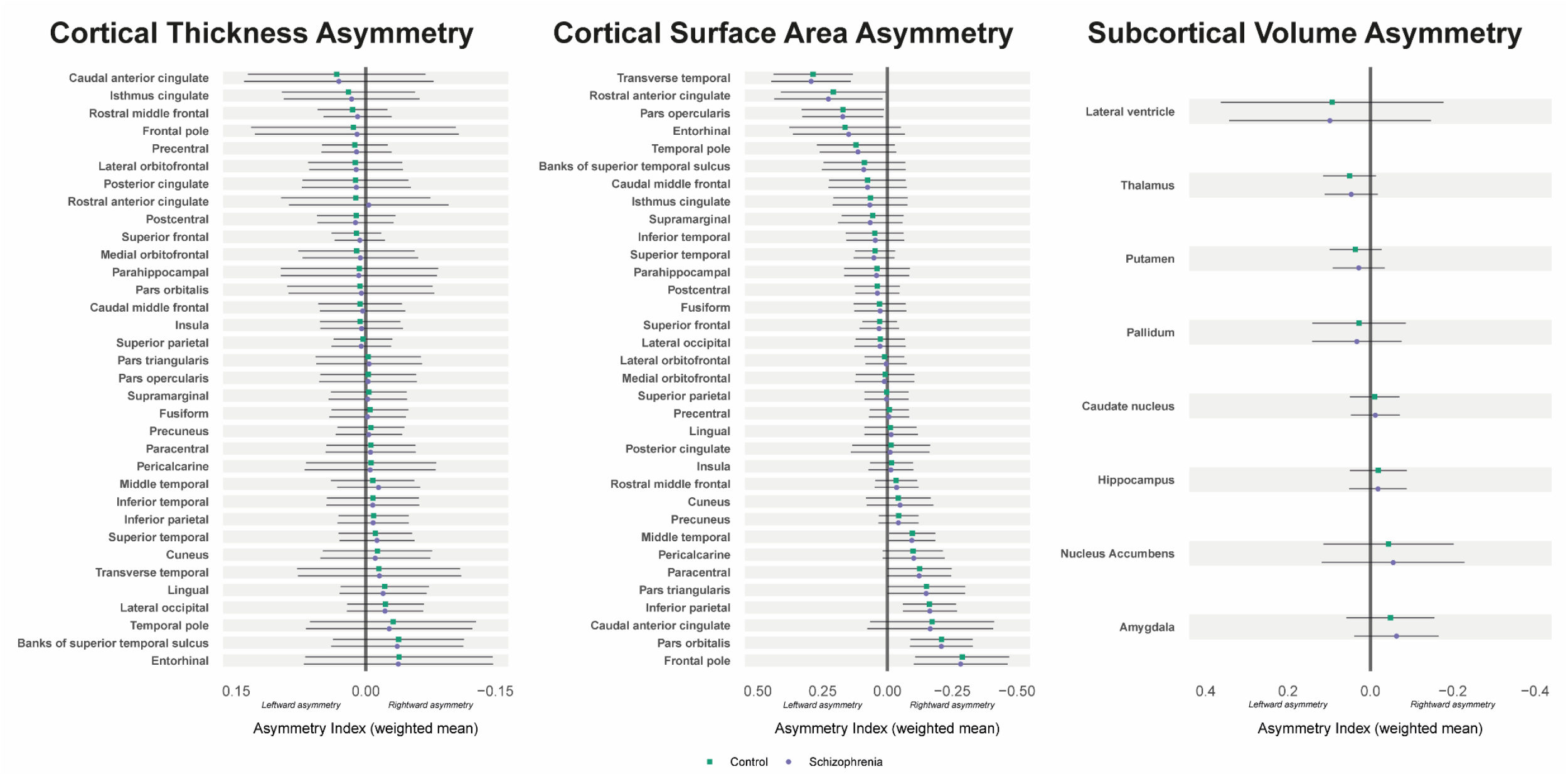
Average structural asymmetries of the brain in individuals with schizophrenia and unaffected controls. For each bilaterally paired structural measure, the mean asymmetry index (AI) across datasets, weighted by sample size, is shown for individuals with schizophrenia (purple) and unaffected controls (green). A positive AI indicates left > right asymmetry, whereas a negative AI indicates right > left asymmetry. Error bars show pooled standard deviations. Figure generated in R using package *ggplot2* (105).

**Figure 3.**
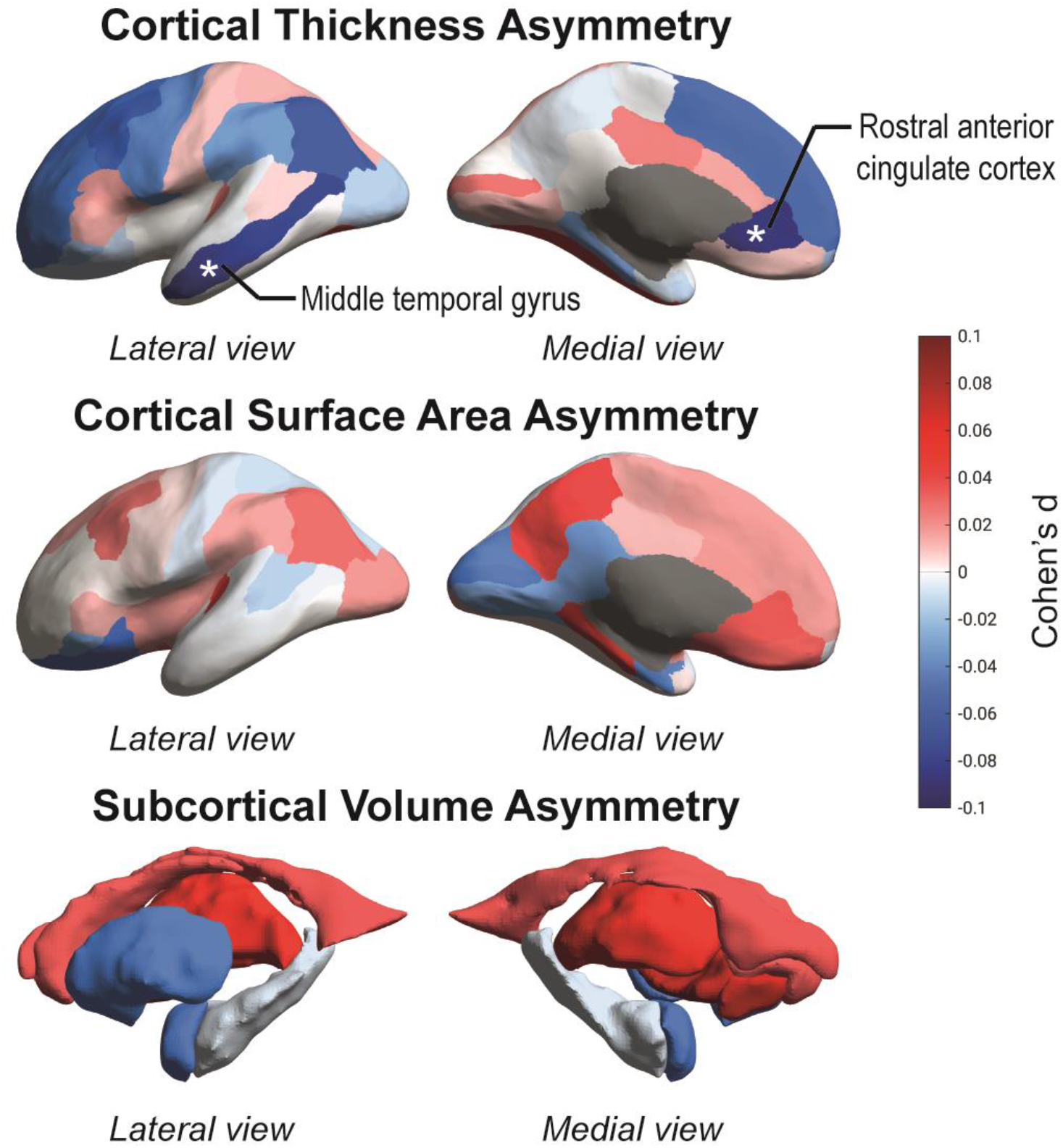
Map of cortical and subcortical asymmetry differences between individuals with schizophrenia and unaffected controls. Cohen’s *d* effect sizes from random-effects meta-analysis are projected on inflated left hemisphere cortical surface models (for cortical thickness and surface area) or subcortical structures (for subcortical volumes). Positive effects are shown in red shades (larger leftward or smaller rightward asymmetry in cases versus controls), while negative effects are shown in blue shades (smaller leftward or larger rightward asymmetry in cases versus controls). Gray shades indicate masked out structures. See also Fig. 2 and Table S4 for directions of effects. Regions significant at *p*_FDR_ < 0.05 are labelled and marked with asterisks.

**Table 2.** Significant brain regional asymmetry differences between individuals with schizophrenia and unaffected controls.

| Structural AI | Sample size (N) |  | Mean AI (SD) |  | Cohen's *d* effect size [95% CI] |  |  | Average asymmetry |  |
| --- | --- | --- | --- | --- | --- | --- | --- | --- | --- |
|  | Control | Schizophrenia | Control | Schizophrenia | Left | Right | AI | Control | Schizophrenia |
| **Rostral anterior cingulate cortex**<br>**(cortical thickness asymmetry)** | 5,811 | 4,851 | 0.012 (0.086) | -0.0035 (0.092) | -0.20<br>[-0.28, -0.11] | -0.094<br>[-0.15, -0.036] | -0.083<br>[-0.13, -0.032] | Leftward | Reversed to<br>rightward |
| **Middle temporal gyrus**<br>**(cortical thickness asymmetry)** | 5,673 | 4,684 | -0.0080 (0.048) | -0.015 (0.048) | -0.41<br>[-0.50, -0.32] | -0.36<br>[-0.44, -0.27] | -0.074<br>[-0.12, -0.026] | Rightward | Increased<br>rightward |

Mean AI = weighted mean asymmetry index across datasets. SD = pooled standard deviation across datasets (positive mean indicates average leftward asymmetry; negative mean indicates average rightward asymmetry). Cohen's *d* effect sizes are shown from separate meta-analysis of left-hemisphere, right-hemisphere and asymmetry index differences between cases and controls. No regional measures of cortical surface area asymmetry or subcortical volume asymmetry showed significant case-control differences after false discovery rate correction.

### Sensitivity analyses

For rostral anterior cingulate thickness asymmetry, there were three datasets in the primary meta-analysis which had outlier case-control effect sizes when compared to the meta-analyzed effect (see Methods). After excluding these datasets and repeating the meta-analysis for this AI, the case-control difference remained, with the same directionality (*d* = -0.073, *z* = -3.51, *p* = 4.5 × 10^−4^) (Table S7). For middle temporal gyrus thickness asymmetry, the exclusion of two outlier datasets also yielded a similar result compared to the primary analysis (*d* = -0.079, *z* = -3.44, *p* = 5.9 × 10^−4^), again with the same directionality (Table S7).

Meta-regression analysis did not identify any significant moderators (no Cochran’s Q omnibus test p-values < 0.05) (Fig. S9-S22), i.e. Cohen’s *d* effect sizes reflecting asymmetry differences between individuals with schizophrenia and unaffected controls were not significantly influenced by scanner strength, scanner manufacturer, use of a single scanner versus multiple scanners, image slice orientation, FreeSurfer version, diagnostic tool, or the geographic origin of datasets.

In models that included either handedness, ICV, both handedness and ICV, or age^2^ as additional covariates (models 2-5), the case-control differences for both of these regional AIs remained nominally significant, with similar directions and magnitudes of effect compared to the case-control differences found in the primary analysis (Table S8), despite differences in sample sizes resulting from limited availability of some of these variables.

### Medication group differences

Rostral anterior cingulate thickness asymmetry did not differ between affected individuals across medication groups (model 6) (Table S9). For the middle temporal gyrus, there was a nominally significant increase in average rightward asymmetry in affected individuals taking first generation versus second generation antipsychotics at the time of scanning (*d* = -0.21, *z* = -2.56, *p* = 0.011, *p*_FDR_ = 0.13), i.e., this was not significant after multiple testing correction (Table S9).

### Correlations of asymmetries with clinical variables

We found nominally significant correlations between rostral anterior cingulate thickness asymmetry and negative symptom severity measured with SANS (*r* = 0.049, *z* = 2.08, *p* = 0.038, *p*_FDR_ = 0.32, decreased rightward asymmetry with higher negative symptom rate) (Table S10A) and between middle temporal gyrus thickness asymmetry and duration of illness (*r* = -0.048, *z* = -1.97, *p* = 0.049, *p*_FDR_ = 0.32, increased rightward asymmetry with longer duration of illness) (Table S10B), but these correlations did not remain significant when correcting for multiple testing. No correlations with chlorpromazine-equivalent medication dose, age at onset, PANSS scores (total or positive and negative subscales), or SAPS or SANS scores, were found for either the rostral anterior cingulate thickness asymmetry or middle temporal gyrus thickness asymmetry (Table S10).

### Age-and sex-specific effects

In secondary analyses across all AIs using models with interaction terms, we found a significant diagnosis-by-age interaction (model 8) for pallidum volume asymmetry (*d* = 0.081, *z* = 3.26, *p* = 1.1 × 10^−3^, *p*_FDR_ = 9.0 × 10^−3^, stronger leftward asymmetry with higher age in cases) (Table S11-S12A, Fig. S23). This association was driven by a significantly decreased average leftward asymmetry with increasing age in controls (*r* = -0.077, *p* = 1.1 × 10^−3^) that was not present in affected individuals (Table S12B; Fig. S24). In terms of the corresponding unilateral effects, left and right pallidum volume decreased with increasing age in individuals with schizophrenia (L: *r* = -0.17, *p* = 4.7 × 10^−9^; R: *r* = -0.20, *p* = 4.7 × 10^−21^) and unaffected controls (L: *r* = -0.27, *p* = 2.1 × 10^−22^; R: *r* = -0.24, *p* = 6.2 × 10^−17^), but the two groups differed with respect to the side showing the stronger effect (Table S12B). No significant sex-by-diagnosis interactions were found (model 9) (Table S13).

### Multivariate analysis of case-control asymmetry differences

Considering all 76 regional structural brain AIs simultaneously in a multivariate model, applied to the 14 datasets for which individual-level data were available to the central analysis team (935 affected individuals and 1,094 controls), there was a significant multivariate structural brain asymmetry difference between cases and controls that accounted for roughly 7% of the variance considered across all 76 AIs (Wilks’ Λ = 0.932, approximate *F*(76, 1950) = 1.87, *p* = 1.25 × 10^−5^). Only three of the *F*-statistics resulting from one million label-swapping permutations (see Methods) were larger than the *F*-statistic from the true analysis, resulting in a permutation *p* = 3.0 × 10^−6^. We also derived univariate (ANCOVA) association statistics from the multivariate model to understand which AIs contributed most to the significant multivariate association. The structural AIs that showed nominally significant, univariate case-control differences in the 14 datasets available for this analysis were those for pallidum volume, nucleus accumbens volume, and eight regional surface area or thickness measures distributed widely over the cerebral cortex (Table 3). These did not include the two cortical regional AIs that showed significant case-control differences in the meta-analysis over all 45 case-control datasets, but did include AIs of other language-related regions of the temporal lobe: superior temporal sulcus surface area asymmetry and transverse temporal gyrus thickness asymmetry (Table 3). The large differences in overall sample size and contributing datasets between the multivariate analysis and main meta-analysis are a likely cause of these somewhat different results.

**Table 3.**
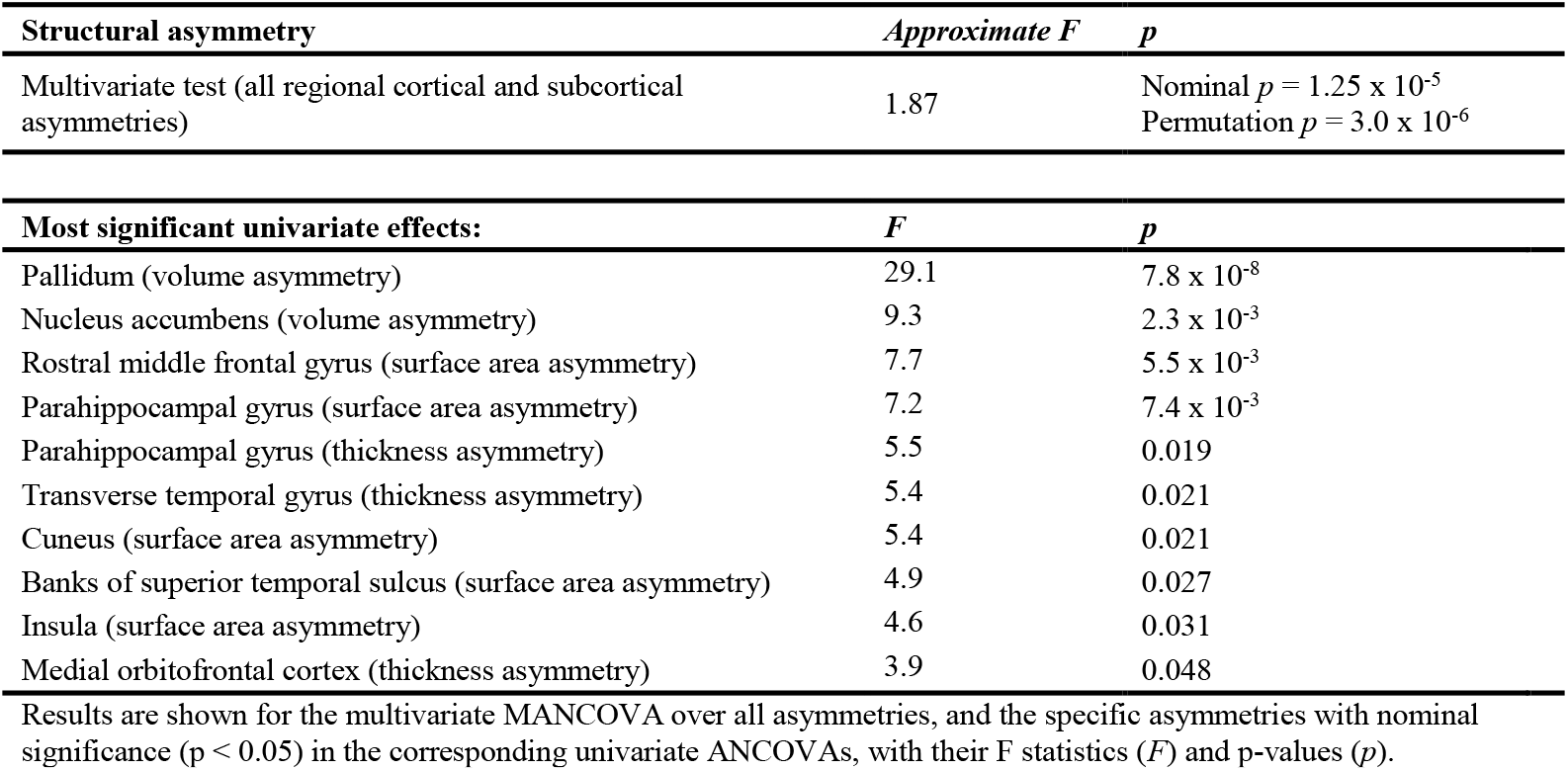
Multivariate analysis of case-control brain asymmetry differences between 935 individuals with schizophrenia and 1,094 controls for which individual-level data were available (14 datasets).

## Discussion

In this study, we investigated group differences in structural brain asymmetries between individuals with schizophrenia and unaffected controls, in the largest sample to date. The large sample size offered unprecedented statistical power to identify group differences based on the clinical diagnosis of schizophrenia, and to measure their effect sizes (37-39). Subtle differences of regional asymmetry were found for rostral anterior cingulate thickness, middle temporal gyrus thickness, and pallidum volume (the latter in older individuals). The Cohen’s *d* effect sizes were less than 0.1; i.e., very small (74). In light of previous large-scale analyses of bilateral cortical and subcortical alterations in schizophrenia (47, 48), our results suggest that morphometric alterations in this disorder are largely the same for the left and right hemispheres, involving only subtle asymmetrical effects at the group average level. This suggests that effect sizes of brain asymmetry differences in schizophrenia reported in earlier, much smaller studies (see Introduction), are likely to have been overestimated. Nonetheless, in a multivariate context, 7% of the total variance across all regional asymmetries was explained by case-control status, indicating a diffuse and subtle alteration of brain asymmetry in schizophrenia.

Subtle group differences of asymmetry in terms of macro-anatomic features, such as those studied here, may reflect effects at other neurobiological levels that have functional relevance for disorder symptoms – for example molecular, cytoarchitectonic and/or circuit levels (75-77). For example, cortical thickness measures can correlate with the degree of myelination (78), such that quantitative neuroimaging methods that are more sensitive to microstructural tissue content may reveal alterations in the regions implicated by this study. Neurite orientation dispersion and density imaging can be used to study cortical microstructural asymmetries (77), or the ratio of T1w and T2w images in grey matter can indicate cortical myelin content (79). We suggest that future studies using such techniques can be focused on the regions identified in this study. In addition, post mortem studies of hemispheric differences in gene expression in schizophrenia are motivated.

The middle temporal gyrus is prominently involved in the brain’s language network (56), so that our finding of lower left-sided cortical thickness in schizophrenia in this region is broadly consistent with a prominent theory in the literature: that left-hemisphere language dominance may be reduced in this disorder (10, 11). Cortical thinning of the left-hemispheric middle temporal gyrus has been associated with auditory verbal hallucinations in schizophrenia (80), and is reported in individuals with first-episode schizophrenia and high familial risk for the disorder (81, 82). In terms of grey matter volume, an opposite pattern (reduced right, increased left) has been reported for the middle temporal gyrus in putatively at-risk children compared to typically developing children (83). However, volume measures confound cortical thickness and surface area, and since these two aspects of cortical anatomy are known to vary substantially independently (28, 84, 85), it is unclear how these earlier volume-based findings may relate to the present findings based on cortical thickness asymmetry. Again, earlier findings in smaller samples may have been false positives with over-estimated effect sizes.

The rostral anterior cingulate cortex is an important hub in emotional and cognitive control (86), both of which are often affected in schizophrenia. In this region we observed a thinner left-sided cortex in affected individuals than controls on average, which was more pronounced than on the right side. This may be consistent with a previous study where adolescent/young adult relatives of individuals with schizophrenia showed a longitudinal decline of gray matter volume in the left rostral anterior cingulate cortex compared to controls (87). It is therefore possible that asymmetrical differences in this region emerge before schizophrenia onset, although the previous study included only 23 relatives, so its reported effects remain equivocal, and it used volume rather than thickness measures. In the present study, we saw no evidence for an age*diagnosis interaction effect for this regional thickness asymmetry, which is consistent with a pre-onset alteration that subsequently remains stable through adulthood.

Multivariate analysis in 14 of the datasets, for which individual-level data were available, resulted in a highly significant case-control difference. Various regional asymmetries contributed to this multivariate association, with pallidum volume asymmetry showing the largest individual contribution. Pallidum volume asymmetry was especially associated with schizophrenia in older individuals, as observed in secondary testing of univariate interaction models across all 45 case-control datasets. Larger pallidum volume in schizophrenia compared to controls – with a stronger effect in the left hemisphere – has been reported before (43, 44, 48, 88), although some datasets in our analysis partly overlapped with three of these studies (43, 44, 48). An age-dependent relationship between familial risk for schizophrenia and larger left pallidum volume has also been described in a small study of young adults (89) – this suggests that alterations of pallidum asymmetry might already be present in a prodromal stage of the disease. However, in the present study, the group difference in pallidum volume was absent in younger individuals and became more apparent in older adults. This also explains why the association was not significant in the primary univariate meta-analysis of all datasets together, i.e. it was driven by a subset of datasets that included older individuals, and that were also available for multivariate analysis (Fig. S24). The pallidum is prominently involved in reward and motivation (90), and impaired reward anticipation and a loss of motivation are well-known negative symptoms of schizophrenia (91). However, how pallidum structural asymmetry may relate to functional disorder-relevant changes remains unknown.

Various brain regional asymmetries have shown significant heritability in a recent genome-wide analysis of general population data (28), including rostral anterior cingulate thickness asymmetry and pallidum volume asymmetry (but not middle temporal gyrus thickness asymmetry). When polygenic risk for schizophrenia was assessed with respect to these heritable asymmetries in a multivariate analysis (29), one of the strongest associations was with rostral anterior cingulate thickness asymmetry. The direction of that effect was consistent with the present study, i.e. a rightward shift of asymmetry with increased polygenic risk for schizophrenia. In contrast, pallidum volume asymmetry showed little relation to schizophrenia polygenic risk (29), suggesting non-heritable contributions to this association. These genetic findings were established with adult general population data (UK Biobank) (29), but together with the current case-control findings, they indicate that altered rostral anterior cingulate thickness asymmetry may be a link between genetic susceptibility and disorder presentation. Left-right asymmetry of the brain originates during development *in utero* (75, 92-97), and specific genomic loci that affect brain asymmetry have recently been identified (28, 98). Some of the implicated genes may be involved in patterning the left-right axis of the embryonic or fetal brain, and genes expressed at different levels on the left and right sides of the embryonic central nervous system were found to be particularly likely to affect schizophrenia susceptibility (92). However, other genes may affect brain asymmetry as it changes throughout the lifespan (2, 99) and therefore may affect susceptibility to asymmetry-associated disorders later in life.

The magnitudes of effects in this study were in line with those reported in recent large-scale studies of brain asymmetry in other psychiatric disorders carried out through the ENIGMA consortium (50-53). In ASD, a similar decreased leftward asymmetry of rostral anterior cingulate thickness was reported (51) – this region is important in cognitive control which can be impaired in both schizophrenia and ASD. For ADHD, a nominally significant increase in rightward asymmetry of middle temporal gyrus thickness was reported, while in adults specifically, less leftward asymmetry of pallidum volume was found (53). The former finding is consistent in its direction of effect with the present study, while the latter is opposite. For OCD, the pallidum was found to be less left lateralized in cases versus controls in a pediatric dataset and this effect was again opposite to our current findings in older individuals with schizophrenia (52). These cross-disorder comparisons suggest that clinical and etiological similarities and differences between schizophrenia and other psychiatric disorders might be partly reflected in asymmetry alterations involving some of the same brain regions. For further discussion of brain asymmetry alterations across multiple psychiatric traits, see Mundorf et al. (100).

Schizophrenia is a highly heterogeneous disorder covering a range of possible symptoms, which may correspond to differing underlying disease mechanisms. Our primary analysis only considered case-control group average differences based on the overall diagnosis of schizophrenia, and in secondary analyses, we did not find significant correlations of asymmetries with major clinical variables within cases after adjusting for multiple testing -including age at onset, duration of illness, and symptom scores. Furthermore, data for several variables were only available from a limited number of study sites (medication, handedness, clinical variables), reducing the sample size and thus statistical power in these secondary analyses. More detailed clinical data would be useful to gather in future large-scale studies of structural asymmetries. For example, a future study could investigate middle temporal gyrus thickness asymmetry in relation to the presence and severity of auditory verbal hallucinations (note that PANSS question 3 does not distinguish between auditory, visual, olfactory or somatic types of hallucination, so a more targeted clinical assessment would be required).

This was the largest study of structural brain asymmetries in schizophrenia to date, and made use of a single image processing and analysis pipeline to support analysis across multiple datasets. The fact that we used data from a range of imaging equipment, diagnostic tools and regions of the world ensures generalizability of our findings, as they pertain to the diverse manner in which schizophrenia is diagnosed and studied internationally. Therefore, a major strength of our approach is in showing consensus effects across inter-site variations in techniques and samples. Unlike in a highly selected, single site or single equipment study, the broad and generalizable total dataset made it unlikely that any single factor confounded our findings. We used a meta-analytic approach after testing for effects separately within each dataset, where cases and controls were matched for technical and demographic factors within each dataset. This allowed us to assume and control for variations between datasets in our main analysis. In addition, meta-regression analyses indicated that between-dataset variability in technical, diagnostic or geographic aspects had no significant impact on the associations between schizophrenia and regional brain asymmetries identified in this study. It is also worth noting that several findings from the ENIGMA-Schizophrenia working group (not related to asymmetry) have been replicated by The Cognitive Genetics Collaborative Research Organization (COCORO) in a sample collected in Japan (101), supporting generalization of findings across populations.

We used cross-sectional datasets, limiting the possible interpretation with respect to cause-effect relations, longitudinal changes in asymmetry, or medication effects on asymmetry. Many of the individuals with schizophrenia were likely to be past or current users of medication, although data on medication were only available for a subset of datasets, and were also limited to medication use at the time of scanning. We found no evidence that the asymmetries of rostral anterior cingulate thickness or middle temporal gyrus thickness were different in affected individuals using medication versus those not using medication, which may indicate that the case-control differences of asymmetry that we detected had a developmental origin, rather than reflecting medication use. Indeed, medication effects on cortical thickness may be predominantly bilateral, without necessarily affecting asymmetry. We are not aware of any comparably sized prospective/randomized study in which medication effects could be disentangled from case-control effects.

We found a tentative difference of middle temporal gyrus thickness asymmetry between individuals who were taking first-generation versus second-generation antipsychotics. In principle this finding might reflect a change of asymmetry in response to first generation medication in particular, or else clinical differences of disorder presentation linked to asymmetry which then affect treatment choices. We saw nominally significant evidence that this same regional asymmetry relates to illness duration. However, the medication subgroup analyses were limited by relatively small sample sizes compared to the primary case-control analysis, and this particular association did not survive multiple testing correction. Also, medication status did not include information on previously used antipsychotics. This association therefore remains uncertain until replicated.

We used macro-anatomical brain atlases for both the cortical and subcortical structures, which is the most feasible approach for large-scale analysis across multiple datasets, but limits spatial resolution. With higher resolution mapping, regions that showed negative results in our study may harbor more focal case-control asymmetry differences, which could be revealed for example through vertex-wise cortical mapping (63, 98, 102), or subcortical partitioning into subfields or nuclei.

This study focused on group average differences, but individual-level deviations in affected individuals may be highly heterogeneous and not well captured by group-average approaches (103). Future studies may investigate individual or patient subgroup asymmetry deviations from a normative range or structural pattern, which may deliver clinical utility, for example through contributing to diagnosis or prognosis. This concept has shown promising results in recent studies even in smaller samples (103, 104). The small group-average effects that we identified in the present study are unlikely to have clinical utility when considered in isolation, although they may contribute to multivariate prediction models in future research, for example when considering brain features across multiple imaging modalities.

In summary, we performed the largest study of asymmetry differences between individuals with schizophrenia and unaffected controls to date. Effect sizes were small, but several regional case-control asymmetry differences in cortical thickness and subcortical volume were suggested, and multivariate analysis indicated that 7% of variation across all regional asymmetries could be explained by the case-control group difference. Our findings therefore support a long-standing theory that the brain’s asymmetry can be different in schizophrenia (10, 11), even if earlier studies in smaller samples were likely to have over-estimated the effect sizes in relation to structural asymmetry. Altered asymmetry in schizophrenia may conceivably occur during development through disruption of a genetically regulated program of asymmetrical brain development, and/or through different trajectories of lifespan-related changes in brain asymmetries. The specific regions implicated here provide targets for future research on the molecular and cellular basis of altered lateralized cognitive functions in schizophrenia, which may ultimately help to identify pathophysiological mechanisms.

## Data Availability

This study made use of 46 separate data sets collected around the world, under a variety of different consent procedures and regulatory bodies, during recent decades. Requests to access the data sets will be considered in relation to the relevant consents, rules and regulations, and can be made via the schizophrenia working group of the ENIGMA consortium: http://enigma.ini.usc.edu/ongoing/enigma-asd-working-group/

## Acknowledgments

The ENIGMA project is in part supported by the National Institute of Biomedical Imaging and Bioengineering of the National Institutes of Health (NIH) (Grant No. U54EB020403). The content is solely the responsibility of the authors and does not necessarily represent the official views of the NIH. D.S., M.C.P., S.E.F. and C.F. [ENIGMA-Laterality] were funded by the Max Planck Society (Germany). R.A-A. [PAFIP] is funded by a Miguel Servet contract from the Carlos III Health Institute (CP18/00003). J.V-B. [PAFIP] has received unrestricted research funding from Instituto de Investigación sanitaria Valdecilla (IDIVAL): INT/A21/10, INT/A20/04. D.A. [TOP] is funded by the South-Eastern Norway Regional Health Authority (grants 2019107 and 2020086). L.T.W. [TOP] is funded by The Research Council of Norway (223273, 300767), the South-Eastern Norway Regional Health Authority (2019101), and the European Research Council under the European Union’s Horizon 2020 research and Innovation program (ERC StG, Grant 802998). O.A.A. [TOP] is funded by the Research Council of Norway (#223273, #275054) and KG Jebsen Stiftelsen, South East Norway Health Authority (2017-112, 2019-108). P.K. [MPRC, Huilong] received support from NIH grants R01MH123163 and R01EB015611. M.J.G. [ASRB, IGP] received funding from National Health and Medical Research Council (NHMRC) Project Grants 630471, 1051672, 1081603. C.P. [ASRB] was supported by a NHMRC Senior Principal Research Fellowship (1105825), and NHMRC L3 Investigator Grant (1196508). V.D.C. [FBIRN, COBRE] was funded by NIH grants R01MH118695 and National Science Foundation (NSF) 2112455. J.M.F. [FBIRN] was funded by a Senior Research Career Scientist Award, Department of Veterans Affairs. P.F-C. [FIDMAG] is funded by Centro de Investigación Biomédica en Red de Salud Mental (CIBERSAM) and by Instituto de Salud Carlos III, co-funded by European Union (European Regional Development Fund (ERDF)/European Social Fund (ESF), ‘Investing in your future’): Sara Borrell Research contract (CD19/00149). G.S. [RomeSL] was funded by Italian Ministry of Health RC17-18-19-20-21/A grants. A.N.V. [CAMH] currently receives funding from the National Institute of Mental Health, Canadian Institutes of Health Research, Canada Foundation for Innovation, Centre for Addiction and Mental Health (CAMH) Foundation, and the University of Toronto. K.S. [IMH] received support from research grants from the National Healthcare Group, Singapore (SIG/05004; SIG/05028), and the Singapore Bioimaging Consortium (RP C009/2006). Y-C.C. [JBNU] was supported by a grant of the Korean Mental Health Technology R&D Project, Ministry of Health & Welfare, Republic of Korea (HL19C0015) and a grant of the Korea Health Technology R&D Project through the Korea Health Industry Development Institute (KHIDI), funded by the Ministry of Health & Welfare, Republic of Korea (HI18C2383). U.D. [FOR2107 Münster] was funded by the German Research Foundation (DFG, grant FOR2107 DA1151/5-1 and DA1151/5-2; SFB-TRR58, Projects C09 and Z02) and the Interdisciplinary Center for Clinical Research (IZKF) of the medical faculty of Münster (grant Dan3/012/17). J.M.S. [COBRE] was funded by NIH grant 1P20RR021938-01. A.R.M. [COBRE] received funding from NIH grants P30GM122734 and R01MH101512. C.M.D-C. [Madrid] has received funding from Instituto de Salud Carlos III, Spanish Ministry of Science and Innovation (PI17/00481, PI20/00721, JR19/00024). S.Ce. [KaSP] received funding from the Swedish Research Council (Grant No. 523-2014-3467). M.Kir. [Zurich] acknowledges funding from the Swiss National Science Foundation (P2SKP3_178175). T.H. [ESO] was supported by funding from the Canadian Institutes of Health Research (142255), Ministry of Health of the Czech Republic (16-32791A, NU20-04-00393), and Brain & Behavior Research Foundation Young and Independent Investigator Awards. A.Jam. [Oxford] was supported by Medical Research Council (MRC) grant G0500092. P.H. [SWIFT] is supported by a NARSAD grant from the Brain & Behavior Research Foundation (28445) and by a Research Grant from the Novartis Foundation (20A058). R.C.G. [UPENN] received funding through NIH grants 1R01MH117014 and 1R01MH119219. N.J. [ENIGMA] is funded by NIH grant R01MH117601. S.E.M. [ENIGMA] is supported in part by Australian NHMRC APP1172917. J.A.T. [FBIRN, MCIC, COBRE] is supported by NIH grant R01MH121246. Further acknowledgments specific to datasets are listed in the Supporting Information.

## Author Contributions

D.S., M.C.P., S.E.F., B.F., D.C.G., R.C.G., R.H., N.J., E.L., S.E.M., P.M.T., J.A.T., T.G.M.vE., and C.F. designed the research; D.S., M.C.P., M.F., J.M., K.M., S.M.C.dZ., N.E.M.vH., W.C., H.E.HP., R.S.K., R.A-A, V.O-GdlF., D.T-G., J.V-B., B.C-F., D.A., A.D., L.T.W., I.A., O.A.A., E.G.J., P.K., J.M.B., S.V.C., P.T.M., B.J.M., Y.Q., P.E.R., U.S., R.J.S., V.J.C., M.J.G., F.A.H., C.M.L., C.P., C.S.W., T.W.W., L.dH., K.B., J-K.P., K.G.R., F.St., A.Jan., T.T.J.K., I.N., B.K., O.G., T.D.S., J.B., D.H.M., A.P., V.D.C., J.M.F., S.G.P., J.C., Y.T., Z.W., H.X., F.F., F.B., S.E., P.F-C., MA.G-L., A.G-P., R.S., S.S., E.P-C., V.C., F.P., D.V., N.B., G.S., S.M., T.vA., E.W.D., A.N.V., K.S., S.Ci., P.D., R.M.M., W-S.K., Y-C.C., C.An., A.Sc., S.B., A.M.McI., H.C.W., S.M.L., S.dP., H.K.L., F.Sc., R.E., D.G., R.L., U.D., J.T.E., K.R-M., J.M.S., A.R.M., L.A.A., L.F., G.P., A.B., C.M.D-C., J.J., N.G.L., C.Ar., A.S.T., I.L., S.Ce., C.M.S., F.G., M.Kir., S.K., T.H., A.Sk., F.Sp., M.Kim, YB.K., S.O., JS.K., A.Jam., G.B., C.K., M.S., V.O., A.U., F.M.H., D.J.S., H.S.T., A.M.D-Z., J.A.P-Z., C.L-J., S.H., E.J., W.S., P.H., D.C.G., R.C.G., R.H., J.A.T., and T.G.M.vE. acquired data and/or analyzed data and/or performed research; D.S. and C.F. wrote the paper; D.S., M.C.P., M.F., J.M., K.M., S.M.C.dZ., N.E.M.vH., W.C., H.E.HP., R.S.K., R.A-A, V.O-GdlF., D.T-G., J.V-B., B.C-F., D.A., A.D., L.T.W., I.A., O.A.A., E.G.J., P.K., J.M.B., S.V.C., P.T.M., B.J.M., Y.Q., P.E.R., U.S., R.J.S., V.J.C., M.J.G., F.A.H., C.M.L., C.P., C.S.W., T.W.W., L.dH., K.B., J-K.P., K.G.R., F.St., A.Jan., T.T.J.K., I.N., B.K., O.G., T.D.S., J.B., D.H.M., A.P., V.D.C., J.M.F., S.G.P., J.C., Y.T., Z.W., H.X., F.F., F.B., S.E., P.F-C., MA.G-L., A.G-P., R.S., S.S., E.P-C., V.C., F.P., D.V., N.B., G.S., S.M., T.vA., E.W.D., A.N.V., K.S., S.Ci., P.D., R.M.M., W-S.K., Y-C.C., C.An., A.Sc., S.B., A.M.McI., H.C.W., S.M.L., S.dP., H.K.L., F.Sc., R.E., D.G., R.L., U.D., J.T.E., K.R-M., J.M.S., A.R.M., L.A.A., L.F., G.P., A.B., C.M.D-C., J.J., N.G.L., C.Ar., A.S.T., I.L., S.Ce., C.M.S., F.G., M.Kir., S.K., T.H., A.Sk., F.Sp., M.Kim, YB.K., S.O., JS.K., A.Jam., G.B., C.K., M.S., V.O., A.U., F.M.H., D.J.S., H.S.T., A.M.D-Z., J.A.P-Z., C.L-J., S.H., E.J., W.S., P.H., S.E.F., B.F., D.C.G., R.C.G., R.H., N.J., E.L., S.E.M., P.M.T., J.A.T., T.G.M.vE., and C.F. critically reviewed the paper prior to submission.

## Data availability

This study made use of 46 separate data sets collected around the world, under a variety of different consent procedures and regulatory bodies, during recent decades. Requests to access the data sets will be considered in relation to the relevant consents, rules and regulations, and can be made via the schizophrenia working group of the ENIGMA consortium: http://enigma.ini.usc.edu/ongoing/enigma-schizophrenia-working-group/.

