## Supporting Information for "Large-scale analysis of structural brain asymmetries in schizophrenia via the ENIGMA consortium"

### Supporting Information 1: Overview of statistical models for regression and partial correlation analyses

Below is an overview of the models for the linear regression and partial correlation analyses that were run by each participating site, and from which summary statistics for meta-analysis by the central analysis group were extracted. Model numbers refer to those indicated in the main manuscript text. The independent variable highlighted in **bold** is the predictor of interest in each model, for which effects were combined across datasets in random-effects meta-analysis.

Abbreviations used in models:

| Variable | Type | Description |
|---|---|---|
| AI | Continuous | Asymmetry index |
| Dx | Categorical (binary) | Diagnosis: schizophrenia or unaffected control |
| HAND | Categorical (binary) | Hand preference: right or non-right (left + ambidextrous) |
| ICV | Continuous | Intracranial volume |
| Scanner | Categorical (binary) | Optional covariate: If a site used multiple scanners to obtain images, *n*-1 binary dummy covariates (where *n* is the number of scanners in a given dataset) were added, to differentiate which scanner an individual's data came from. |
| AP-group | Categorical | Antipsychotic medication groups, tested as binary variables for between-group comparisons (see main text). |
| Clinical variable | Continuous | Schizophrenia-specific clinical variable. We included chlorpromazine-equivalent (CPZ) medication dose, age at onset, duration of illness, PANSS total score, PANSS positive symptom score, PANSS negative symptom score, SAPS score or SANS score. |

***Models to assess case-control differences***

Primary model:

[1] AI ~ **Dx** + Age + Sex (+ Scanner)

Primary model with additional covariates:

[2] AI ~ **Dx** + Age + Sex + HAND (+ Scanner)

[3] AI ~ **Dx** + Age + Sex + ICV (+ Scanner)

[4] AI ~ **Dx** + Age + Sex + HAND + ICV (+ Scanner)

[5] AI ~ **Dx** + Age + $Age^2$ + Sex (+ Scanner)

***Models to assess medication group differences***

Antipsychotic medication between-group comparisons within affected individuals:

[6] AI ~ **AP-group** + Age + Sex (+ Scanner)

***Models to assess correlations with clinical variables in affected individuals***

[7] Linear model: AI ~ **Clinical variable** + Sex + Age (+ Scanner)

Partial correlation: ρ(AI)(**Clinical variable**)·{Sex, Age, (Scanner)}

***Models to assess diagnosis-by-age and diagnosis-by-sex interactions, including correlations with age***

[8] AI ~ Dx + Age + Sex + **Dx*Age +** (Scanner)

[8b] Linear model: AI ~ **Age** + Sex (+ Scanner)

Partial correlation: ρ(AI)(**Age**)·{Sex, (Scanner)}

[9] AI ~ Dx + Age + Sex + **Dx*Sex** + (Scanner)

### Supporting figures

**Fig. S1 (page 6-14). Overall and per-dataset average and range for cortical thickness asymmetries.** For each cortical thickness asymmetry measure, the average in controls (green circles) and individuals affected with schizophrenia (purple squares) is shown. The top (highlighted) row contains the grand sample size-weighted mean and standard deviation (thick line segments). The other rows contain per-dataset averages, standard deviations and minimum and maximum values (indicated with thin line segments).

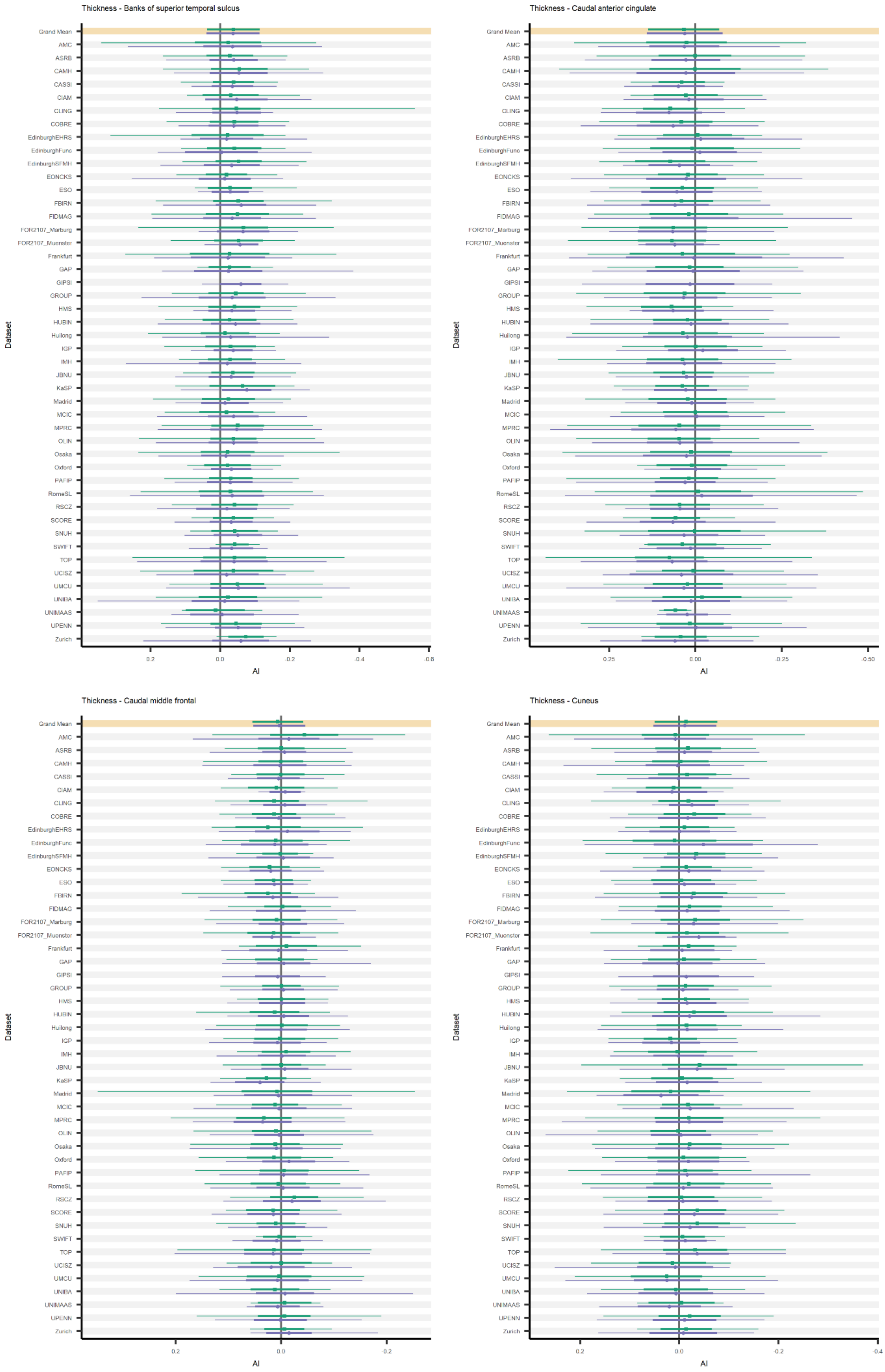

Thickness - Banks of superior temporal sulcus
Dataset
Grand Mean
AMC
ASRB
CAMH
CASSI
CIAM
CLING
COBRE
EdinburghEHRS
EdinburghFunc
EdinburghSFMH
EONCKS
ESO
FBIRN
FIDMAG
FOR2107_Marburg
FOR2107_Muenster
Frankfurt
GAP
GIPSI
GROUP
HMS
HUBIN
Huilong
IGP
IMH
JBNU
KaSP
Madrid
MCIC
MPRC
OLIN
Osaka
Oxford
PAFIP
RomeSL
RSCZ
SCORE
SNUH
SWIFT
TOP
UCISZ
UMCU
UNIBA
UNIMAAS
UPENN
Zurich
0.2
0.0
-0.2
-0.4
-0.6
AI
Thickness - Caudal anterior cingulate
Dataset
Grand Mean
AMC
ASRB
CAMH
CASSI
CIAM
CLING
COBRE
EdinburghEHRS
EdinburghFunc
EdinburghSFMH
EONCKS
ESO
FBIRN
FIDMAG
FOR2107_Marburg
FOR2107_Muenster
Frankfurt
GAP
GIPSI
GROUP
HMS
HUBIN
Huilong
IGP
IMH
JBNU
KaSP
Madrid
MCIC
MPRC
OLIN
Osaka
Oxford
PAFIP
RomeSL
RSCZ
SCORE
SNUH
SWIFT
TOP
UCISZ
UMCU
UNIBA
UNIMAAS
UPENN
Zurich
0.25
0.00
-0.25
-0.50
AI
Thickness - Caudal middle frontal
Dataset
Grand Mean
AMC
ASRB
CAMH
CASSI
CIAM
CLING
COBRE
EdinburghEHRS
EdinburghFunc
EdinburghSFMH
EONCKS
ESO
FBIRN
FIDMAG
FOR2107_Marburg
FOR2107_Muenster
Frankfurt
GAP
GIPSI
GROUP
HMS
HUBIN
Huilong
IGP
IMH
JBNU
KaSP
Madrid
MCIC
MPRC
OLIN
Osaka
Oxford
PAFIP
RomeSL
RSCZ
SCORE
SNUH
SWIFT
TOP
UCISZ
UMCU
UNIBA
UNIMAAS
UPENN
Zurich
0.2
0.0
-0.2
AI
Thickness - Cuneus
Dataset
Grand Mean
AMC
ASRB
CAMH
CASSI
CIAM
CLING
COBRE
EdinburghEHRS
EdinburghFunc
EdinburghSFMH
EONCKS
ESO
FBIRN
FIDMAG
FOR2107_Marburg
FOR2107_Muenster
Frankfurt
GAP
GIPSI
GROUP
HMS
HUBIN
Huilong
IGP
IMH
JBNU
KaSP
Madrid
MCIC
MPRC
OLIN
Osaka
Oxford
PAFIP
RomeSL
RSCZ
SCORE
SNUH
SWIFT
TOP
UCISZ
UMCU
UNIBA
UNIMAAS
UPENN
Zurich
0.2
0.0
-0.2
-0.4
AI

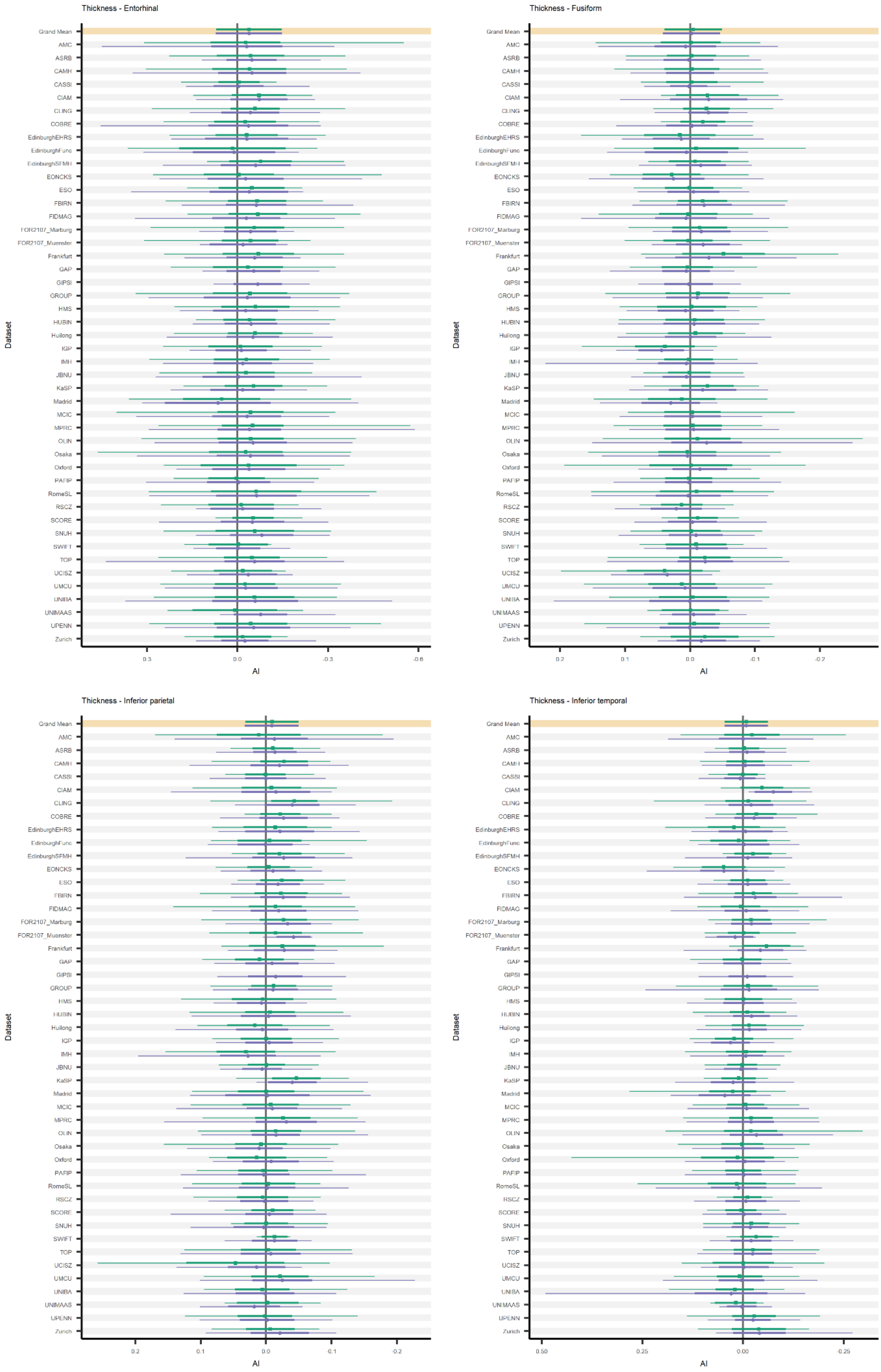
Thickness - Entorhinal
Grand Mean
AMC
ASRB
CAMH
CASSI
CIAM
CLING
COBRE
EdinburghEHRS
EdinburghFunc
EdinburghSFMH
EONCKS
ESO
FBIRN
FIDMAG
FOR2107_Marburg
FOR2107_Muenster
Frankfurt
GAP
GIPSI
GROUP
HMS
HUBIN
Huilong
IGP
IMH
JBNU
KaSP
Madrid
MCIC
MPRC
OLIN
Osaka
Oxford
PAFIP
RomeSL
RSCZ
SCORE
SNUH
SWIFT
TOP
UCISZ
UMCU
UNIBA
UNIMAAS
UPENN
Zurich
Dataset
0.3
0.0
-0.3
-0.6
AI
Thickness - Fusiform
Dataset
0.2
0.1
0.0
-0.1
-0.2
AI
Thickness - Inferior parietal
Dataset
0.2
0.1
0.0
-0.1
-0.2
AI
Thickness - Inferior temporal
Dataset
0.50
0.25
0.00
-0.25
AI

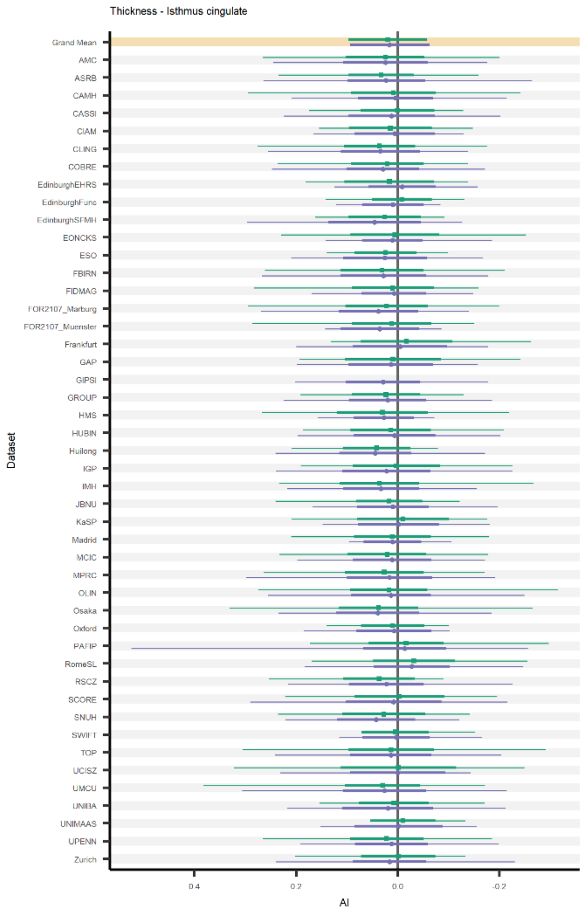
Thickness - Isthmus cingulate
Dataset
AI

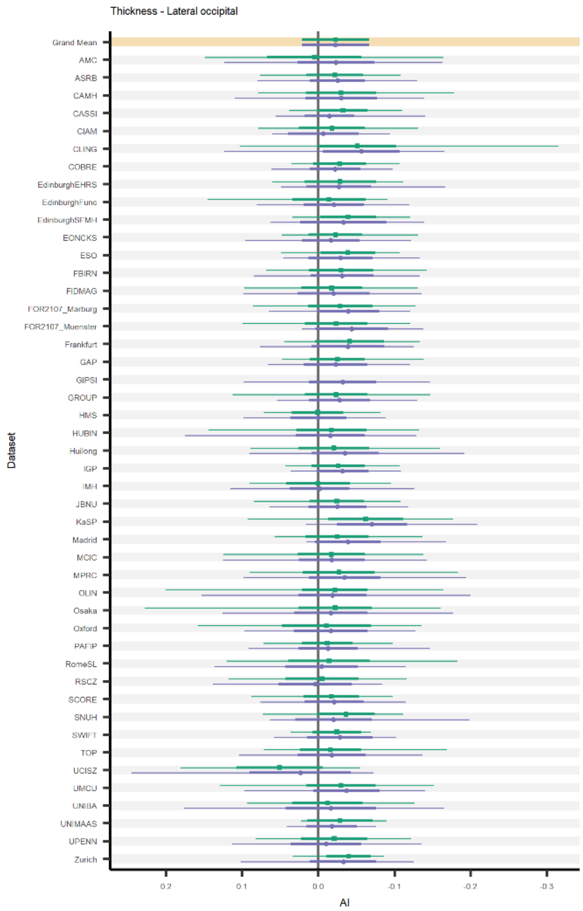
Thickness - Lateral occipital
Dataset
AI

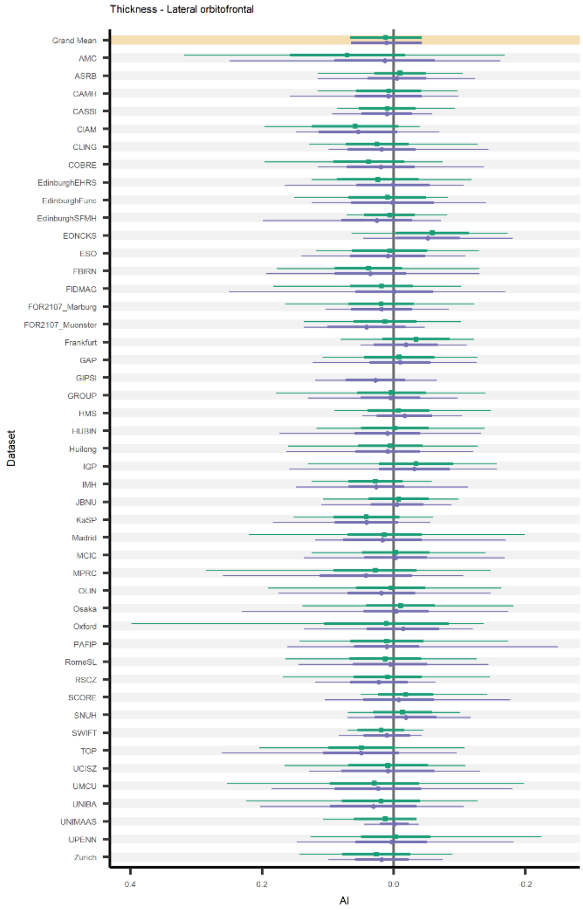
Thickness - Lateral orbitofrontal
Dataset
AI

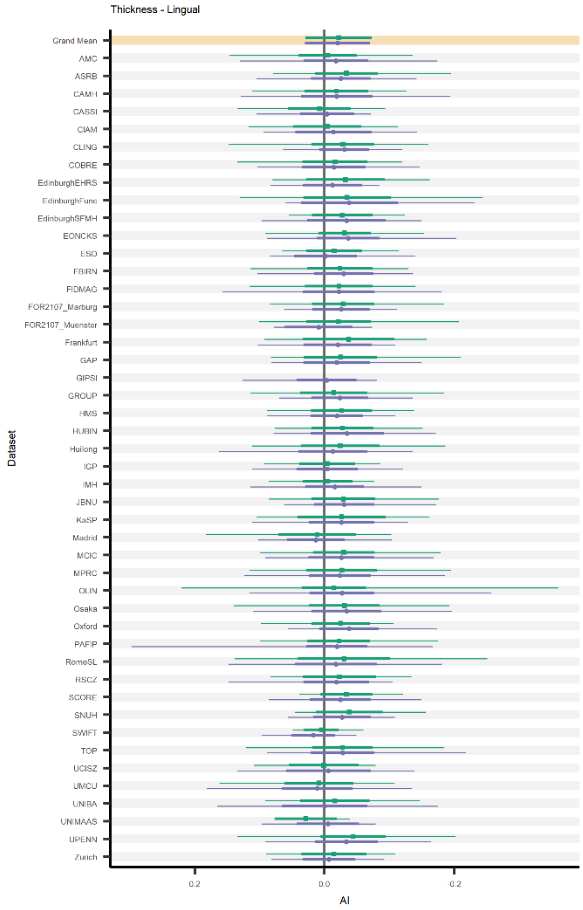
Thickness - Lingual
Dataset
AI

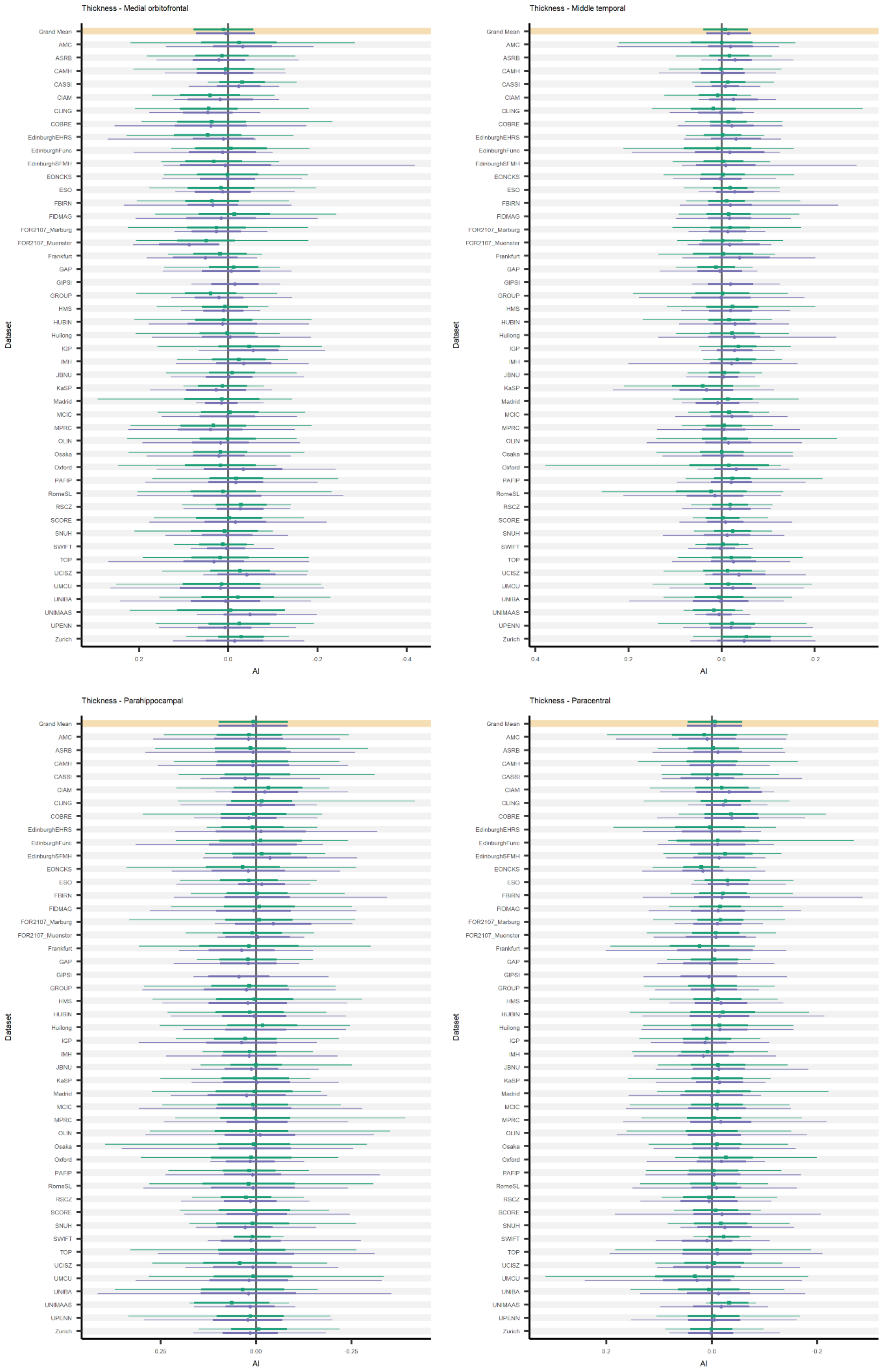
Thickness - Medial orbitofrontal
Thickness - Middle temporal
Thickness - Parahippocampal
Thickness - Paracentral
Dataset
AI

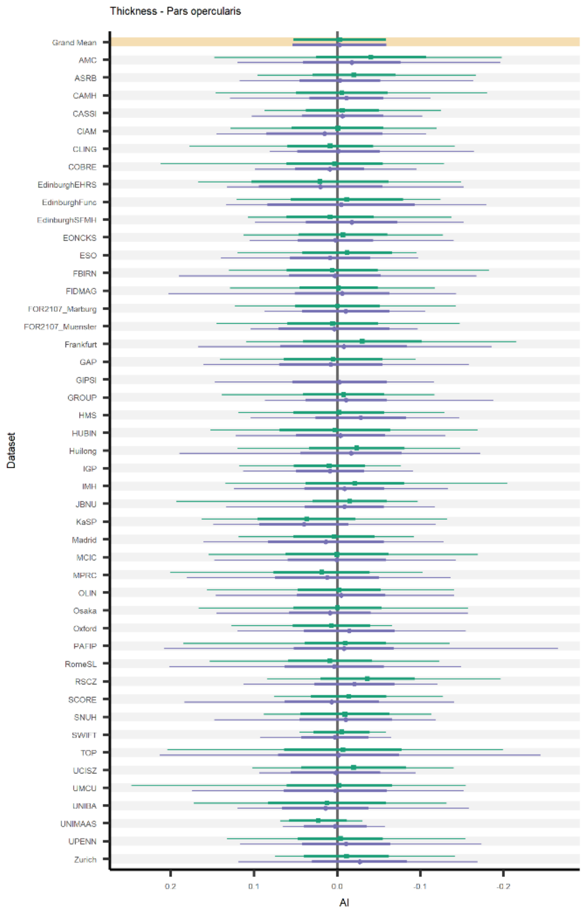
Thickness - Pars opercularis
Dataset
Grand Mean
AMC
ASRB
CAMH
CASSI
CIAM
CLING
COBRE
EdinburghEHRS
EdinburghFunc
EdinburghSFMH
EONCKS
ESO
FBIRN
FIDMAG
FOR2107_Marburg
FOR2107_Muenster
Frankfurt
GAP
GIPSI
GROUP
HMS
HUBIN
Huilong
IGP
IMH
JBNU
KaSP
Madrid
MCIC
MPRC
OLIN
Osaka
Oxford
PAFIP
RomeSL
RSCZ
SCORE
SNUH
SWIFT
TOP
UCISZ
UMCU
UNIBA
UNIMAAS
UPENN
Zurich
0.2
0.1
0.0
-0.1
-0.2
AI

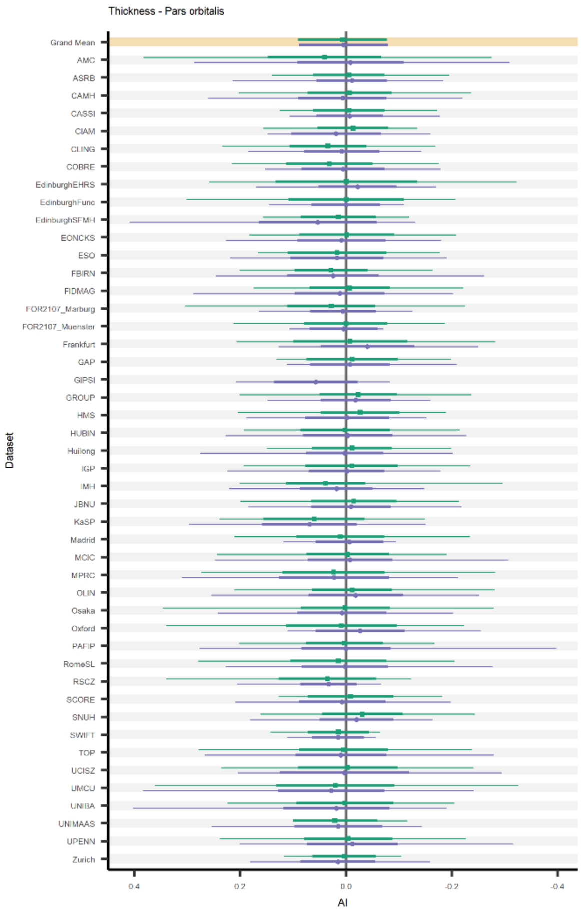
Thickness - Pars orbitalis
Dataset
Grand Mean
AMC
ASRB
CAMH
CASSI
CIAM
CLING
COBRE
EdinburghEHRS
EdinburghFunc
EdinburghSFMH
EONCKS
ESO
FBIRN
FIDMAG
FOR2107_Marburg
FOR2107_Muenster
Frankfurt
GAP
GIPSI
GROUP
HMS
HUBIN
Huilong
IGP
IMH
JBNU
KaSP
Madrid
MCIC
MPRC
OLIN
Osaka
Oxford
PAFIP
RomeSL
RSCZ
SCORE
SNUH
SWIFT
TOP
UCISZ
UMCU
UNIBA
UNIMAAS
UPENN
Zurich
0.4
0.2
0.0
-0.2
-0.4
AI

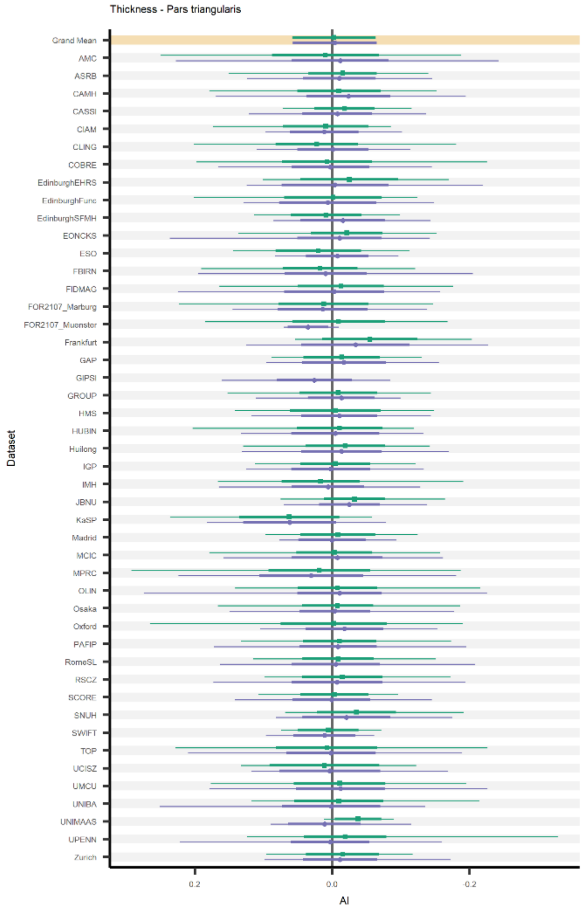
Thickness - Pars triangularis
Dataset
Grand Mean
AMC
ASRB
CAMH
CASSI
CIAM
CLING
COBRE
EdinburghEHRS
EdinburghFunc
EdinburghSFMH
EONCKS
ESO
FBIRN
FIDMAG
FOR2107_Marburg
FOR2107_Muenster
Frankfurt
GAP
GIPSI
GROUP
HMS
HUBIN
Huilong
IGP
IMH
JBNU
KaSP
Madrid
MCIC
MPRC
OLIN
Osaka
Oxford
PAFIP
RomeSL
RSCZ
SCORE
SNUH
SWIFT
TOP
UCISZ
UMCU
UNIBA
UNIMAAS
UPENN
Zurich
0.2
0.0
-0.2
AI

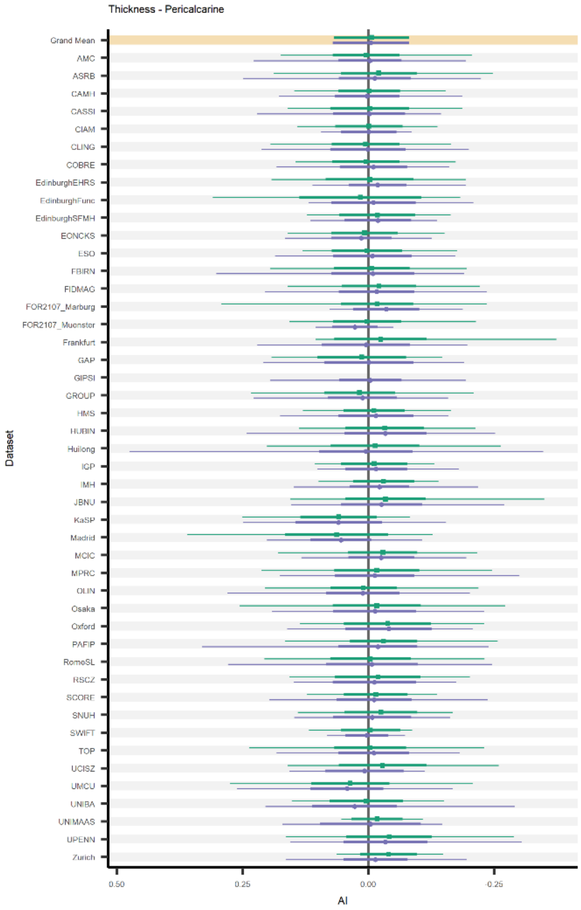
Thickness - Pericalcarine
Dataset
Grand Mean
AMC
ASRB
CAMH
CASSI
CIAM
CLING
COBRE
EdinburghEHRS
EdinburghFunc
EdinburghSFMH
EONCKS
ESO
FBIRN
FIDMAG
FOR2107_Marburg
FOR2107_Muenster
Frankfurt
GAP
GIPSI
GROUP
HMS
HUBIN
Huilong
IGP
IMH
JBNU
KaSP
Madrid
MCIC
MPRC
OLIN
Osaka
Oxford
PAFIP
RomeSL
RSCZ
SCORE
SNUH
SWIFT
TOP
UCISZ
UMCU
UNIBA
UNIMAAS
UPENN
Zurich
0.50
0.25
0.00
-0.25
AI

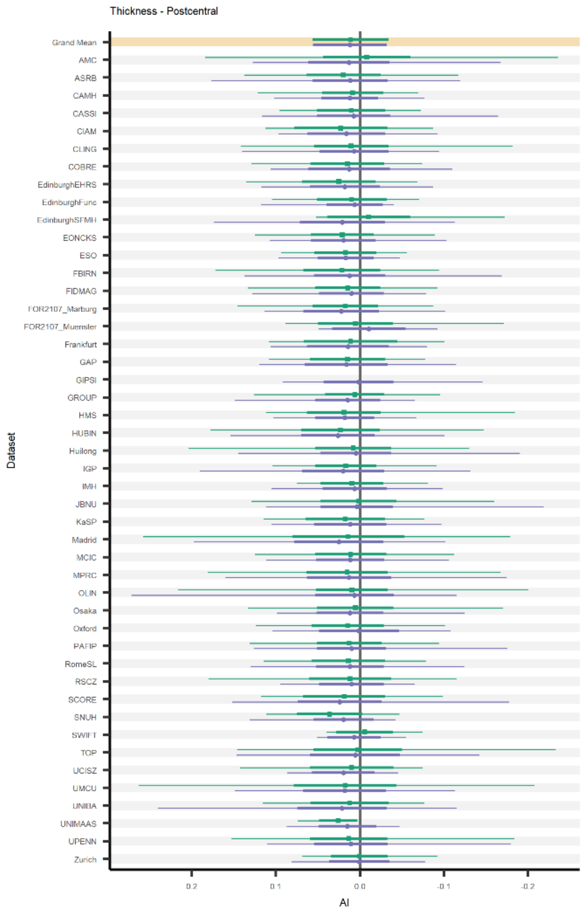
Thickness - Postcentral
Dataset
Grand Mean
AMC
ASRB
CAMH
CASSI
CIAM
CLING
COBRE
EdinburghEHRS
EdinburghFunc
EdinburghSFMH
EONCKS
ESO
FBIRN
FIDMAG
FOR2107_Marburg
FOR2107_Muenster
Frankfurt
GAP
GIPSI
GROUP
HMS
HUBIN
Huilong
IGP
IMH
JBNU
KaSP
Madrid
MCIC
MPRC
OLIN
Osaka
Oxford
PAFIP
RomeSL
RSCZ
SCORE
SNUH
SWIFT
TOP
UCISZ
UMCU
UNIBA
UNIMAAS
UPENN
Zurich
0.2
0.1
0.0
-0.1
-0.2
AI

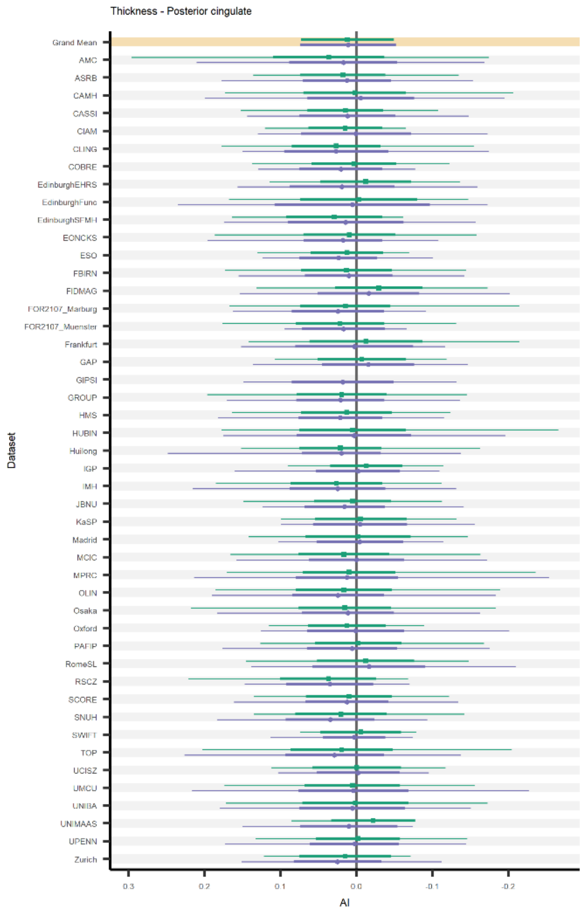
Thickness - Posterior cingulate
Dataset
Grand Mean
AMC
ASRB
CAMH
CASSI
CIAM
CLING
COBRE
EdinburghEHRS
EdinburghFunc
EdinburghSFMH
EONCKS
ESO
FBIRN
FIDMAG
FOR2107_Marburg
FOR2107_Muenster
Frankfurt
GAP
GIPSI
GROUP
HMS
HUBIN
Huilong
IGP
IMH
JBNU
KaSP
Madrid
MCIC
MPRC
OLIN
Osaka
Oxford
PAFIP
RomeSL
RSCZ
SCORE
SNUH
SWIFT
TOP
UCISZ
UMCU
UNIBA
UNIMAAS
UPENN
Zurich
0.3
0.2
0.1
0.0
-0.1
-0.2
AI

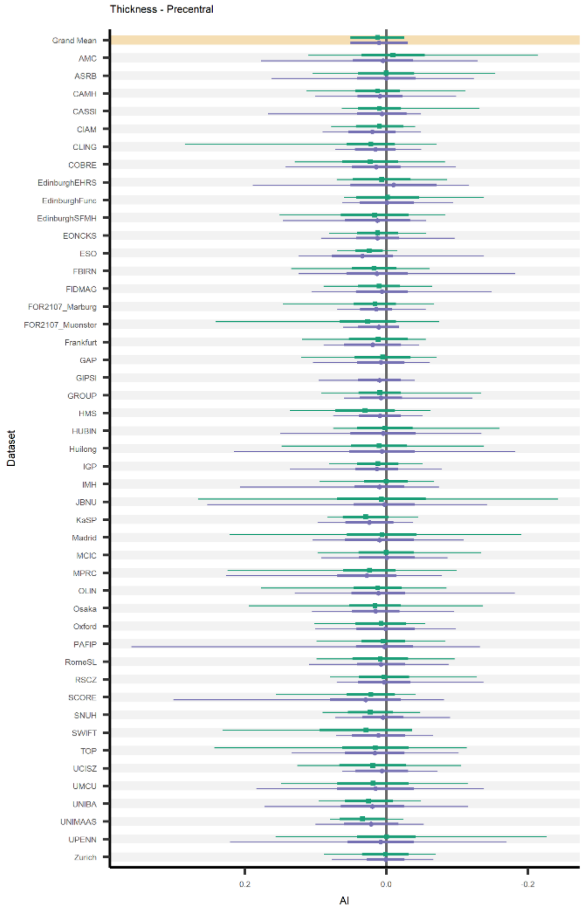
Thickness - Precentral
Dataset
Grand Mean
AMC
ASRB
CAMH
CASSI
CIAM
CLING
COBRE
EdinburghEHRS
EdinburghFunc
EdinburghSFMH
EONCKS
ESO
FBIRN
FIDMAG
FOR2107_Marburg
FOR2107_Muenster
Frankfurt
GAP
GIPSI
GROUP
HMS
HUBIN
Huilong
IGP
IMH
JBNU
KaSP
Madrid
MCIC
MPRC
OLIN
Osaka
Oxford
PAFIP
RomeSL
RSCZ
SCORE
SNUH
SWIFT
TOP
UCISZ
UMCU
UNIBA
UNIMAAS
UPENN
Zurich
0.2
0.0
-0.2
AI

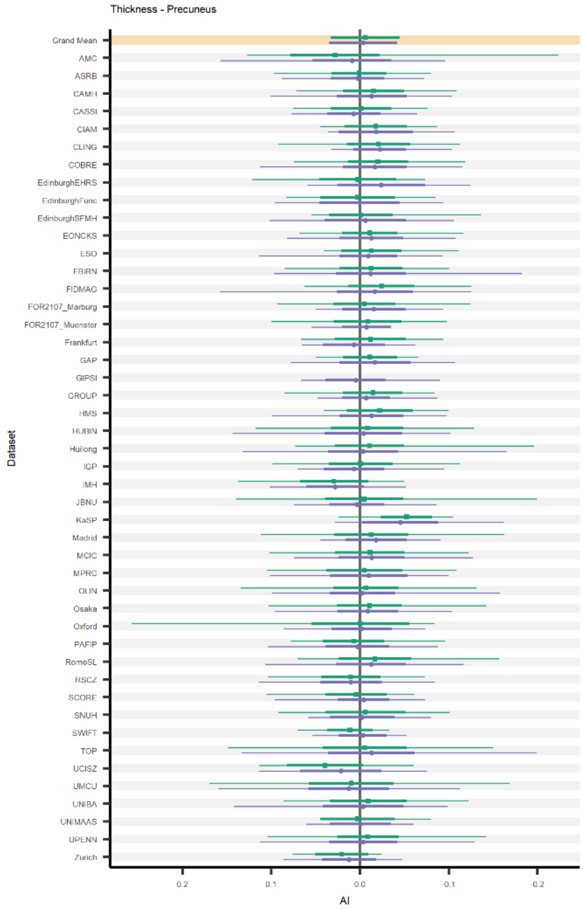
Thickness - Precuneus
Dataset
Grand Mean
AMC
ASRB
CAMH
CASSI
CIAM
CLING
COBRE
EdinburghEHRS
EdinburghFunc
EdinburghSFMH
EONCKS
ESO
FBIRN
FIDMAG
FOR2107_Marburg
FOR2107_Muenster
Frankfurt
GAP
GIPSI
GROUP
HMS
HUBIN
Huilong
IGP
IMH
JBNU
KaSP
Madrid
MCIC
MPRC
OLIN
Osaka
Oxford
PAFIP
RomeSL
RSCZ
SCORE
SNUH
SWIFT
TOP
UCISZ
UMCU
UNIBA
UNIMAAS
UPENN
Zurich
0.2
0.1
0.0
-0.1
-0.2
AI

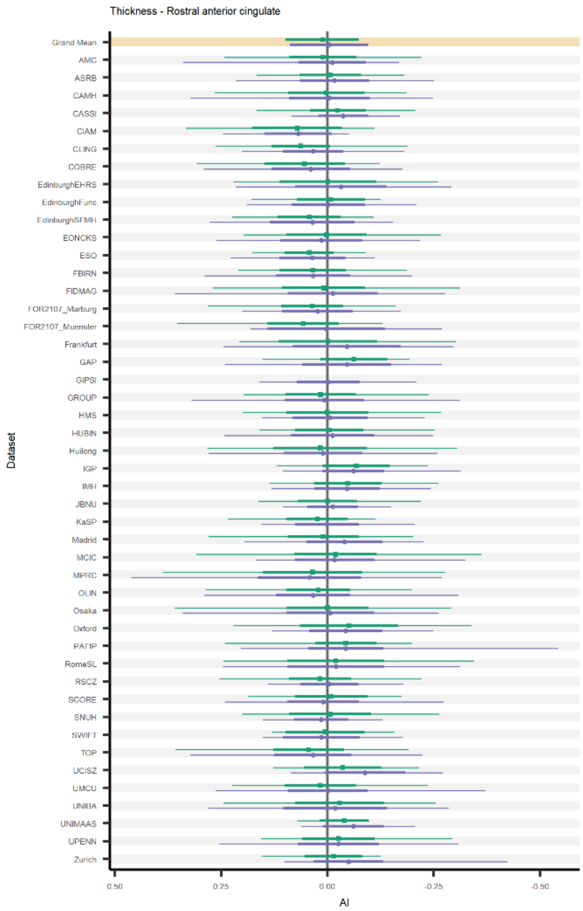
Thickness - Rostral anterior cingulate
Grand Mean
AMC
ASRB
CAMH
CASSI
CIAM
CLING
COBRE
EdinburghEHRS
EdinburghFunc
EdinburghSFMH
EONCKS
ESO
FBIRN
FIDMAG
FOR2107_Marburg
FOR2107_Muenster
Frankfurt
GAP
GIPSI
GROUP
HMS
HUBIN
Huilong
IGP
IMH
JBNU
KaSP
Madrid
MCIC
MPRC
OLIN
Osaka
Oxford
PAFIP
RomeSL
RSCZ
SCORE
SNUH
SWIFT
TOP
UCISZ
UMCU
UNIBA
UNIMAAS
UPENN
Zurich
Dataset
0.50
0.25
0.00
-0.25
-0.50
AI

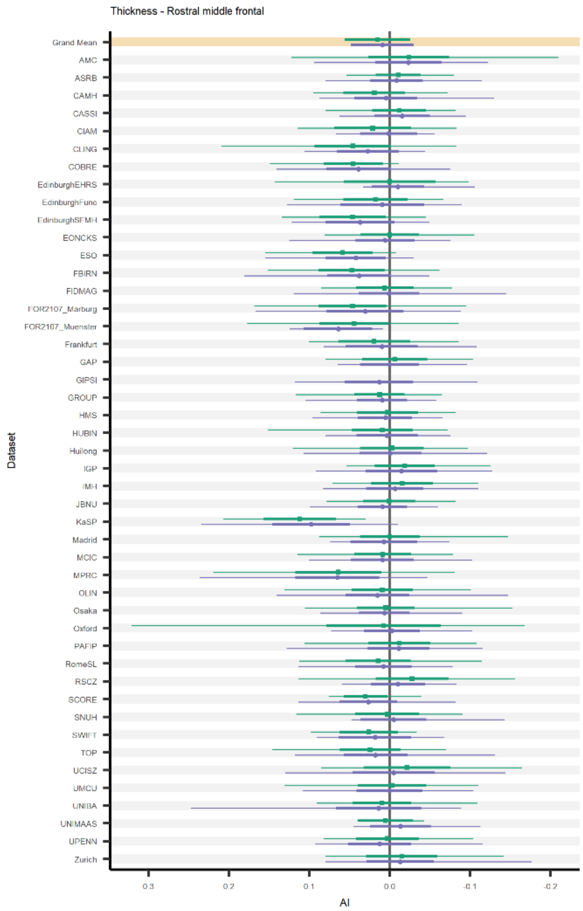
Thickness - Rostral middle frontal
Grand Mean
AMC
ASRB
CAMH
CASSI
CIAM
CLING
COBRE
EdinburghEHRS
EdinburghFunc
EdinburghSFMH
EONCKS
ESO
FBIRN
FIDMAG
FOR2107_Marburg
FOR2107_Muenster
Frankfurt
GAP
GIPSI
GROUP
HMS
HUBIN
Huilong
IGP
IMH
JBNU
KaSP
Madrid
MCIC
MPRC
OLIN
Osaka
Oxford
PAFIP
RomeSL
RSCZ
SCORE
SNUH
SWIFT
TOP
UCISZ
UMCU
UNIBA
UNIMAAS
UPENN
Zurich
Dataset
0.3
0.2
0.1
0.0
-0.1
-0.2
AI

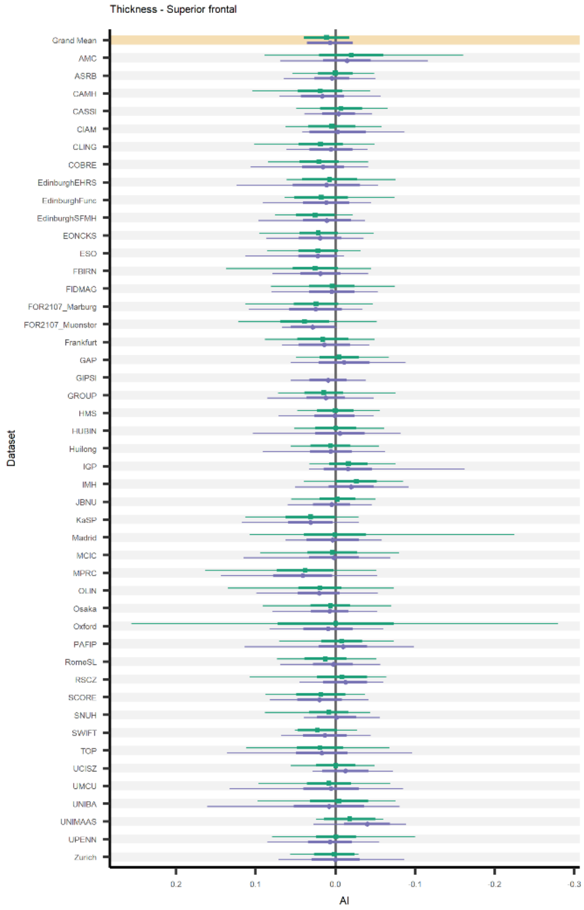
Thickness - Superior frontal
Grand Mean
AMC
ASRB
CAMH
CASSI
CIAM
CLING
COBRE
EdinburghEHRS
EdinburghFunc
EdinburghSFMH
EONCKS
ESO
FBIRN
FIDMAG
FOR2107_Marburg
FOR2107_Muenster
Frankfurt
GAP
GIPSI
GROUP
HMS
HUBIN
Huilong
IGP
IMH
JBNU
KaSP
Madrid
MCIC
MPRC
OLIN
Osaka
Oxford
PAFIP
RomeSL
RSCZ
SCORE
SNUH
SWIFT
TOP
UCISZ
UMCU
UNIBA
UNIMAAS
UPENN
Zurich
Dataset
0.2
0.1
0.0
-0.1
-0.2
-0.3
AI

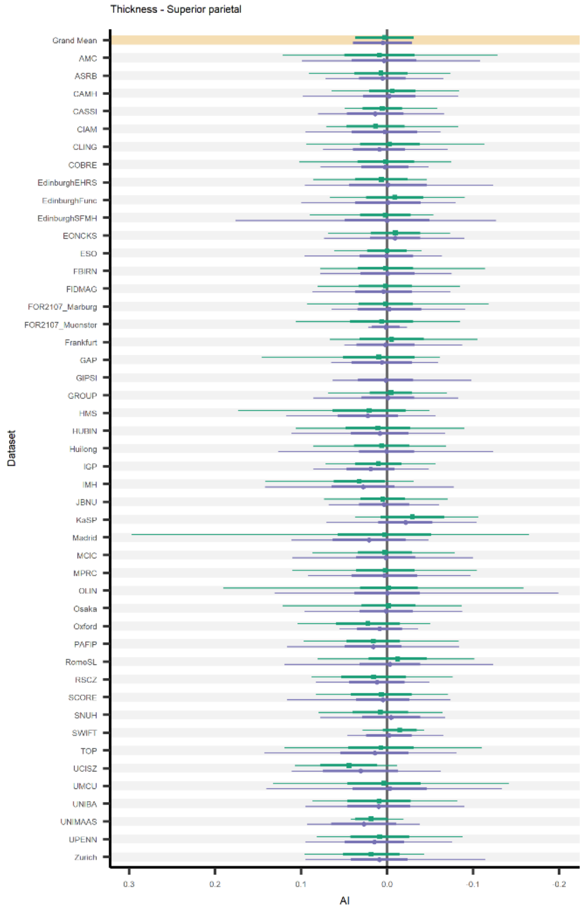
Thickness - Superior parietal
Grand Mean
AMC
ASRB
CAMH
CASSI
CIAM
CLING
COBRE
EdinburghEHRS
EdinburghFunc
EdinburghSFMH
EONCKS
ESO
FBIRN
FIDMAG
FOR2107_Marburg
FOR2107_Muenster
Frankfurt
GAP
GIPSI
GROUP
HMS
HUBIN
Huilong
IGP
IMH
JBNU
KaSP
Madrid
MCIC
MPRC
OLIN
Osaka
Oxford
PAFIP
RomeSL
RSCZ
SCORE
SNUH
SWIFT
TOP
UCISZ
UMCU
UNIBA
UNIMAAS
UPENN
Zurich
Dataset
0.3
0.2
0.1
0.0
-0.1
-0.2
AI

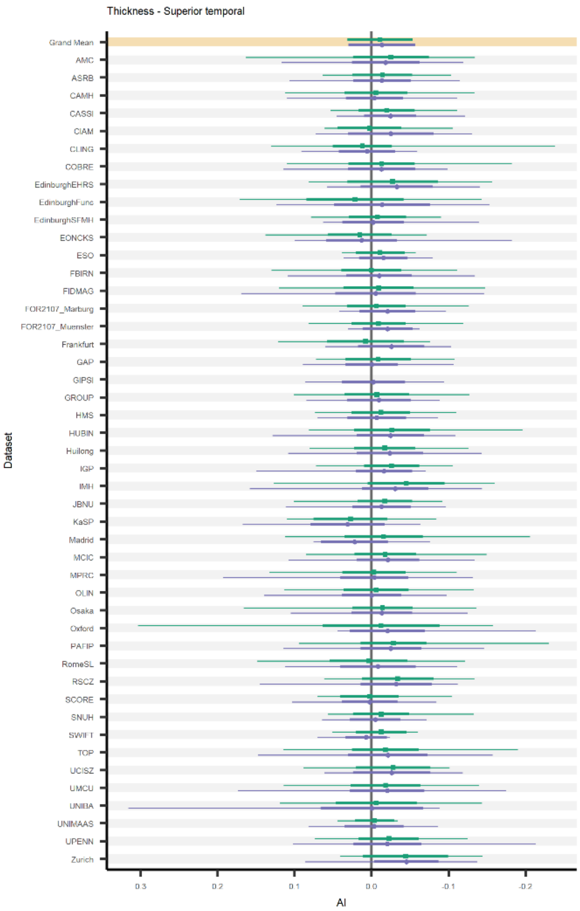
Thickness - Superior temporal
Grand Mean
AMC
ASRB
CAMH
CASSI
CIAM
CLING
COBRE
EdinburghEHRS
EdinburghFunc
EdinburghSFMH
EONCKS
ESO
FBIRN
FIDMAG
FOR2107_Marburg
FOR2107_Muenster
Frankfurt
GAP
GIPSI
GROUP
HMS
HUBIN
Huilong
IGP
IMH
JBNU
KaSP
Madrid
MCIC
MPRC
OLIN
Osaka
Oxford
PAFIP
RomeSL
RSCZ
SCORE
SNUH
SWIFT
TOP
UCISZ
UMCU
UNIBA
UNIMAAS
UPENN
Zurich
Dataset
0.3
0.2
0.1
0.0
-0.1
-0.2
AI

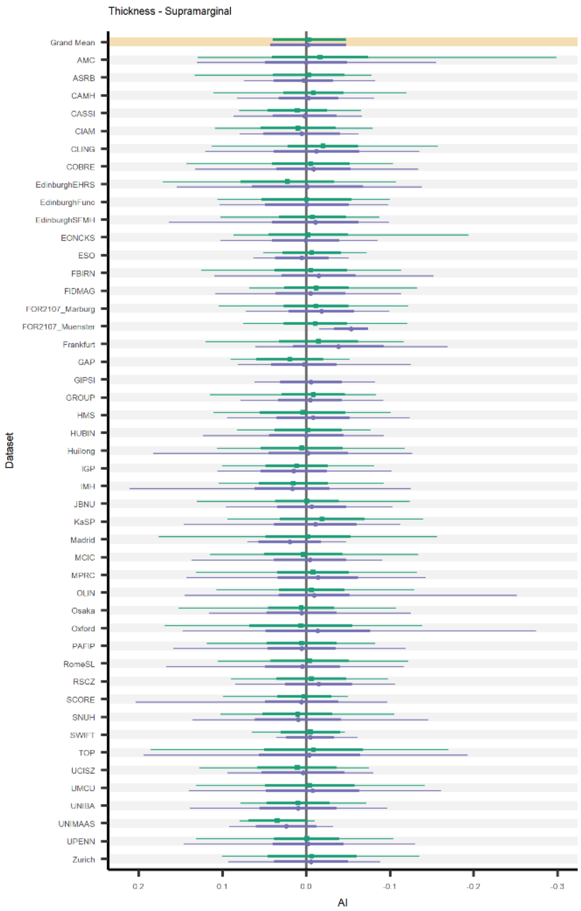
Thickness - Supramarginal
Grand Mean
AMC
ASRB
CAMH
CASSI
CIAM
CLING
COBRE
EdinburghEHRS
EdinburghFunc
EdinburghSFMH
EONCKS
ESO
FBIRN
FIDMAG
FOR2107_Marburg
FOR2107_Muenster
Frankfurt
GAP
GIPSI
GROUP
HMS
HUBIN
Huilong
IGP
IMH
JBNU
KaSP
Madrid
MCIC
MPRC
OLIN
Osaka
Oxford
PAFIP
RomeSL
RSCZ
SCORE
SNUH
SWIFT
TOP
UCISZ
UMCU
UNIBA
UNIMAAS
UPENN
Zurich
Dataset
0.2
0.1
0.0
-0.1
-0.2
-0.3
AI

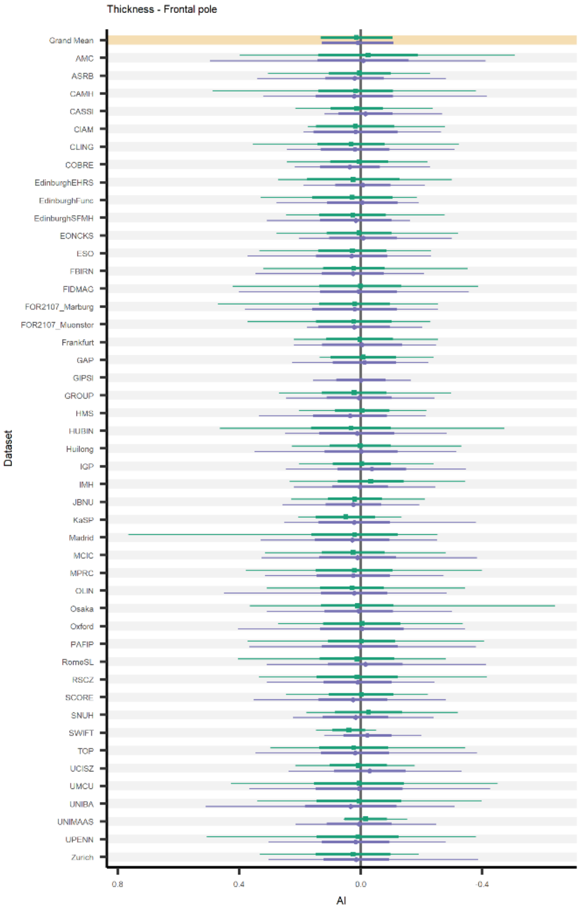
Thickness - Frontal pole
Grand Mean
AMC
ASRB
CAMH
CASSI
CIAM
CLING
COBRE
EdinburghEHRS
EdinburghFunc
EdinburghSFMH
EONCKS
ESO
FBIRN
FIDMAG
FOR2107_Marburg
FOR2107_Muenster
Frankfurt
GAP
GIPSI
GROUP
HMS
HUBIN
Huilong
IGP
IMH
JBNU
KaSP
Madrid
MCIC
MPRC
OLIN
Osaka
Oxford
PAFIP
RomeSL
RSCZ
SCORE
SNUH
SWIFT
TOP
UCISZ
UMCU
UNIBA
UNIMAAS
UPENN
Zurich
Dataset
0.8
0.4
0.0
-0.4
AI

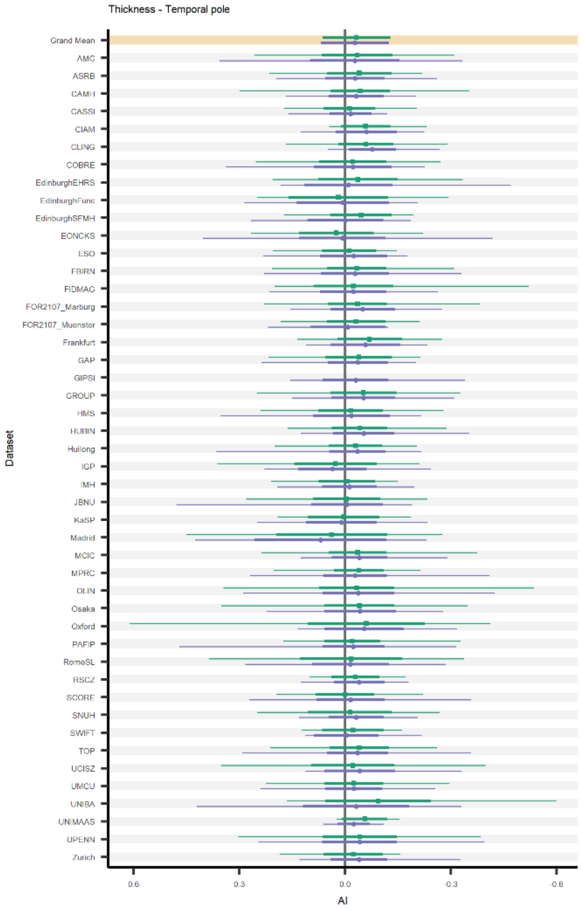
Thickness - Temporal pole
Grand Mean
AMC
ASRB
CAMH
CASSI
CIAM
CLING
COBRE
EdinburghEHRS
EdinburghFunc
EdinburghSFMH
EONCKS
ESO
FBIRN
FIDMAG
FOR2107_Marburg
FOR2107_Muenster
Frankfurt
GAP
GIPSI
GROUP
HMS
HUBIN
Huilong
IGP
IMH
JBNU
KaSP
Madrid
MCIC
MPRC
OLIN
Osaka
Oxford
PAFIP
RomeSL
RSCZ
SCORE
SNUH
SWIFT
TOP
UCISZ
UMCU
UNIBA
UNIMAAS
UPENN
Zurich
Dataset
0.6
0.3
0.0
-0.3
-0.6
AI

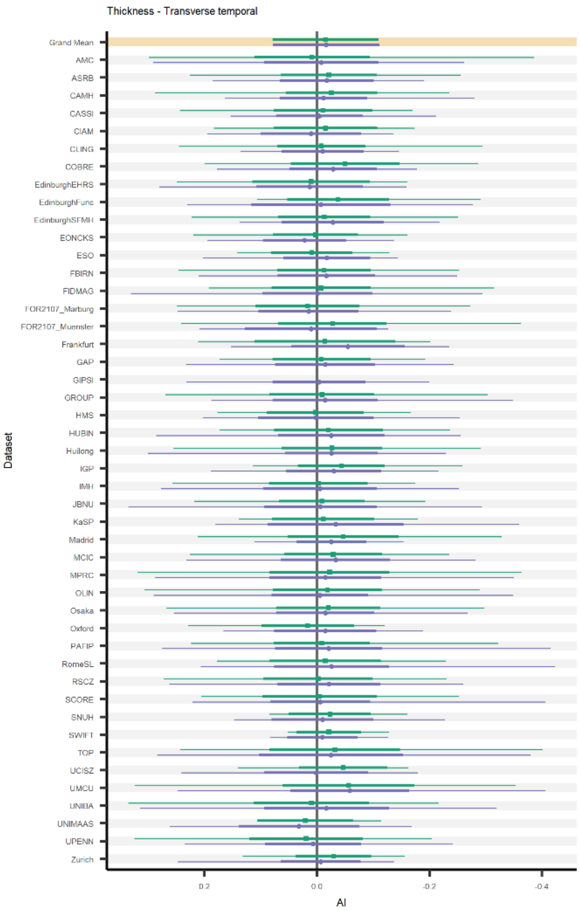
Thickness - Transverse temporal
Grand Mean
AMC
ASRB
CAMH
CASSI
CIAM
CLING
COBRE
EdinburghEHRS
EdinburghFunc
EdinburghSFMH
EONCKS
ESO
FBIRN
FIDMAG
FOR2107_Marburg
FOR2107_Muenster
Frankfurt
GAP
GIPSI
GROUP
HMS
HUBIN
Huilong
IGP
IMH
JBNU
KaSP
Madrid
MCIC
MPRC
OLIN
Osaka
Oxford
PAFIP
RomeSL
RSCZ
SCORE
SNUH
SWIFT
TOP
UCISZ
UMCU
UNIBA
UNIMAAS
UPENN
Zurich
Dataset
AI

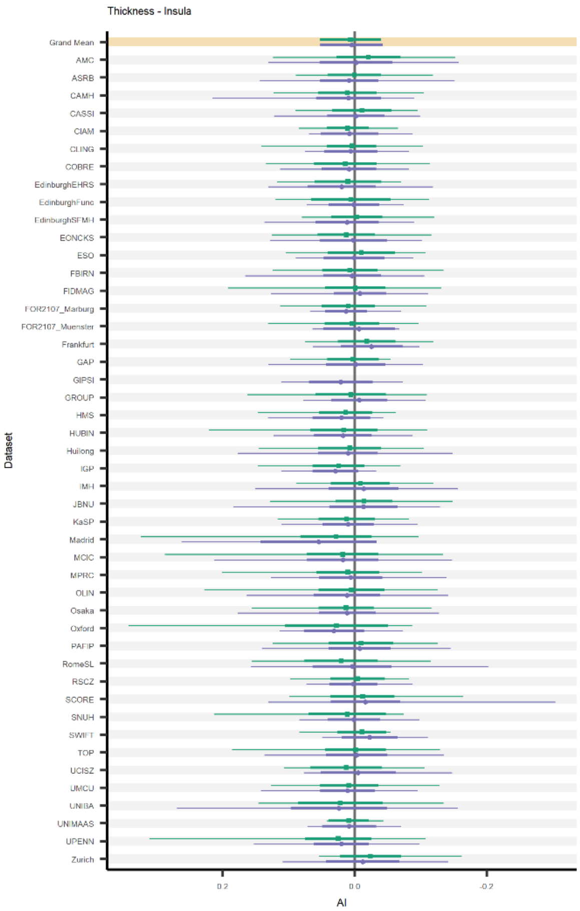
Thickness - Insula
Grand Mean
AMC
ASRB
CAMH
CASSI
CIAM
CLING
COBRE
EdinburghEHRS
EdinburghFunc
EdinburghSFMH
EONCKS
ESO
FBIRN
FIDMAG
FOR2107_Marburg
FOR2107_Muenster
Frankfurt
GAP
GIPSI
GROUP
HMS
HUBIN
Huilong
IGP
IMH
JBNU
KaSP
Madrid
MCIC
MPRC
OLIN
Osaka
Oxford
PAFIP
RomeSL
RSCZ
SCORE
SNUH
SWIFT
TOP
UCISZ
UMCU
UNIBA
UNIMAAS
UPENN
Zurich
Dataset
AI

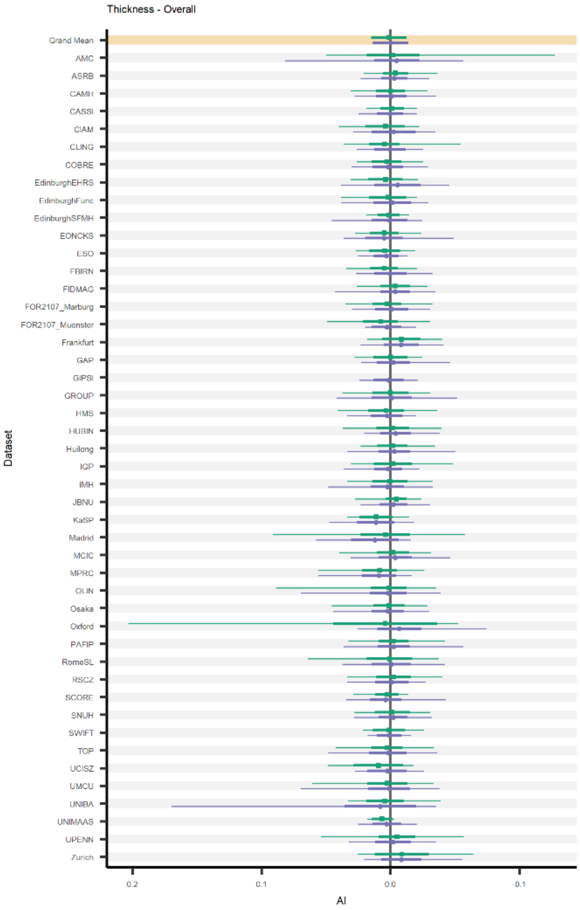
Thickness - Overall
Grand Mean
AMC
ASRB
CAMH
CASSI
CIAM
CLING
COBRE
EdinburghEHRS
EdinburghFunc
EdinburghSFMH
EONCKS
ESO
FBIRN
FIDMAG
FOR2107_Marburg
FOR2107_Muenster
Frankfurt
GAP
GIPSI
GROUP
HMS
HUBIN
Huilong
IGP
IMH
JBNU
KaSP
Madrid
MCIC
MPRC
OLIN
Osaka
Oxford
PAFIP
RomeSL
RSCZ
SCORE
SNUH
SWIFT
TOP
UCISZ
UMCU
UNIBA
UNIMAAS
UPENN
Zurich
Dataset
AI

**Fig. S2 (page 16-24). Overall and per-dataset average and range for cortical surface area asymmetries.** For each cortical surface area asymmetry measure, the average in controls (green circles) and individuals affected with schizophrenia (purple squares) is shown. The top (highlighted) row contains the grand sample size-weighted mean and standard deviation (thick line segments). The other rows contain per-dataset averages, standard deviations and minimum and maximum values (indicated with thin line segments).

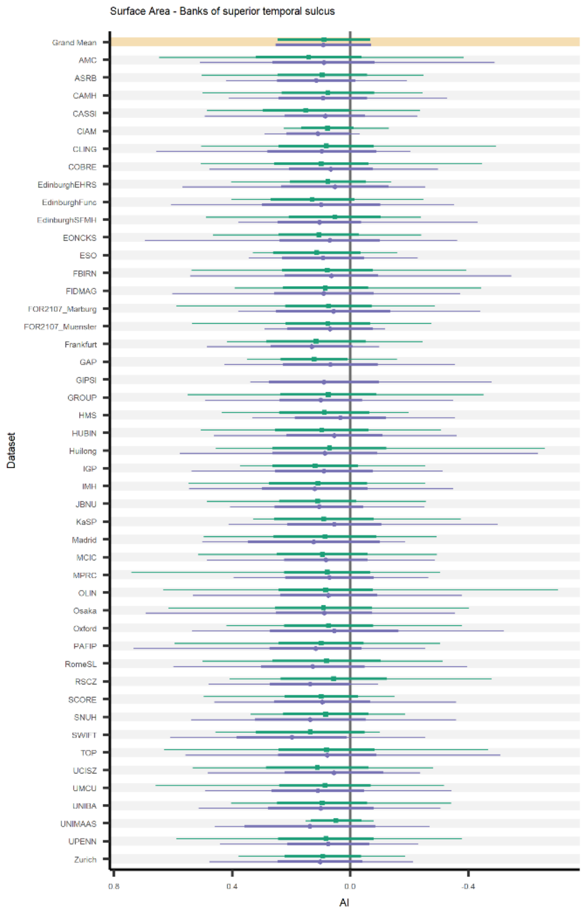
Surface Area - Banks of superior temporal sulcus
Dataset
Grand Mean
AMC
ASRB
CAMH
CASSI
CIAM
CLING
COBRE
EdinburghEHRS
EdinburghFunc
EdinburghSFMH
EONCKS
ESO
FBIRN
FIDMAG
FOR2107_Marburg
FOR2107_Muenster
Frankfurt
GAP
GIPSI
GROUP
HMS
HUBIN
Huilong
IGP
IMH
JBNU
KaSP
Madrid
MCIC
MPRC
OLIN
Osaka
Oxford
PAFIP
RomeSL
RSCZ
SCORE
SNUH
SWIFT
TOP
UCISZ
UMCU
UNIBA
UNIMAAS
UPENN
Zurich
0.8
0.4
0.0
-0.4
AI

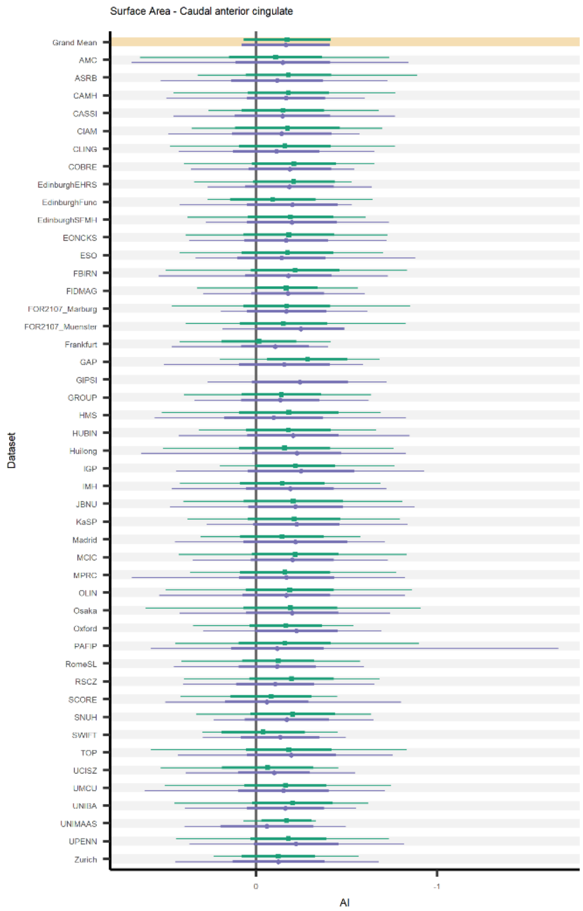
Surface Area - Caudal anterior cingulate
Dataset
Grand Mean
AMC
ASRB
CAMH
CASSI
CIAM
CLING
COBRE
EdinburghEHRS
EdinburghFunc
EdinburghSFMH
EONCKS
ESO
FBIRN
FIDMAG
FOR2107_Marburg
FOR2107_Muenster
Frankfurt
GAP
GIPSI
GROUP
HMS
HUBIN
Huilong
IGP
IMH
JBNU
KaSP
Madrid
MCIC
MPRC
OLIN
Osaka
Oxford
PAFIP
RomeSL
RSCZ
SCORE
SNUH
SWIFT
TOP
UCISZ
UMCU
UNIBA
UNIMAAS
UPENN
Zurich
0
-1
AI

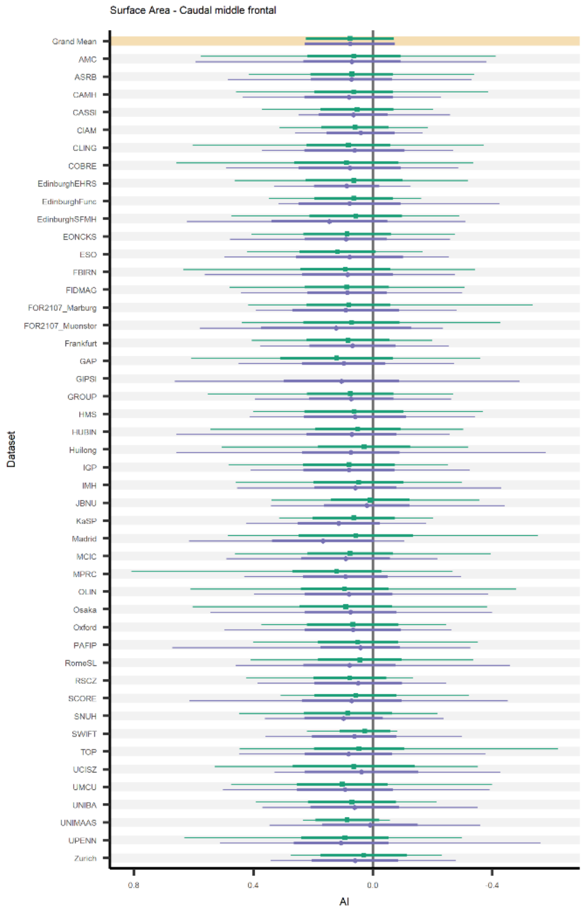
Surface Area - Caudal middle frontal
Dataset
Grand Mean
AMC
ASRB
CAMH
CASSI
CIAM
CLING
COBRE
EdinburghEHRS
EdinburghFunc
EdinburghSFMH
EONCKS
ESO
FBIRN
FIDMAG
FOR2107_Marburg
FOR2107_Muenster
Frankfurt
GAP
GIPSI
GROUP
HMS
HUBIN
Huilong
IGP
IMH
JBNU
KaSP
Madrid
MCIC
MPRC
OLIN
Osaka
Oxford
PAFIP
RomeSL
RSCZ
SCORE
SNUH
SWIFT
TOP
UCISZ
UMCU
UNIBA
UNIMAAS
UPENN
Zurich
0.8
0.4
0.0
-0.4
AI

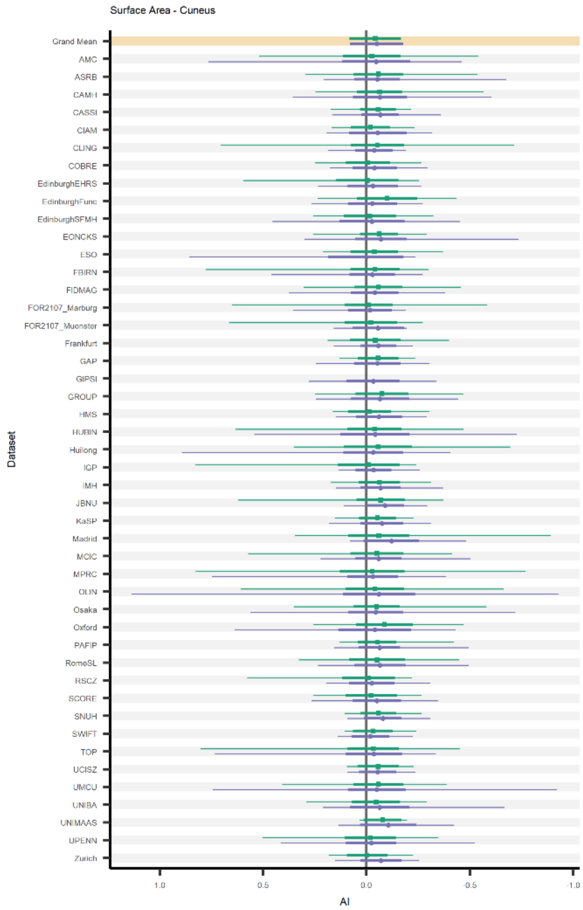
Surface Area - Cuneus
Dataset
Grand Mean
AMC
ASRB
CAMH
CASSI
CIAM
CLING
COBRE
EdinburghEHRS
EdinburghFunc
EdinburghSFMH
EONCKS
ESO
FBIRN
FIDMAG
FOR2107_Marburg
FOR2107_Muenster
Frankfurt
GAP
GIPSI
GROUP
HMS
HUBIN
Huilong
IGP
IMH
JBNU
KaSP
Madrid
MCIC
MPRC
OLIN
Osaka
Oxford
PAFIP
RomeSL
RSCZ
SCORE
SNUH
SWIFT
TOP
UCISZ
UMCU
UNIBA
UNIMAAS
UPENN
Zurich
1.0
0.5
0.0
-0.5
-1.0
AI

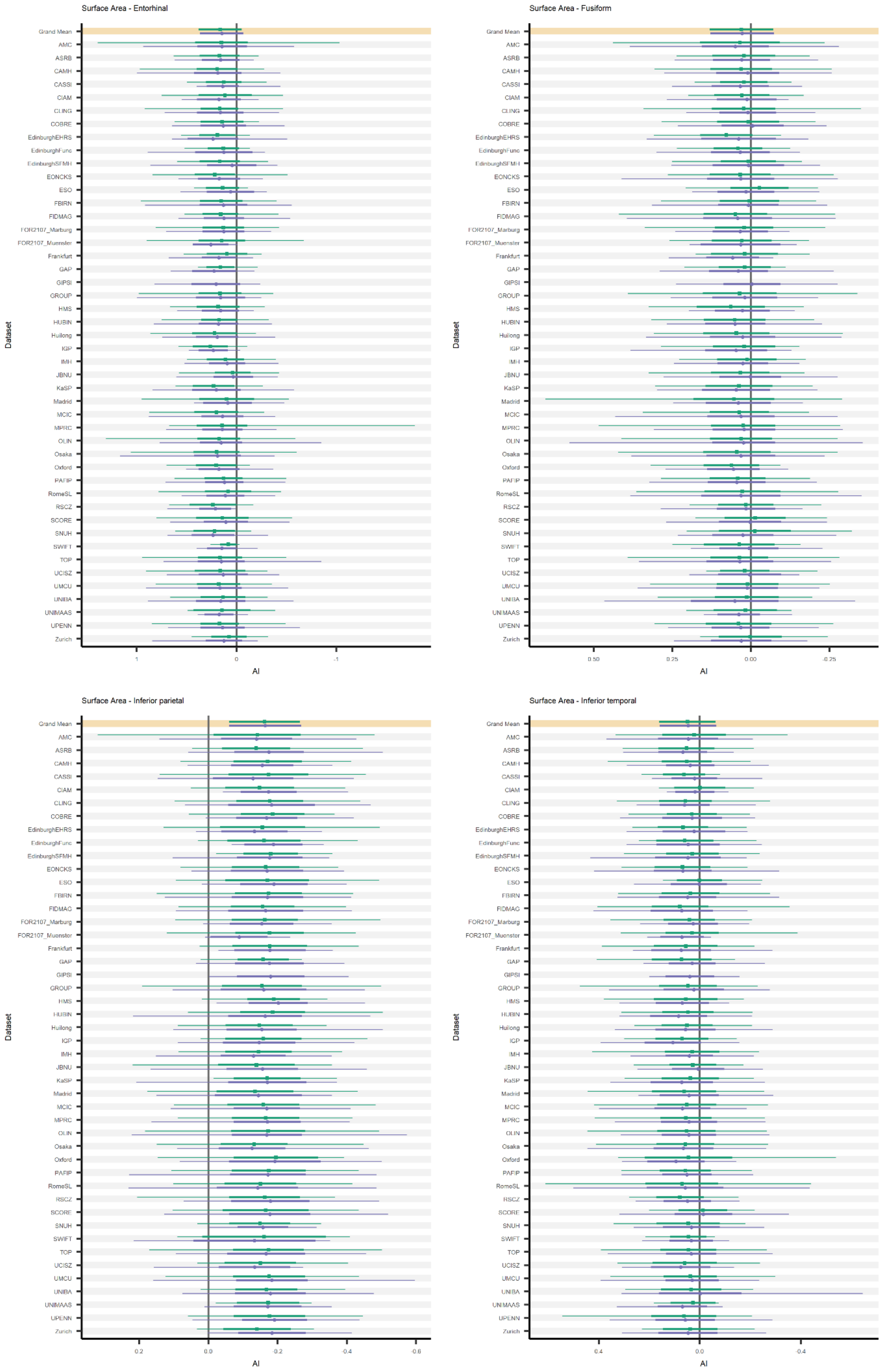
Surface Area - Entorhinal
Surface Area - Fusiform
Surface Area - Inferior parietal
Surface Area - Inferior temporal
Grand Mean
AMC
ASRB
CAMH
CASSI
CIAM
CLING
COBRE
EdinburghEHRS
EdinburghFunc
EdinburghSFMH
EONCKS
ESO
FBIRN
FIDMAG
FOR2107_Marburg
FOR2107_Muenster
Frankfurt
GAP
GIPSI
GROUP
HMS
HUBIN
Huilong
IGP
IMH
JBNU
KaSP
Madrid
MCIC
MPRC
OLIN
Osaka
Oxford
PAFIP
RomeSL
RSCZ
SCORE
SNUH
SWIFT
TOP
UCISZ
UMCU
UNIBA
UNIMAAS
UPENN
Zurich
Dataset
AI

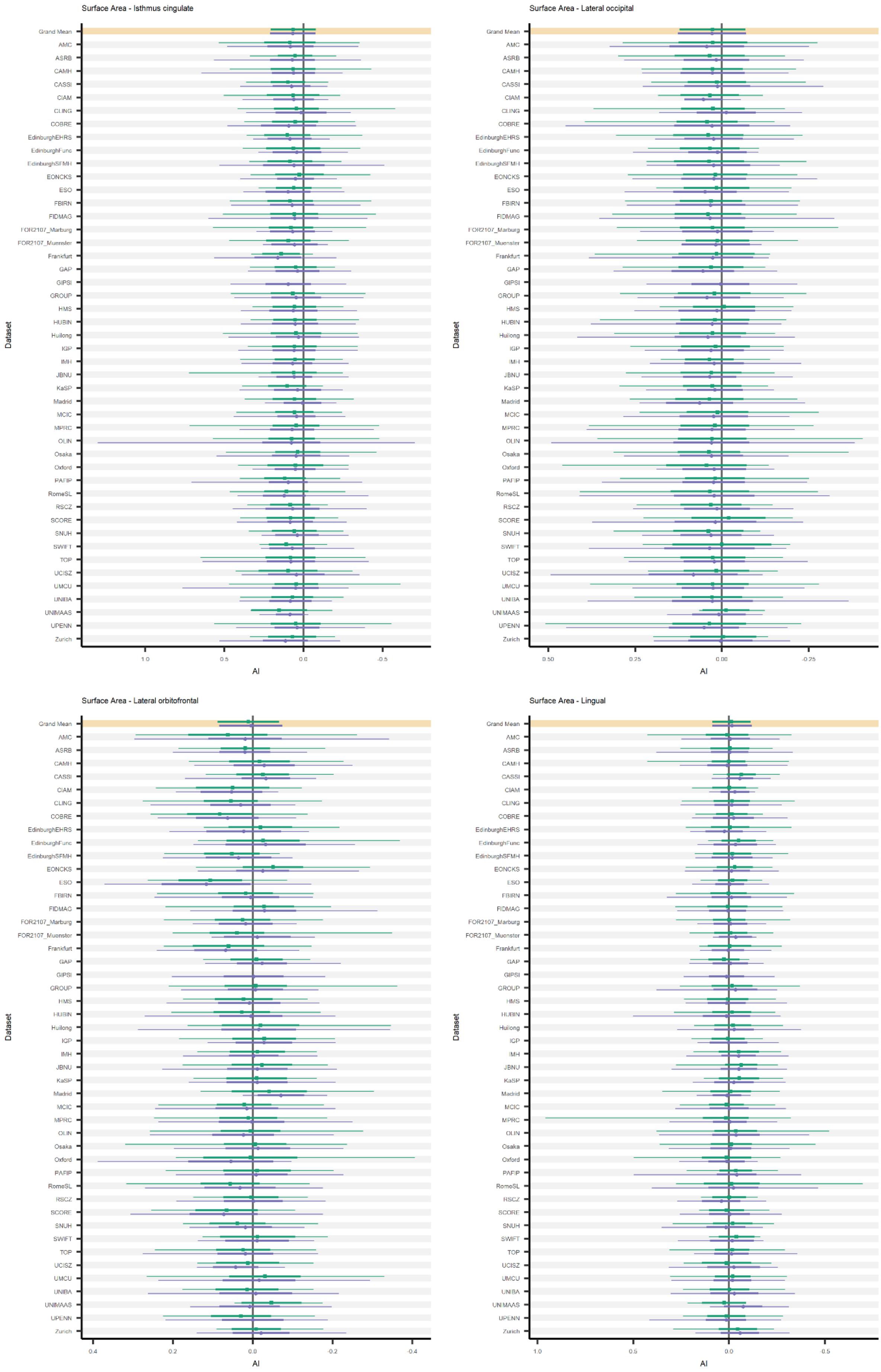
Surface Area - Isthmus cingulate
Surface Area - Lateral occipital
Surface Area - Lateral orbitofrontal
Surface Area - Lingual
Dataset
Grand Mean
AMC
ASRB
CAMH
CASSI
CIAM
CLING
COBRE
EdinburghEHRS
EdinburghFunc
EdinburghSFMH
EONCKS
ESO
FBIRN
FIDMAG
FOR2107_Marburg
FOR2107_Muenster
Frankfurt
GAP
GIPSI
GROUP
HMS
HUBIN
Huilong
IGP
IMH
JBNU
KaSP
Madrid
MCIC
MPRC
OLIN
Osaka
Oxford
PAFIP
RomeSL
RSCZ
SCORE
SNUH
SWIFT
TOP
UCISZ
UMCU
UNIBA
UNIMAAS
UPENN
Zurich
1.0
0.5
0.0
-0.5
0.50
0.25
0.00
-0.25
0.4
0.2
0.0
-0.2
-0.4
AI

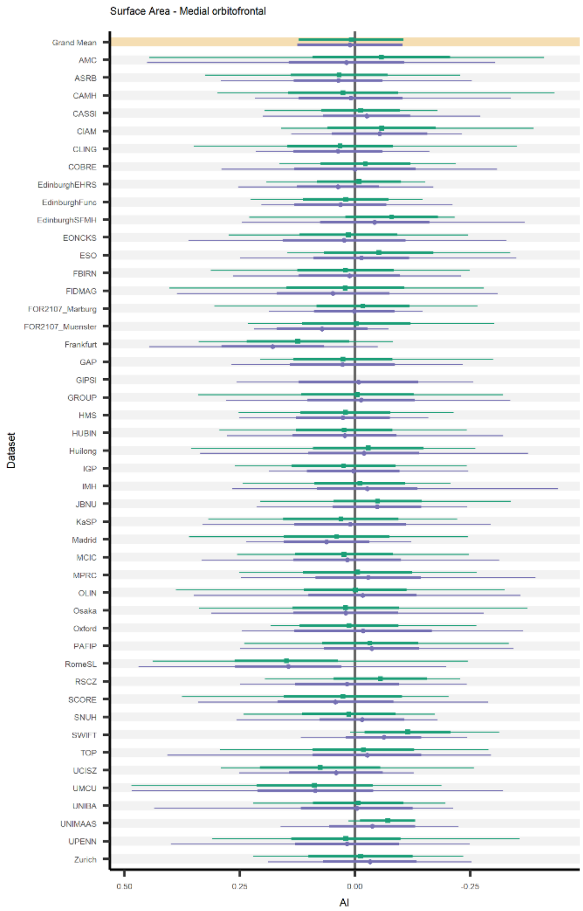
Surface Area - Medial orbitofrontal
Dataset
Grand Mean
AMC
ASRB
CAMH
CASSI
CIAM
CLING
COBRE
EdinburghEHRS
EdinburghFunc
EdinburghSFMH
EONCKS
ESO
FBIRN
FIDMAG
FOR2107_Marburg
FOR2107_Muenster
Frankfurt
GAP
GIPSI
GROUP
HMS
HUBIN
Huilong
IGP
IMH
JBNU
KaSP
Madrid
MCIC
MPRC
OLIN
Osaka
Oxford
PAFIP
RomeSL
RSCZ
SCORE
SNUH
SWIFT
TOP
UCISZ
UMCU
UNIBA
UNIMAAS
UPENN
Zurich
0.50
0.25
0.00
-0.25
AI

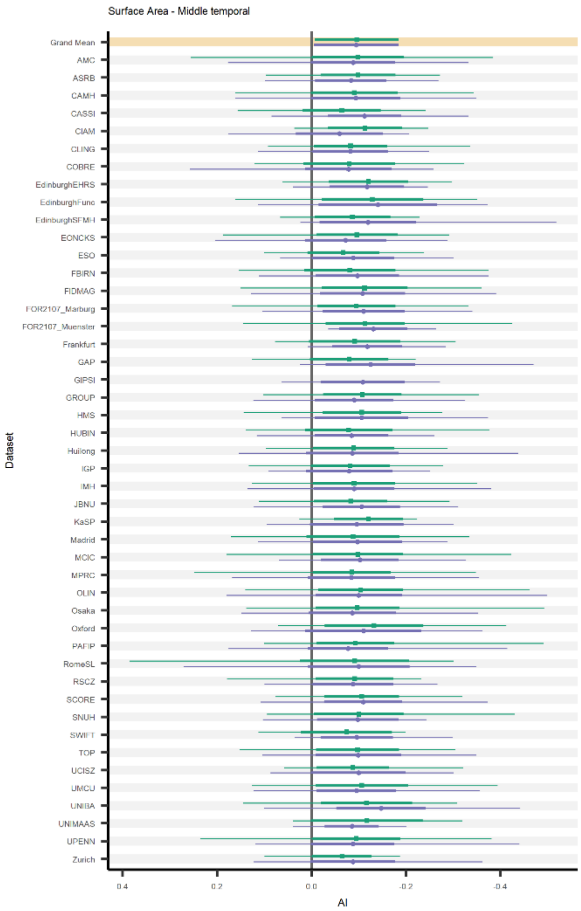
Surface Area - Middle temporal
Dataset
Grand Mean
AMC
ASRB
CAMH
CASSI
CIAM
CLING
COBRE
EdinburghEHRS
EdinburghFunc
EdinburghSFMH
EONCKS
ESO
FBIRN
FIDMAG
FOR2107_Marburg
FOR2107_Muenster
Frankfurt
GAP
GIPSI
GROUP
HMS
HUBIN
Huilong
IGP
IMH
JBNU
KaSP
Madrid
MCIC
MPRC
OLIN
Osaka
Oxford
PAFIP
RomeSL
RSCZ
SCORE
SNUH
SWIFT
TOP
UCISZ
UMCU
UNIBA
UNIMAAS
UPENN
Zurich
0.4
0.2
0.0
-0.2
-0.4
AI

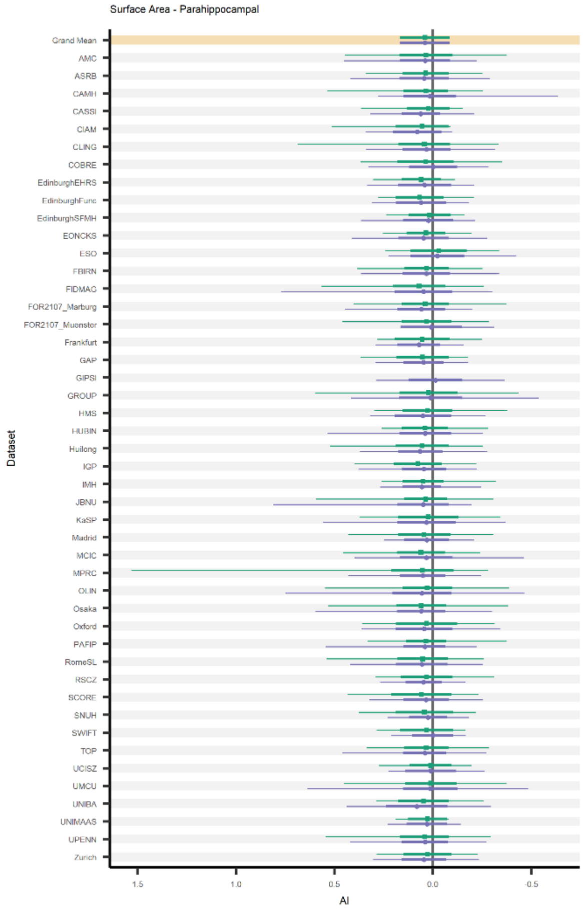
Surface Area - Parahippocampal
Dataset
Grand Mean
AMC
ASRB
CAMH
CASSI
CIAM
CLING
COBRE
EdinburghEHRS
EdinburghFunc
EdinburghSFMH
EONCKS
ESO
FBIRN
FIDMAG
FOR2107_Marburg
FOR2107_Muenster
Frankfurt
GAP
GIPSI
GROUP
HMS
HUBIN
Huilong
IGP
IMH
JBNU
KaSP
Madrid
MCIC
MPRC
OLIN
Osaka
Oxford
PAFIP
RomeSL
RSCZ
SCORE
SNUH
SWIFT
TOP
UCISZ
UMCU
UNIBA
UNIMAAS
UPENN
Zurich
1.5
1.0
0.5
0.0
-0.5
AI

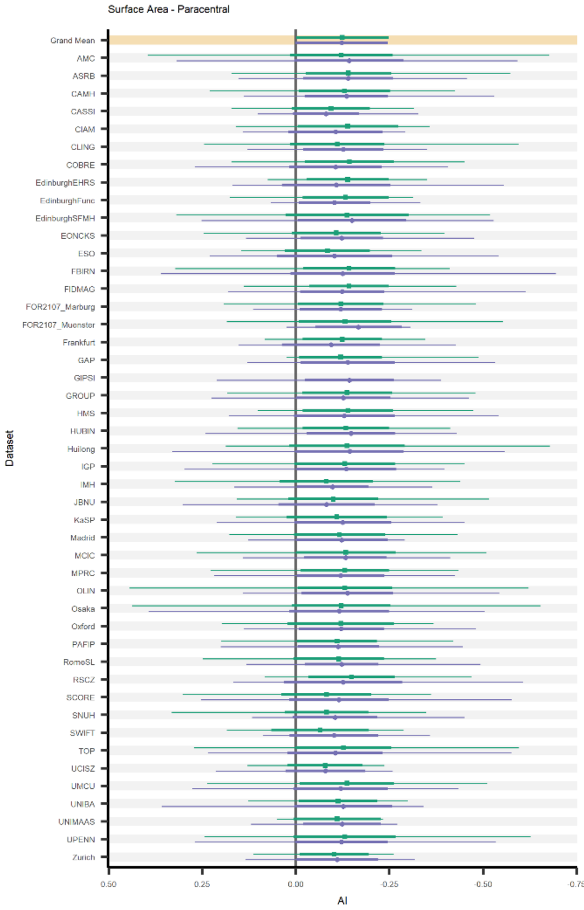
Surface Area - Paracentral
Dataset
Grand Mean
AMC
ASRB
CAMH
CASSI
CIAM
CLING
COBRE
EdinburghEHRS
EdinburghFunc
EdinburghSFMH
EONCKS
ESO
FBIRN
FIDMAG
FOR2107_Marburg
FOR2107_Muenster
Frankfurt
GAP
GIPSI
GROUP
HMS
HUBIN
Huilong
IGP
IMH
JBNU
KaSP
Madrid
MCIC
MPRC
OLIN
Osaka
Oxford
PAFIP
RomeSL
RSCZ
SCORE
SNUH
SWIFT
TOP
UCISZ
UMCU
UNIBA
UNIMAAS
UPENN
Zurich
0.50
0.25
0.00
-0.25
-0.50
-0.75
AI

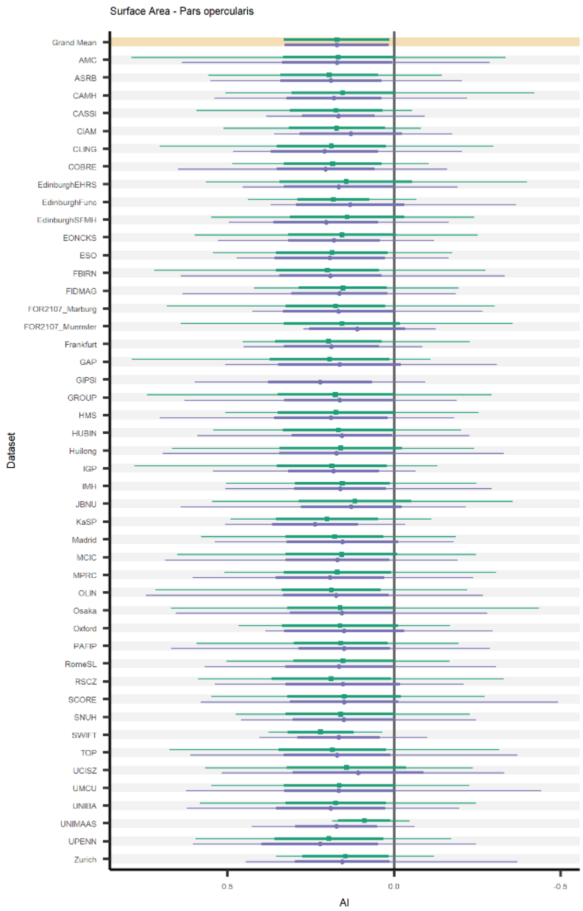
Surface Area - Pars opercularis
Grand Mean
AMC
ASRB
CAMH
CASSI
CIAM
CLING
COBRE
EdinburghEHRS
EdinburghFunc
EdinburghSFMH
EONCKS
ESO
FBIRN
FIDMAG
FOR2107_Marburg
FOR2107_Muenster
Frankfurt
GAP
GIPSI
GROUP
HMS
HUBIN
Huilong
IGP
IMH
JBNU
KaSP
Madrid
MCIC
MPRC
OLIN
Osaka
Oxford
PAFIP
RomeSL
RSCZ
SCORE
SNUH
SWIFT
TOP
UCISZ
UMCU
UNIBA
UNIMAAS
UPENN
Zurich
Dataset
0.5
0.0
-0.5
AI

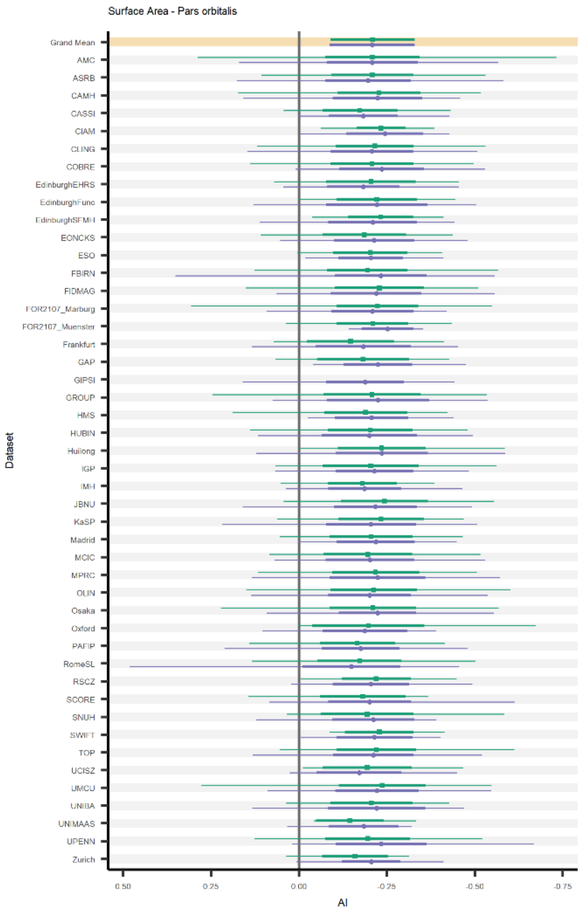
Surface Area - Pars orbitalis
Grand Mean
AMC
ASRB
CAMH
CASSI
CIAM
CLING
COBRE
EdinburghEHRS
EdinburghFunc
EdinburghSFMH
EONCKS
ESO
FBIRN
FIDMAG
FOR2107_Marburg
FOR2107_Muenster
Frankfurt
GAP
GIPSI
GROUP
HMS
HUBIN
Huilong
IGP
IMH
JBNU
KaSP
Madrid
MCIC
MPRC
OLIN
Osaka
Oxford
PAFIP
RomeSL
RSCZ
SCORE
SNUH
SWIFT
TOP
UCISZ
UMCU
UNIBA
UNIMAAS
UPENN
Zurich
Dataset
0.50
0.25
0.00
-0.25
-0.50
-0.75
AI

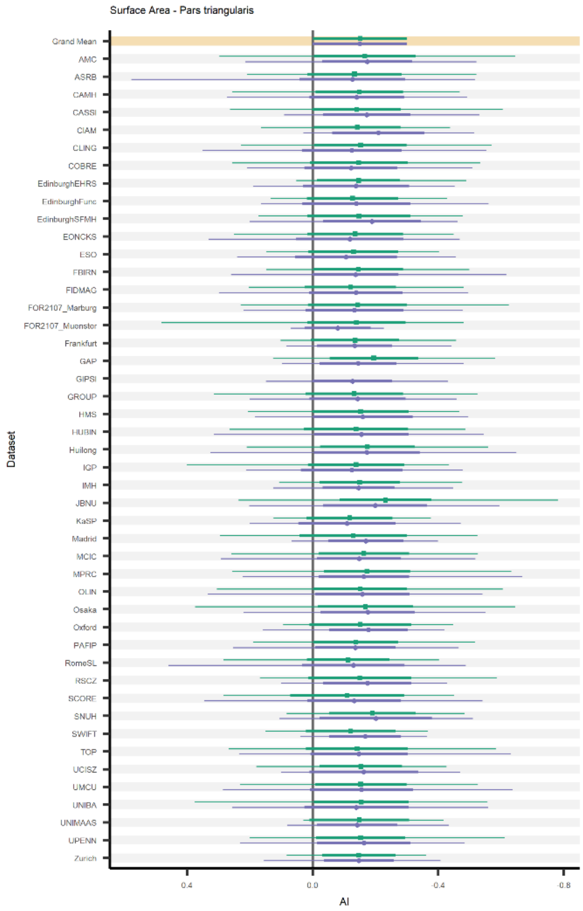
Surface Area - Pars triangularis
Grand Mean
AMC
ASRB
CAMH
CASSI
CIAM
CLING
COBRE
EdinburghEHRS
EdinburghFunc
EdinburghSFMH
EONCKS
ESO
FBIRN
FIDMAG
FOR2107_Marburg
FOR2107_Muenster
Frankfurt
GAP
GIPSI
GROUP
HMS
HUBIN
Huilong
IGP
IMH
JBNU
KaSP
Madrid
MCIC
MPRC
OLIN
Osaka
Oxford
PAFIP
RomeSL
RSCZ
SCORE
SNUH
SWIFT
TOP
UCISZ
UMCU
UNIBA
UNIMAAS
UPENN
Zurich
Dataset
0.4
0.0
-0.4
-0.8
AI

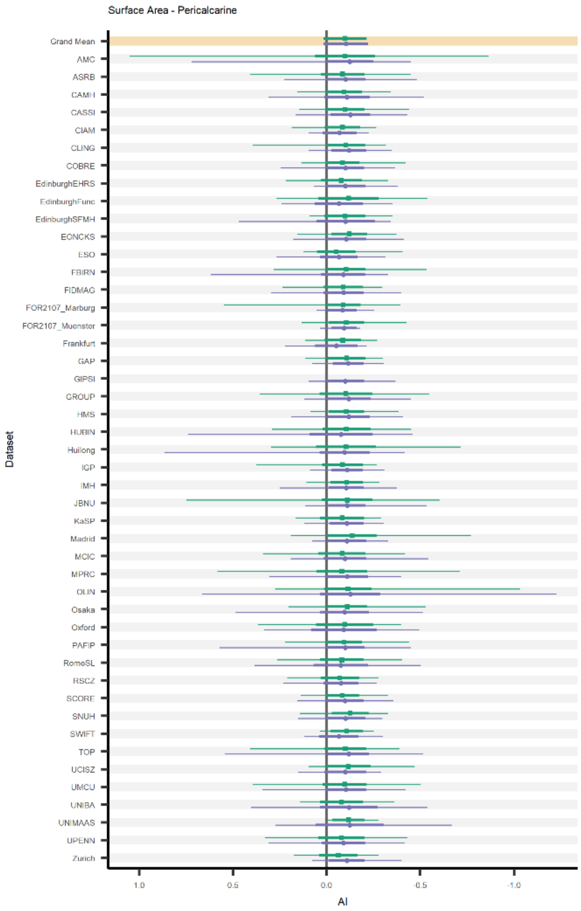
Surface Area - Pericalcarine
Grand Mean
AMC
ASRB
CAMH
CASSI
CIAM
CLING
COBRE
EdinburghEHRS
EdinburghFunc
EdinburghSFMH
EONCKS
ESO
FBIRN
FIDMAG
FOR2107_Marburg
FOR2107_Muenster
Frankfurt
GAP
GIPSI
GROUP
HMS
HUBIN
Huilong
IGP
IMH
JBNU
KaSP
Madrid
MCIC
MPRC
OLIN
Osaka
Oxford
PAFIP
RomeSL
RSCZ
SCORE
SNUH
SWIFT
TOP
UCISZ
UMCU
UNIBA
UNIMAAS
UPENN
Zurich
Dataset
1.0
0.5
0.0
-0.5
-1.0
AI

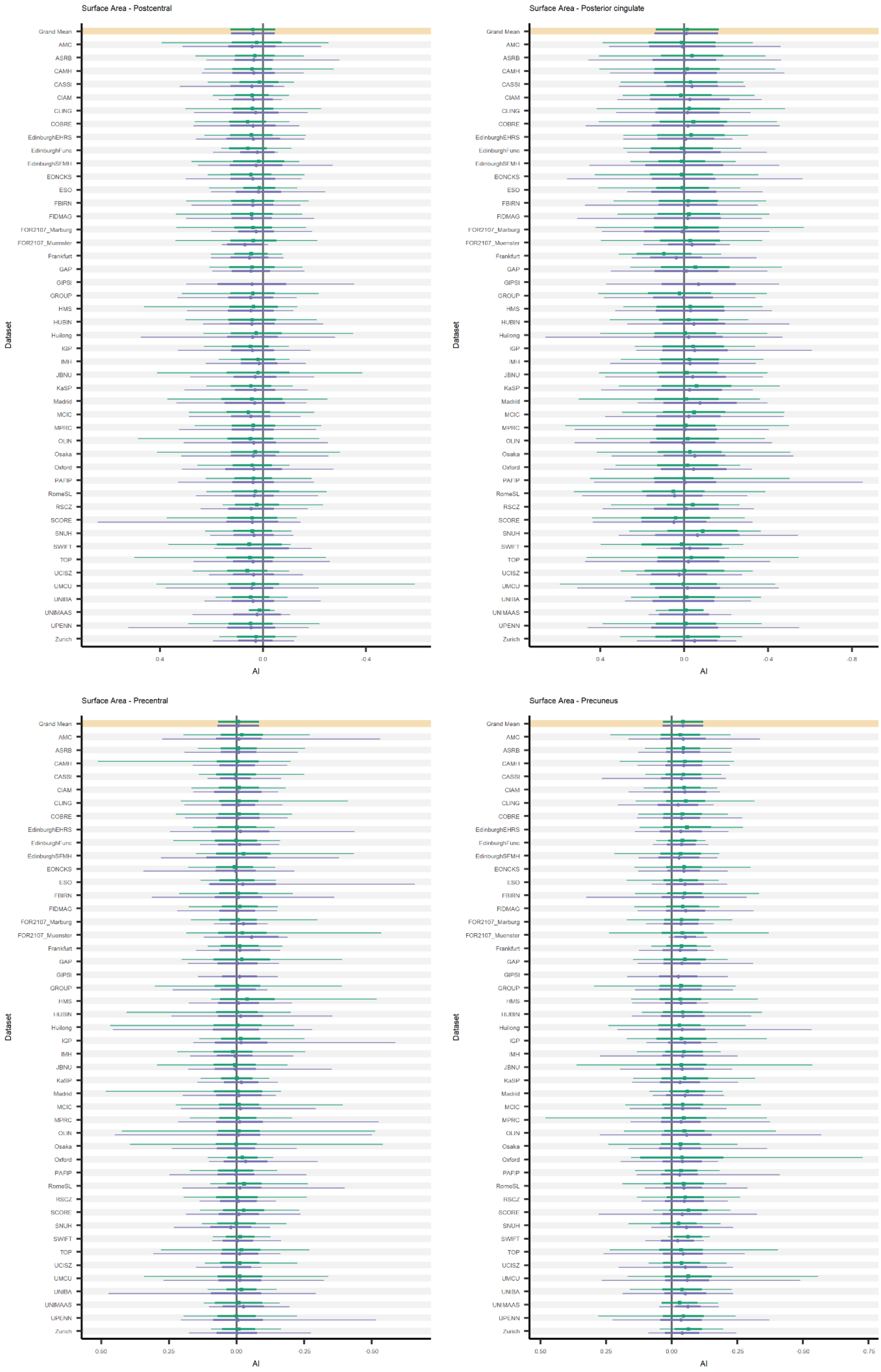

Surface Area - Postcentral
Surface Area - Posterior cingulate
Surface Area - Precentral
Surface Area - Precuneus
Dataset
AI
Grand Mean
AMC
ASRB
CAMH
CASSI
CIAM
CLING
COBRE
EdinburghEHRS
EdinburghFunc
EdinburghSFMH
EONCKS
ESO
FBIRN
FIDMAG
FOR2107_Marburg
FOR2107_Muenster
Frankfurt
GAP
GIPSI
GROUP
HMS
HUBIN
Huilong
IGP
IMH
JBNU
KaSP
Madrid
MCIC
MPRC
OLIN
Osaka
Oxford
PAFIP
RomeSL
RSCZ
SCORE
SNUH
SWIFT
TOP
UCISZ
UMCU
UNIBA
UNIMAAS
UPENN
Zurich

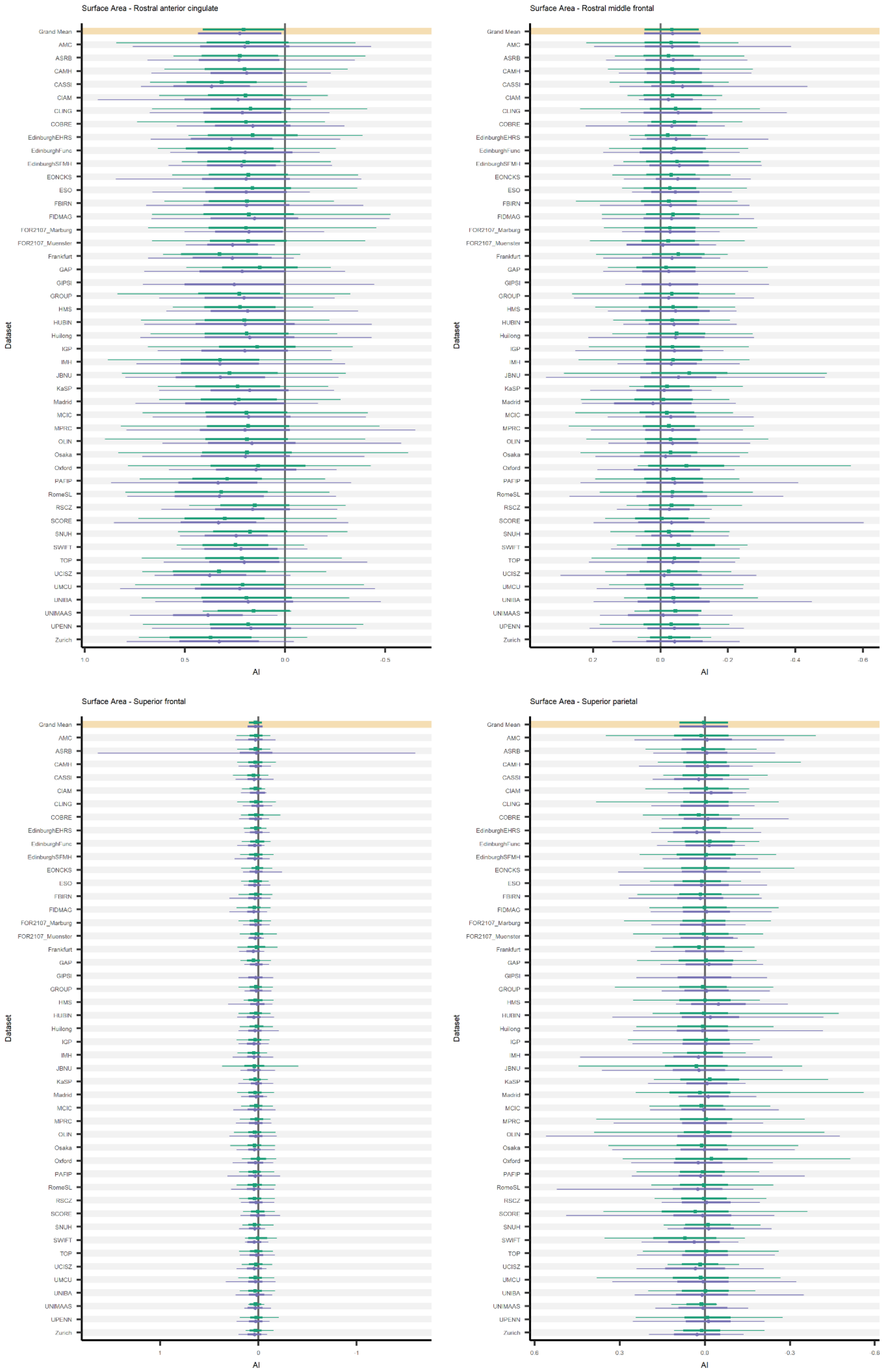
Surface Area - Rostral anterior cingulate
Surface Area - Rostral middle frontal
Surface Area - Superior frontal
Surface Area - Superior parietal
Dataset
AI
Grand Mean
AMC
ASRB
CAMH
CASSI
CIAM
CLING
COBRE
EdinburghEHRS
EdinburghFunc
EdinburghSFMH
EONCKS
ESO
FBIRN
FIDMAG
FOR2107_Marburg
FOR2107_Muenster
Frankfurt
GAP
GIPSI
GROUP
HMS
HUBIN
Huilong
IGP
IMH
JBNU
KaSP
Madrid
MCIC
MPRC
OLIN
Osaka
Oxford
PAFIP
RomeSL
RSCZ
SCORE
SNUH
SWIFT
TOP
UCISZ
UMCU
UNIBA
UNIMAAS
UPENN
Zurich

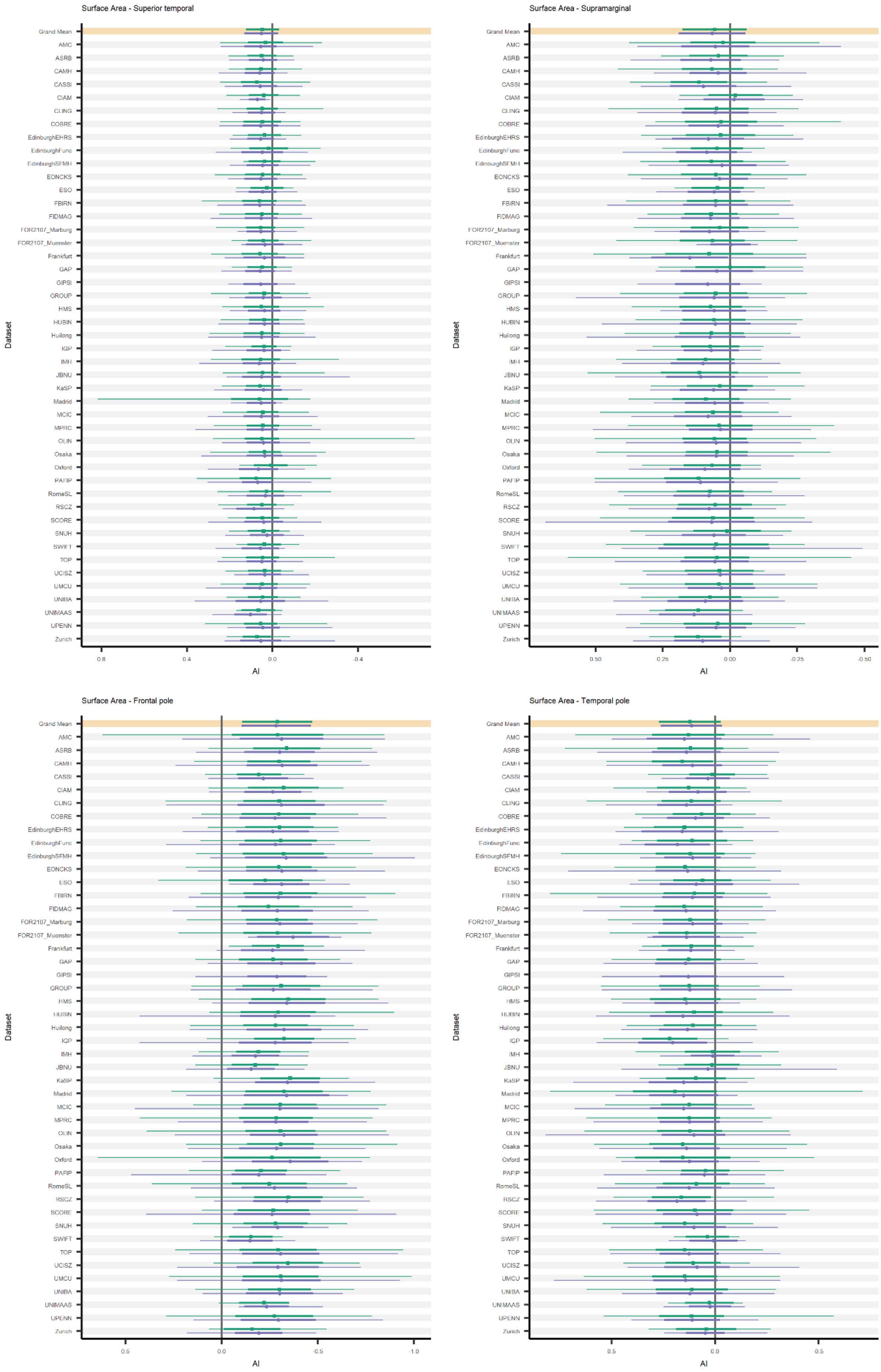
Surface Area - Superior temporal
Surface Area - Supramarginal
Surface Area - Frontal pole
Surface Area - Temporal pole
Dataset
AI
Grand Mean

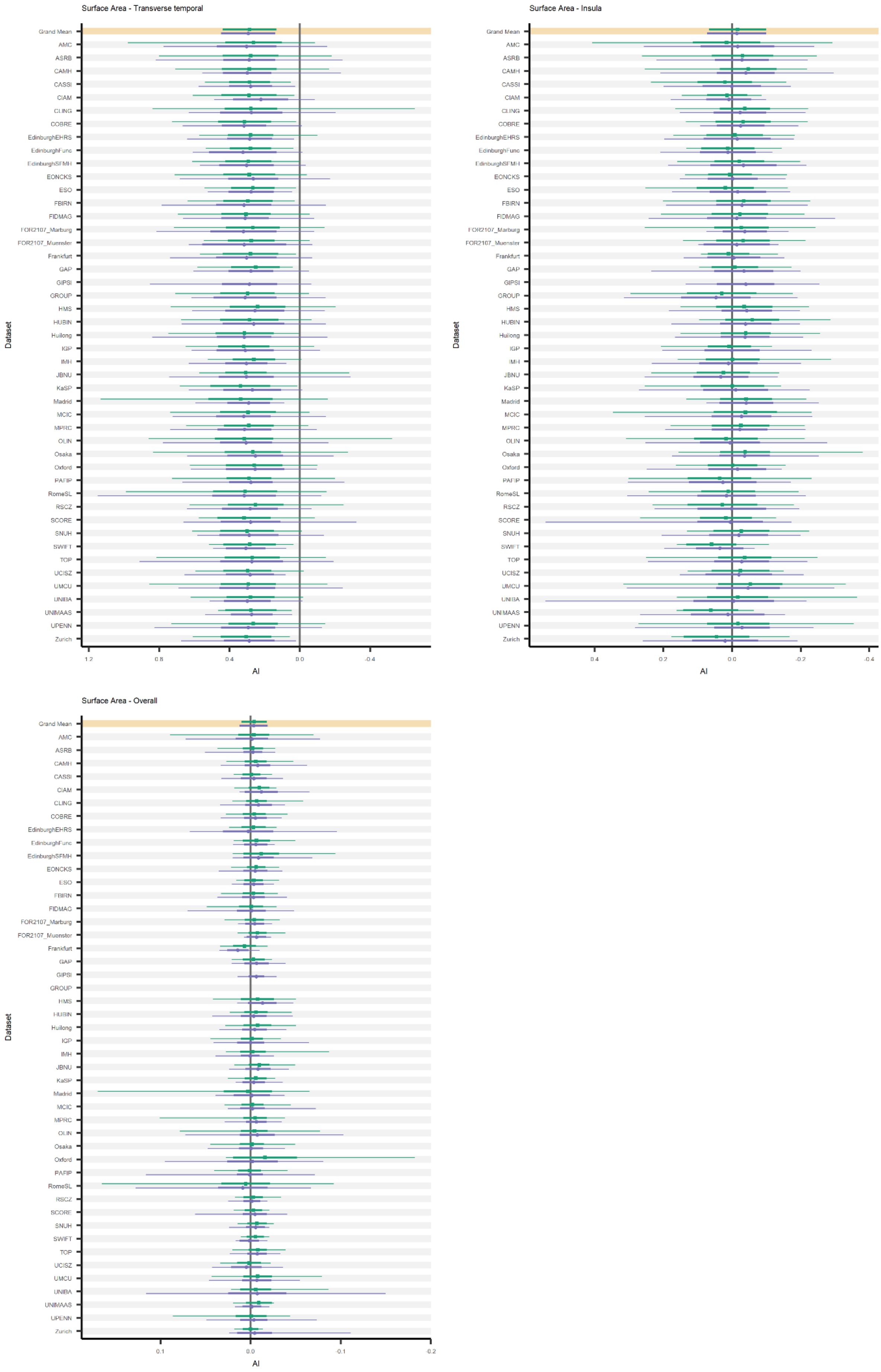
Surface Area - Transverse temporal
Surface Area - Insula
Surface Area - Overall
Dataset
AI
Grand Mean
AMC
ASRB
CAMH
CASSI
CIAM
CLING
COBRE
EdinburghEHRS
EdinburghFunc
EdinburghSFMH
EONCKS
ESO
FBIRN
FIDMAG
FOR2107_Marburg
FOR2107_Muenster
Frankfurt
GAP
GIPSI
GROUP
HMS
HUBIN
Huilong
IGP
IMH
JBNU
KaSP
Madrid
MCIC
MPRC
OLIN
Osaka
Oxford
PAFIP
RomeSL
RSCZ
SCORE
SNUH
SWIFT
TOP
UCISZ
UMCU
UNIBA
UNIMAAS
UPENN
Zurich

**Fig. S3 (page 26-27). Overall and per-dataset average and range for subcortical volume asymmetries.** For each subcortical volume asymmetry measure, the average in controls (green circles) and individuals affected with schizophrenia (purple squares) is shown. The top (highlighted) row contains the grand sample size-weighted mean and standard deviation (thick line segments). The other rows contain per-dataset averages, standard deviations and minimum and maximum values (indicated with thin line segments).

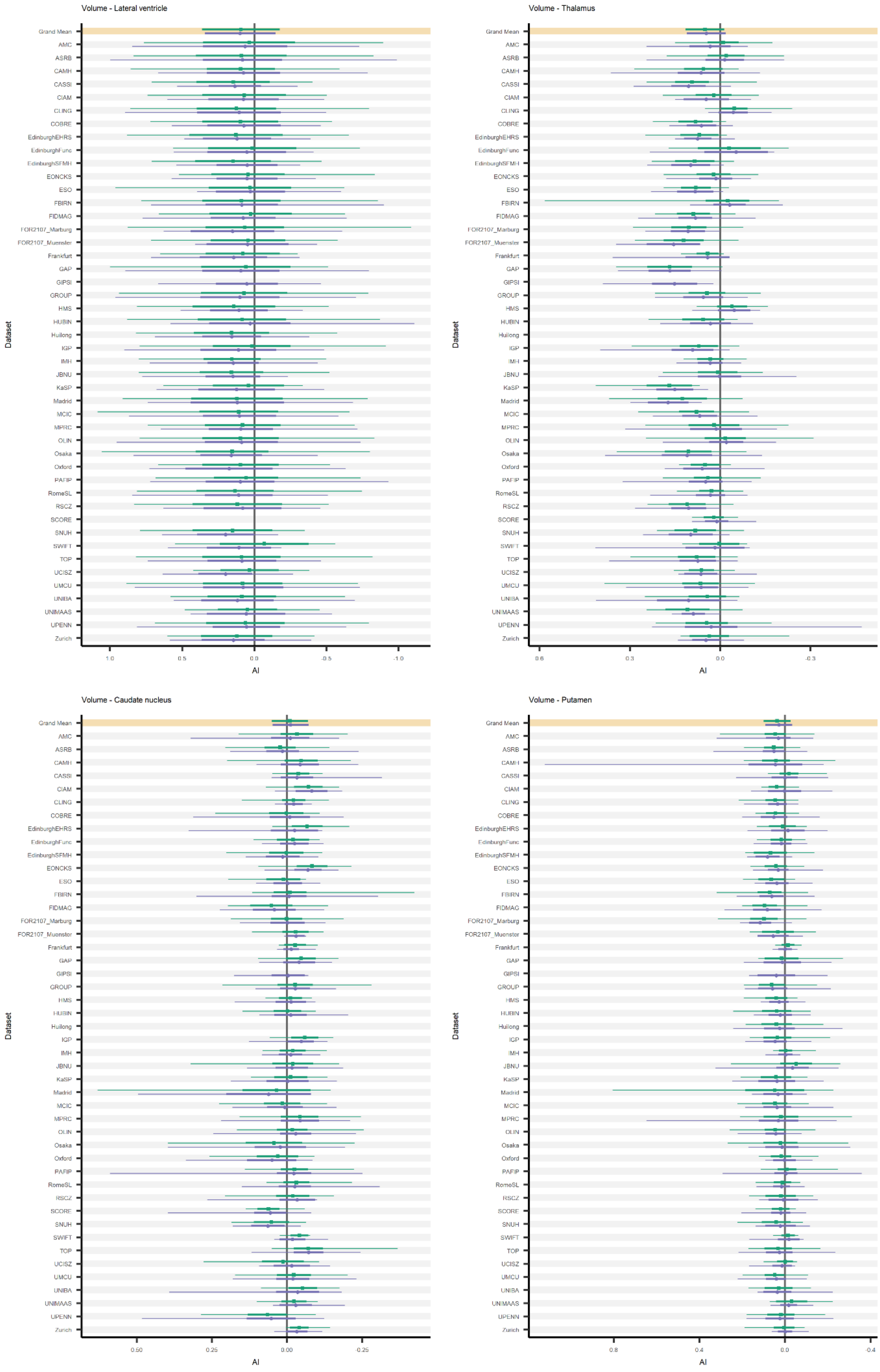
Volume - Lateral ventricle
Volume - Thalamus
Volume - Caudate nucleus
Volume - Putamen
Dataset
AI
Grand Mean
AMC
ASRB
CAMH
CASSI
CIAM
CLING
COBRE
EdinburghEHRS
EdinburghFunc
EdinburghSFMH
EONCKS
ESO
FBIRN
FIDMAG
FOR2107_Marburg
FOR2107_Muenster
Frankfurt
GAP
GIPSI
GROUP
HMS
HUBIN
Huilong
IGP
IMH
JBNU
KaSP
Madrid
MCIC
MPRC
OLIN
Osaka
Oxford
PAFIP
RomeSL
RSCZ
SCORE
SNUH
SWIFT
TOP
UCISZ
UMCU
UNIBA
UNIMAAS
UPENN
Zurich

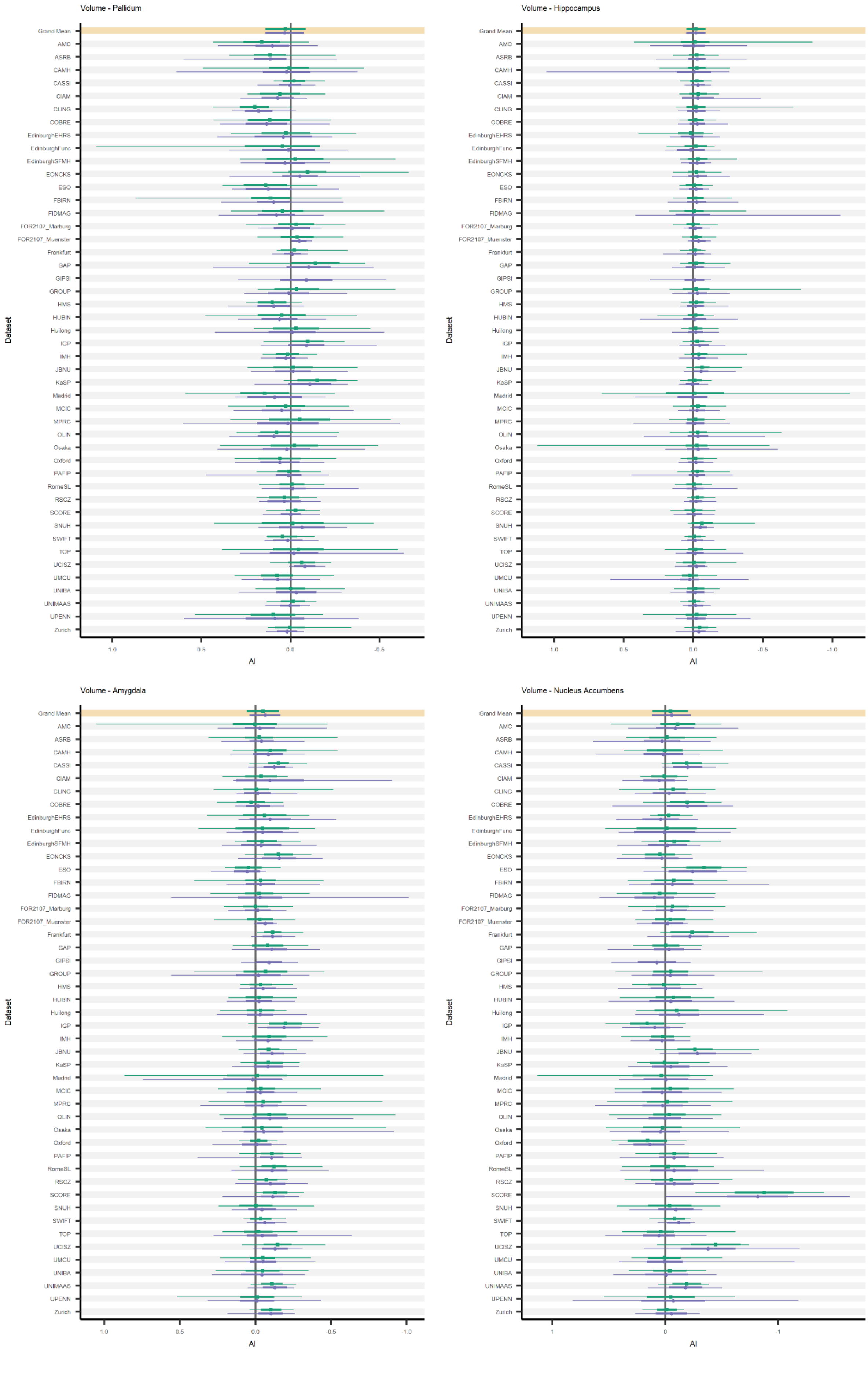
Volume - Pallidum
Dataset
Grand Mean
AMC
ASRB
CAMH
CASSI
CIAM
CLING
COBRE
EdinburghEHRS
EdinburghFunc
EdinburghSFMH
EONCKS
ESO
FBIRN
FIDMAG
FOR2107_Marburg
FOR2107_Muenster
Frankfurt
GAP
GIPSI
GROUP
HMS
HUBIN
Huilong
IGP
IMH
JBNU
KaSP
Madrid
MCIC
MPRC
OLIN
Osaka
Oxford
PAFIP
RomeSL
RSCZ
SCORE
SNUH
SWIFT
TOP
UCISZ
UMCU
UNIBA
UNIMAAS
UPENN
Zurich
1.0
0.5
0.0
-0.5
AI
Volume - Hippocampus
Dataset
Grand Mean
AMC
ASRB
CAMH
CASSI
CIAM
CLING
COBRE
EdinburghEHRS
EdinburghFunc
EdinburghSFMH
EONCKS
ESO
FBIRN
FIDMAG
FOR2107_Marburg
FOR2107_Muenster
Frankfurt
GAP
GIPSI
GROUP
HMS
HUBIN
Huilong
IGP
IMH
JBNU
KaSP
Madrid
MCIC
MPRC
OLIN
Osaka
Oxford
PAFIP
RomeSL
RSCZ
SCORE
SNUH
SWIFT
TOP
UCISZ
UMCU
UNIBA
UNIMAAS
UPENN
Zurich
1.0
0.5
0.0
-0.5
-1.0
AI
Volume - Amygdala
Dataset
Grand Mean
AMC
ASRB
CAMH
CASSI
CIAM
CLING
COBRE
EdinburghEHRS
EdinburghFunc
EdinburghSFMH
EONCKS
ESO
FBIRN
FIDMAG
FOR2107_Marburg
FOR2107_Muenster
Frankfurt
GAP
GIPSI
GROUP
HMS
HUBIN
Huilong
IGP
IMH
JBNU
KaSP
Madrid
MCIC
MPRC
OLIN
Osaka
Oxford
PAFIP
RomeSL
RSCZ
SCORE
SNUH
SWIFT
TOP
UCISZ
UMCU
UNIBA
UNIMAAS
UPENN
Zurich
1.0
0.5
0.0
-0.5
-1.0
AI
Volume - Nucleus Accumbens
Dataset
Grand Mean
AMC
ASRB
CAMH
CASSI
CIAM
CLING
COBRE
EdinburghEHRS
EdinburghFunc
EdinburghSFMH
EONCKS
ESO
FBIRN
FIDMAG
FOR2107_Marburg
FOR2107_Muenster
Frankfurt
GAP
GIPSI
GROUP
HMS
HUBIN
Huilong
IGP
IMH
JBNU
KaSP
Madrid
MCIC
MPRC
OLIN
Osaka
Oxford
PAFIP
RomeSL
RSCZ
SCORE
SNUH
SWIFT
TOP
UCISZ
UMCU
UNIBA
UNIMAAS
UPENN
Zurich
1
0
-1
AI

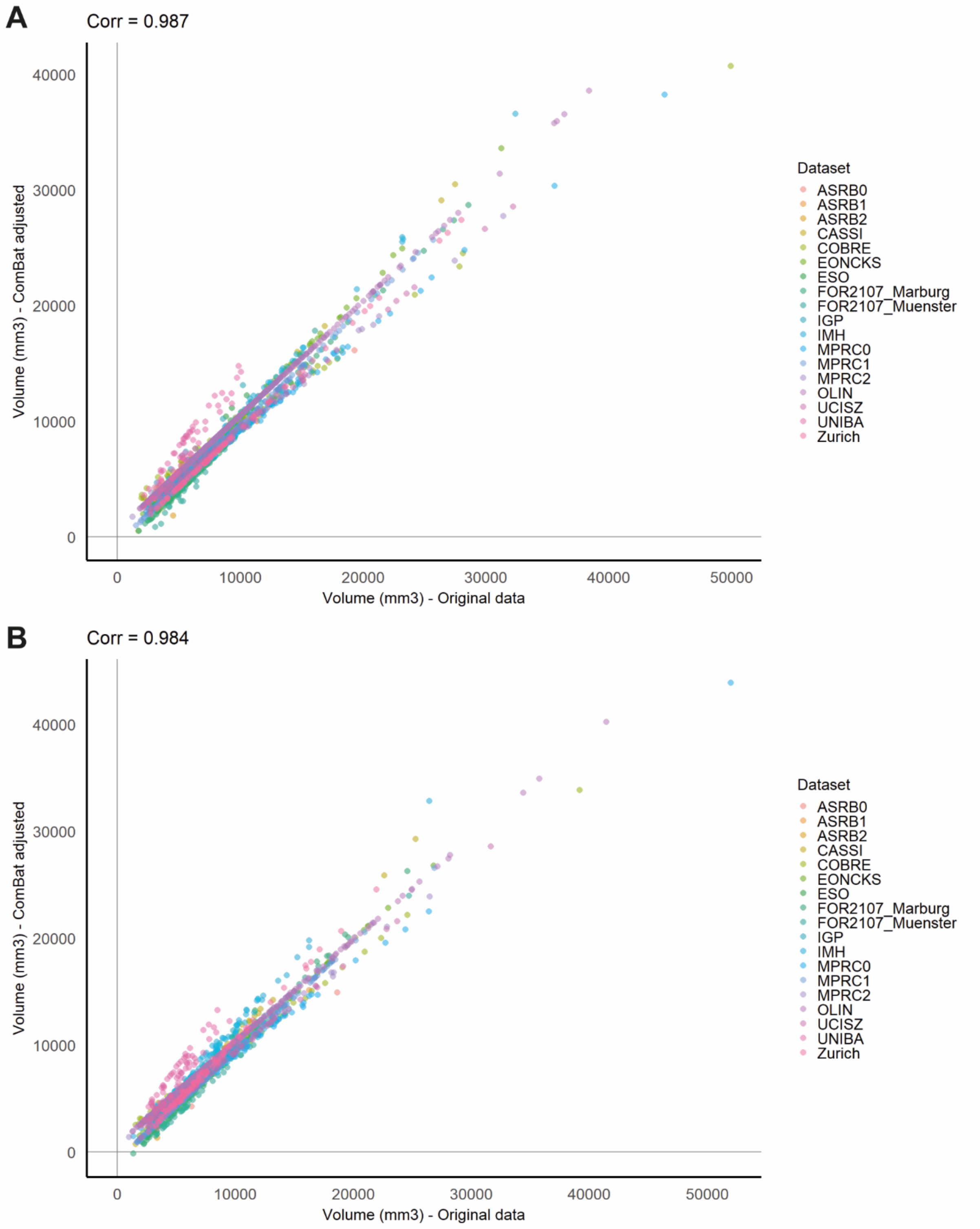

**Fig. S4. Original and ComBat adjusted lateral ventricle volumes.** Original lateral ventricle volumes (x-axis) versus ComBat (1) adjusted measurements (y-axis) for the left **(A)** and right **(B)** hemispheres are shown for individuals across 14 datasets (color-coded, two datasets – ASRB and MPRC – are split because of multiple scanners). Correlations (Corr) between original and ComBat adjusted measurements are shown above each figure. One subject from the FOR2107 Marburg dataset had a slightly negative adjusted right lateral ventricle volume and was therefore excluded.

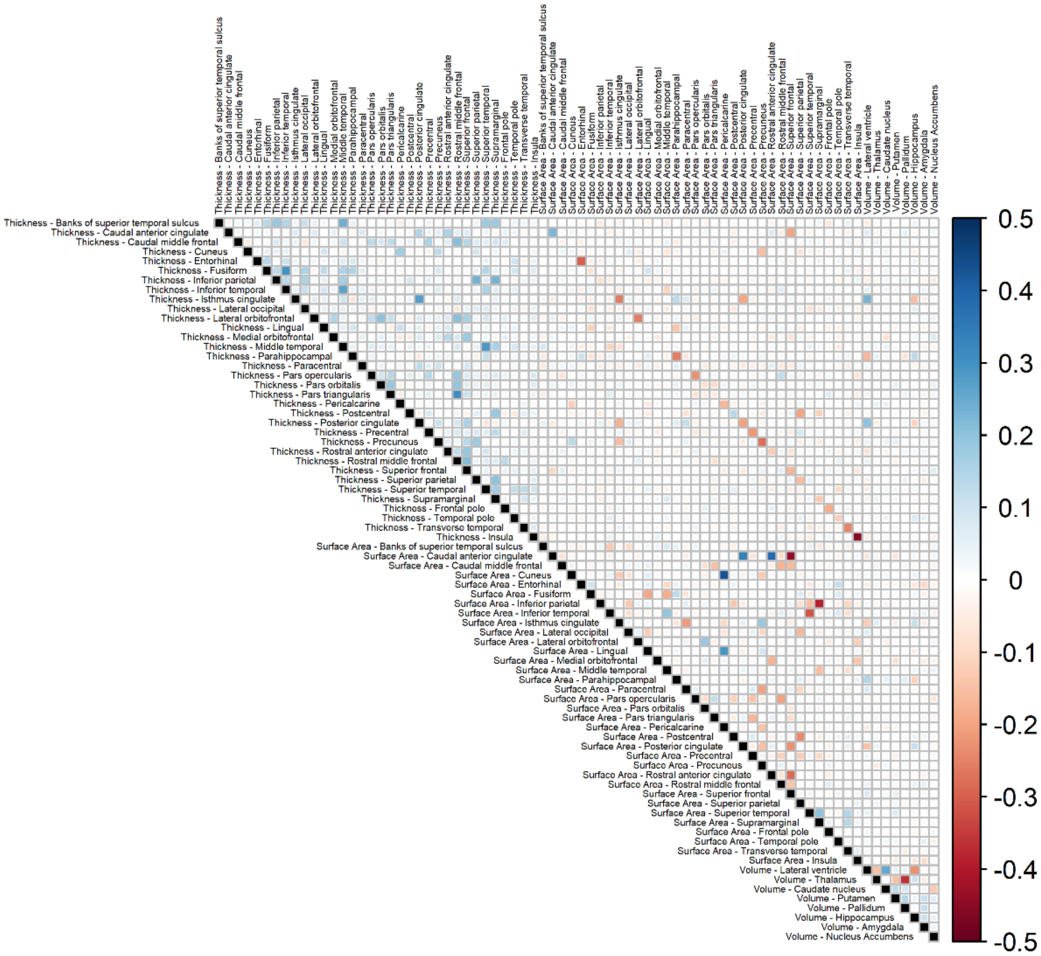

**Fig. S5. Correlations between structural asymmetries in the 14 datasets available for multivariate analysis (i.e. where individual-level data were available to the central analysis team).** The correlations between AIs are shown at the intersections of rows and columns. Positive correlations are shown in blue shades, negative correlations are shown in red shades. Figure generated using the *corrplot* package in R (2).

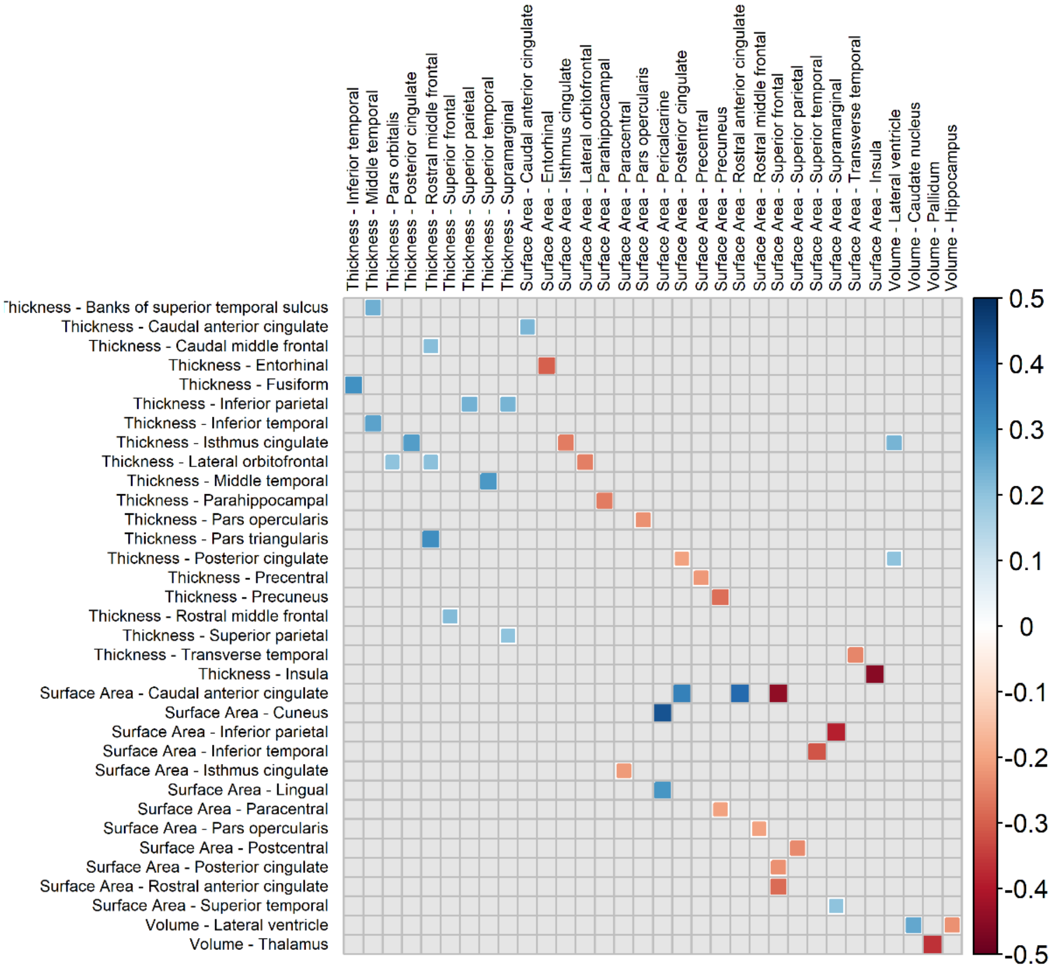

**Fig. S6. Correlations > 0.2 between structural asymmetries in the 14 datasets available for multivariate analysis (i.e. where individual-level data were available to the central analysis team).** The correlations between AIs are shown at the intersections of rows and columns. Only correlations > 0.2 are shown and structural asymmetries not having any such large correlations are excluded from the matrix (i.e. this figure shows a subset of the same correlation matrix as in Fig. S5, to aid in visualization of the larger correlations only). Positive correlations are shown in blue shades, negative correlations are shown in red shades. Figure generated using the *corrplot* package in R (2).

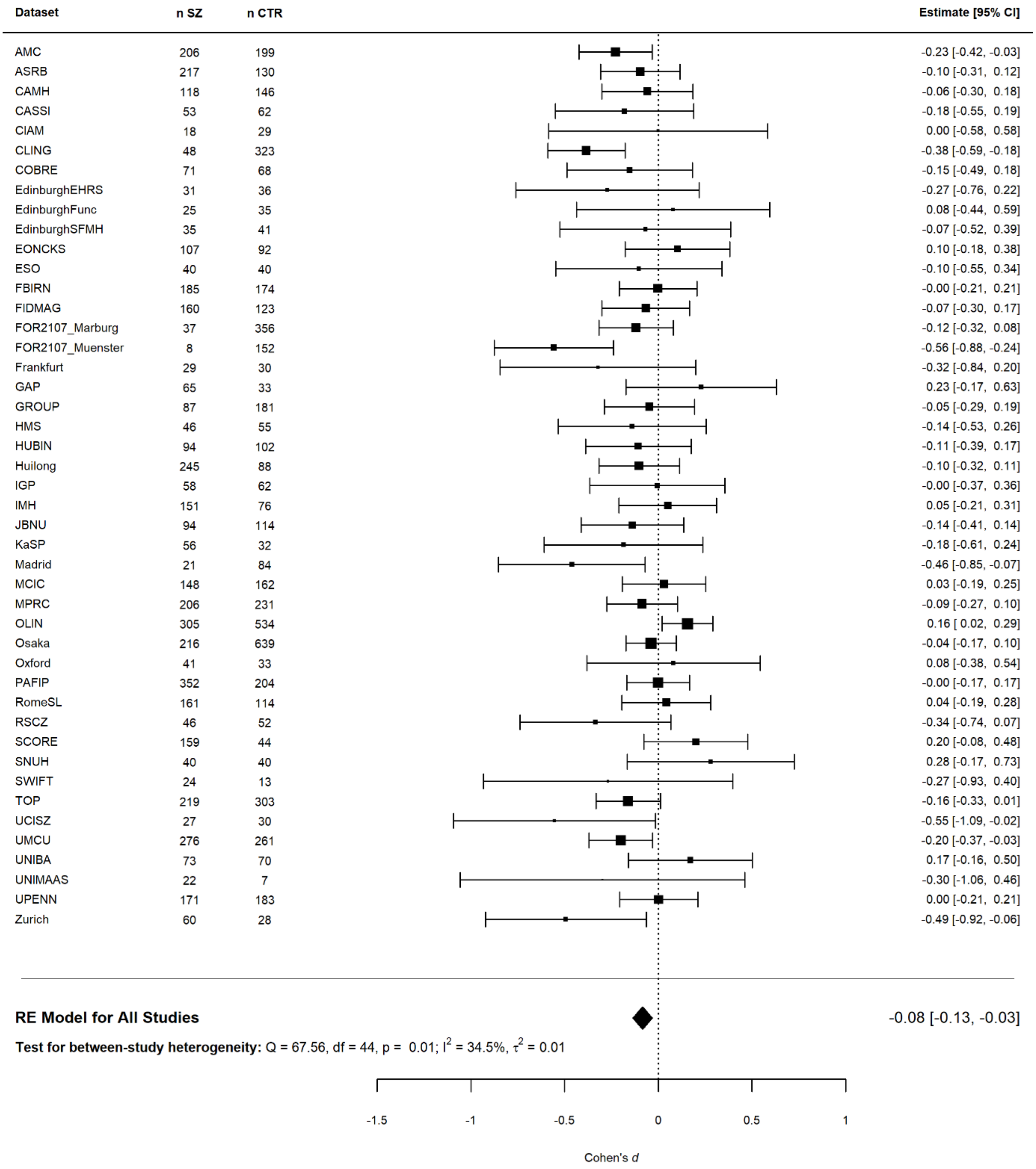

**Fig. S7. Forest plot for random effects meta-analysis of rostral anterior cingulate thickness asymmetry differences between schizophrenia individuals and unaffected controls.** Shown are the per-dataset effect sizes, including confidence intervals, of affected individuals (n SZ) and unaffected controls (n CTR). Cohen's *d* dot sizes represent relative dataset sizes. The meta-analyzed effect sizes across all studies is shown (black diamond), as well as between-dataset heterogeneity statistics (Cochran's Q test statistics).

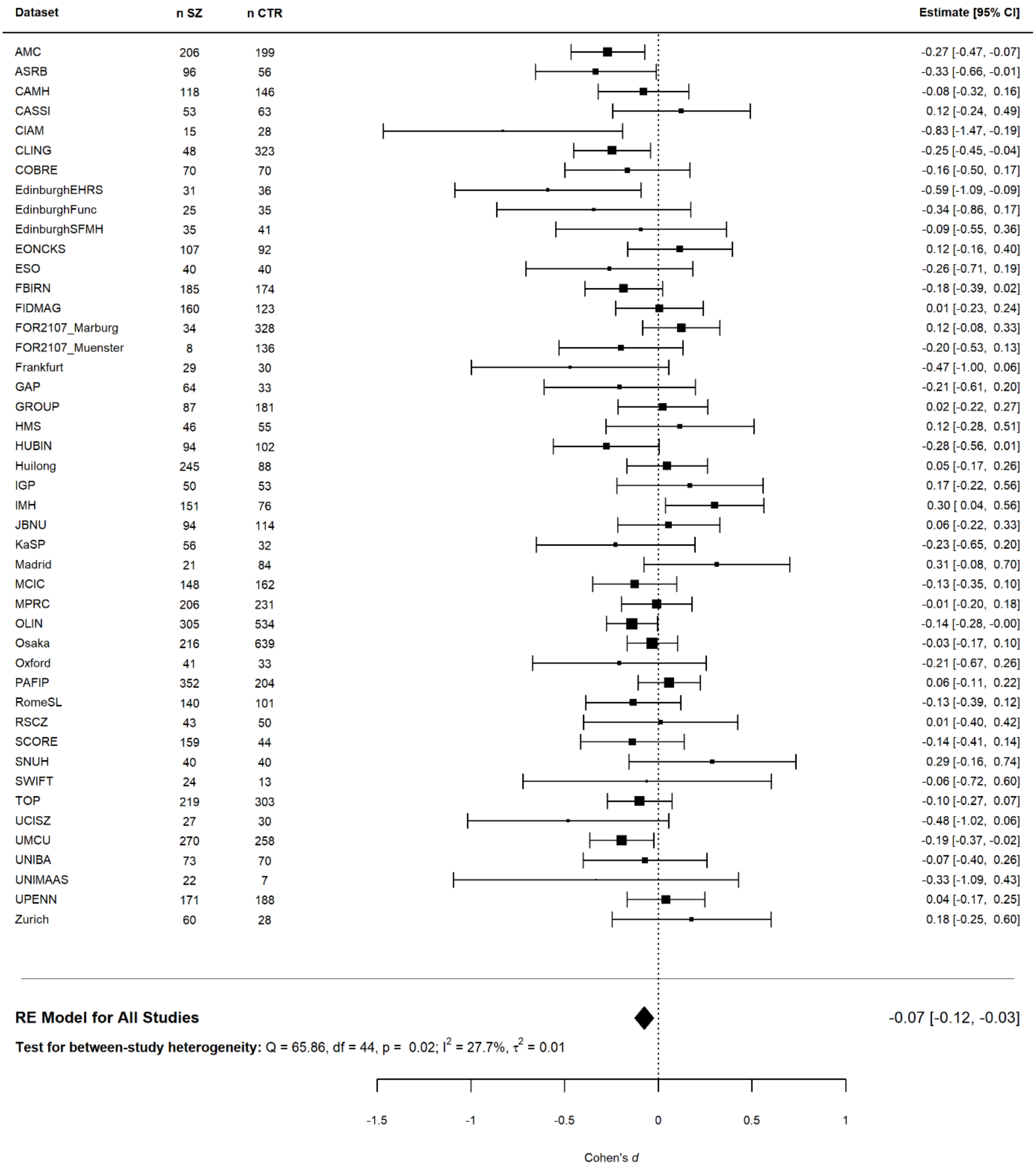

**Fig. S8. Forest plot for random effects meta-analysis of middle temporal gyrus thickness asymmetry differences between schizophrenia individuals and unaffected controls.** Shown are the per-dataset effect sizes, including confidence intervals, of affected individuals (n SZ) and unaffected controls (n CTR). Cohen's *d* dot sizes represent relative dataset sizes. The meta-analyzed effect sizes across all studies is shown (black diamond), as well as between-dataset heterogeneity statistics (Cochran's Q test statistics).

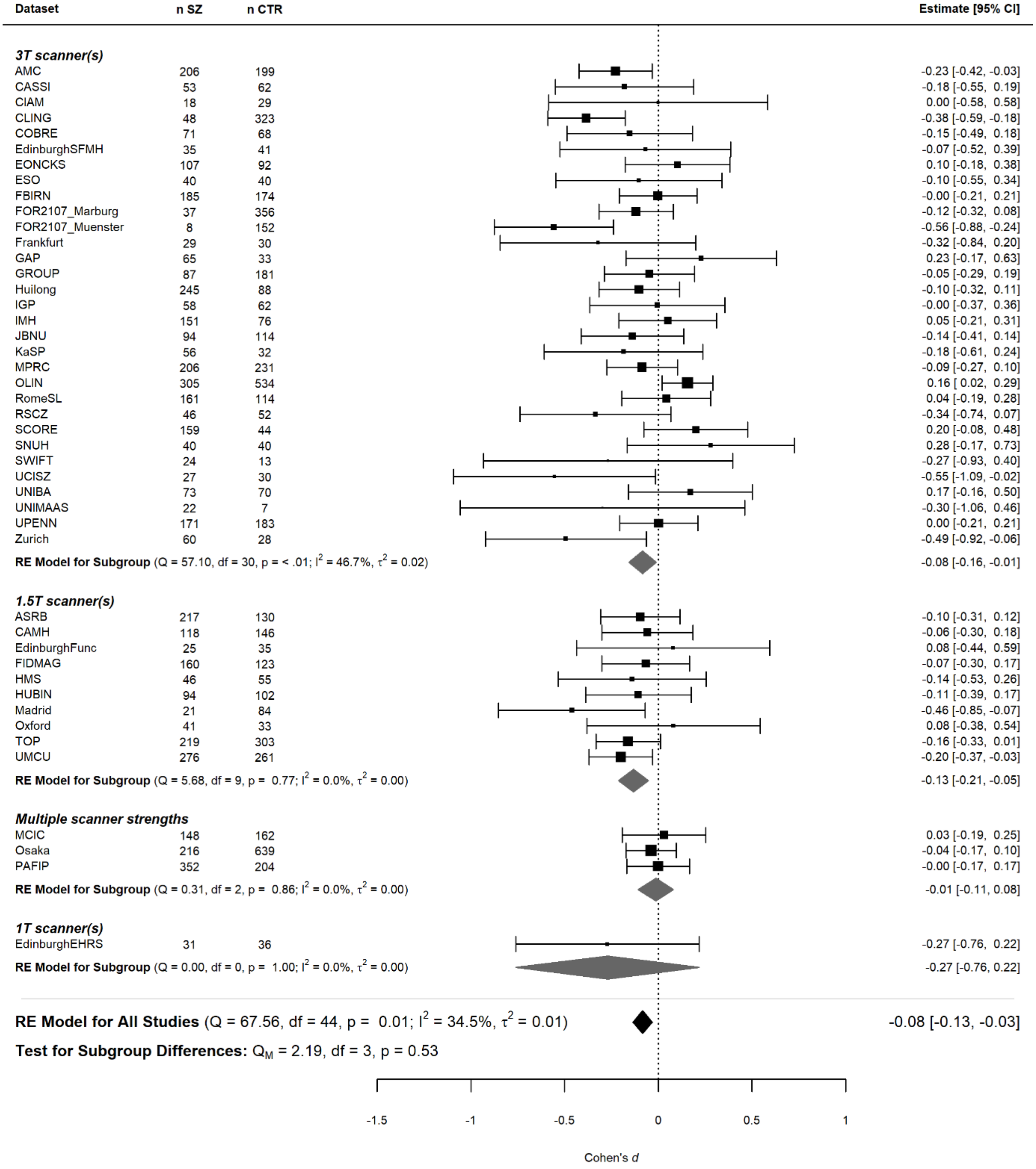

**Fig. S9. Forest plot grouped by scanner field strength, for random effects meta-analysis of rostral anterior cingulate thickness asymmetry differences between individuals with schizophrenia and unaffected controls.** Shown are the per-dataset effect sizes, including confidence intervals, and the numbers of affected individuals (n SZ) and unaffected controls (n CTR). Cohen's *d* dot sizes represent relative dataset sizes. Meta-analyzed effect sizes are shown at a group level (gray diamonds) and across all studies (black diamonds), as well as between-dataset heterogeneity statistics (Cochran's Q test statistics). Scanner field strength group differences were tested using an omnibus test for heterogeneity. Groups are ordered from largest to smallest.

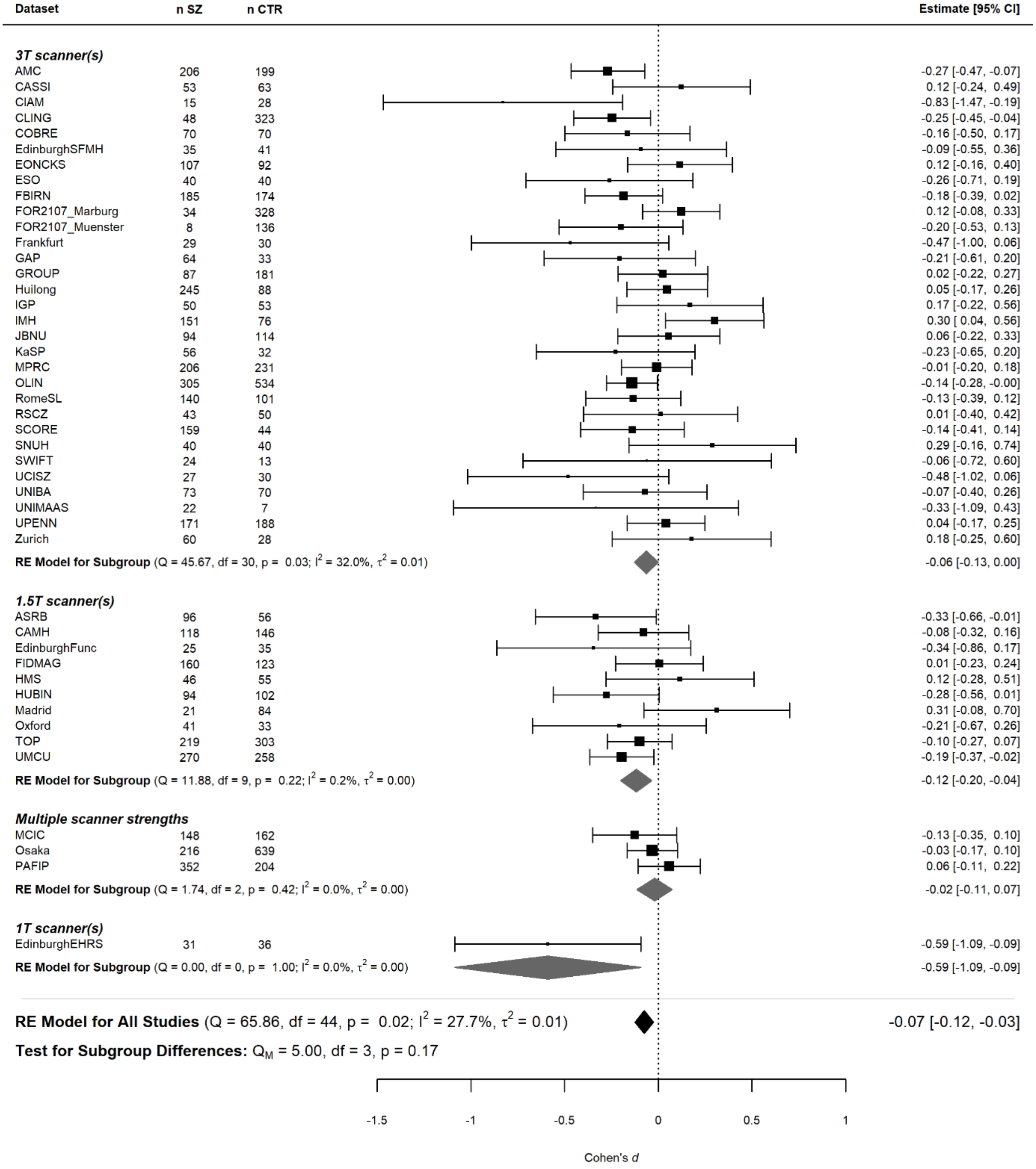

**Fig. S10. Forest plot grouped by scanner field strength, for random effects meta-analysis of middle temporal gyrus thickness asymmetry differences between schizophrenia individuals and unaffected controls.** Shown are the per-dataset effect sizes, including confidence intervals, and the numbers of affected individuals (n SZ) and unaffected controls (n CTR). Cohen's *d* dot sizes represent relative dataset sizes. Meta-analyzed effect sizes are shown at a group level (gray diamonds) and across all studies (black diamonds), as well as between-dataset heterogeneity statistics (Cochran's Q test statistics). Scanner field strength group differences were tested using an omnibus test for heterogeneity. Groups are ordered from largest to smallest.

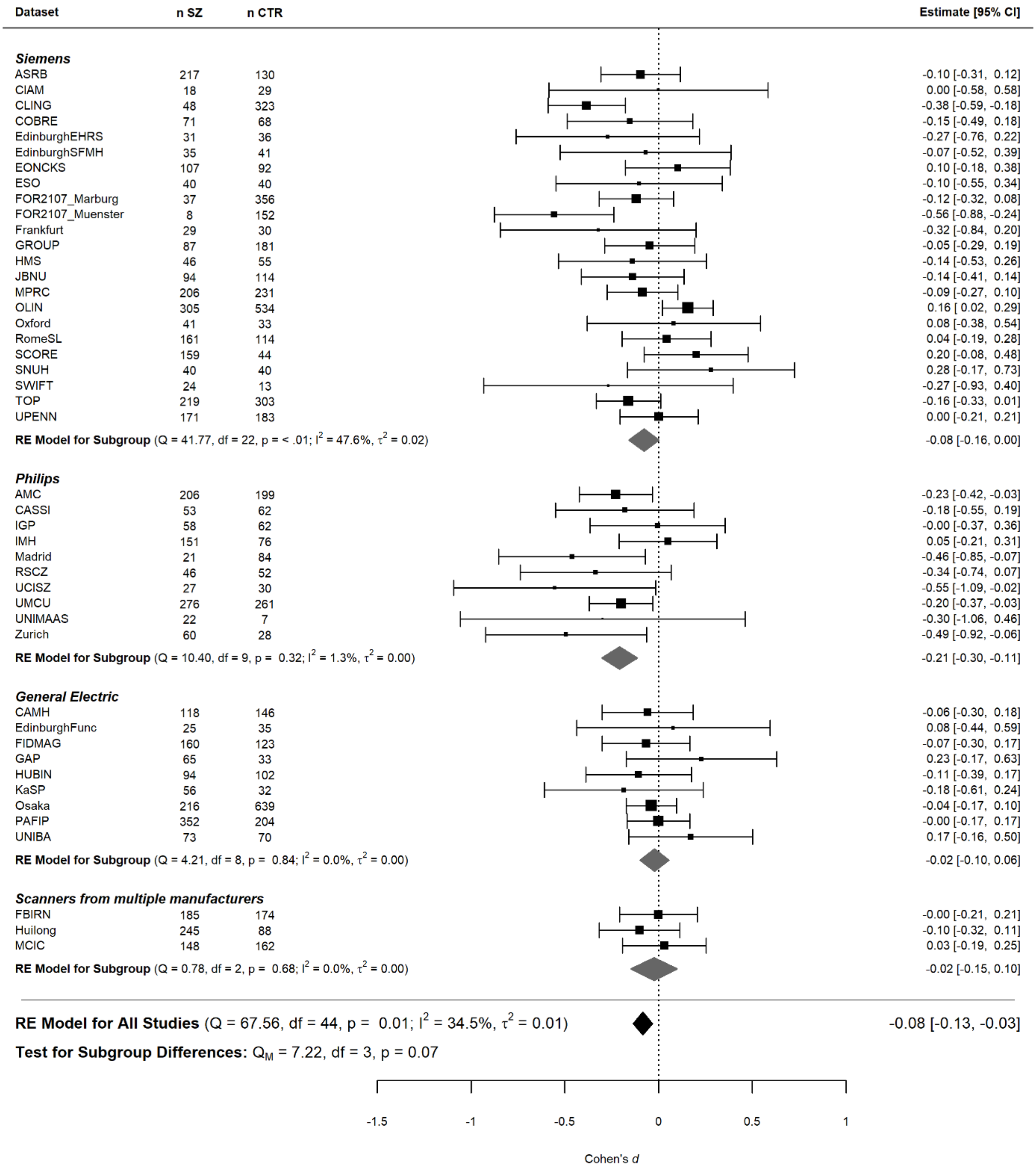

**Fig. S11. Forest plot grouped by scanner manufacturer, for random effects meta-analysis of rostral anterior cingulate thickness asymmetry differences between schizophrenia individuals and unaffected controls.** Shown are the per-dataset effect sizes, including confidence intervals, and the numbers of affected individuals (n SZ) and unaffected controls (n CTR). Cohen's *d* dot sizes represent relative dataset sizes. Meta-analyzed effect sizes are shown at a group level (gray diamonds) and across all studies (black diamonds), as well as between-dataset heterogeneity statistics (Cochran's Q test statistics). Scanner manufacturer group differences were tested using an omnibus test for heterogeneity. Groups are ordered from largest to smallest.

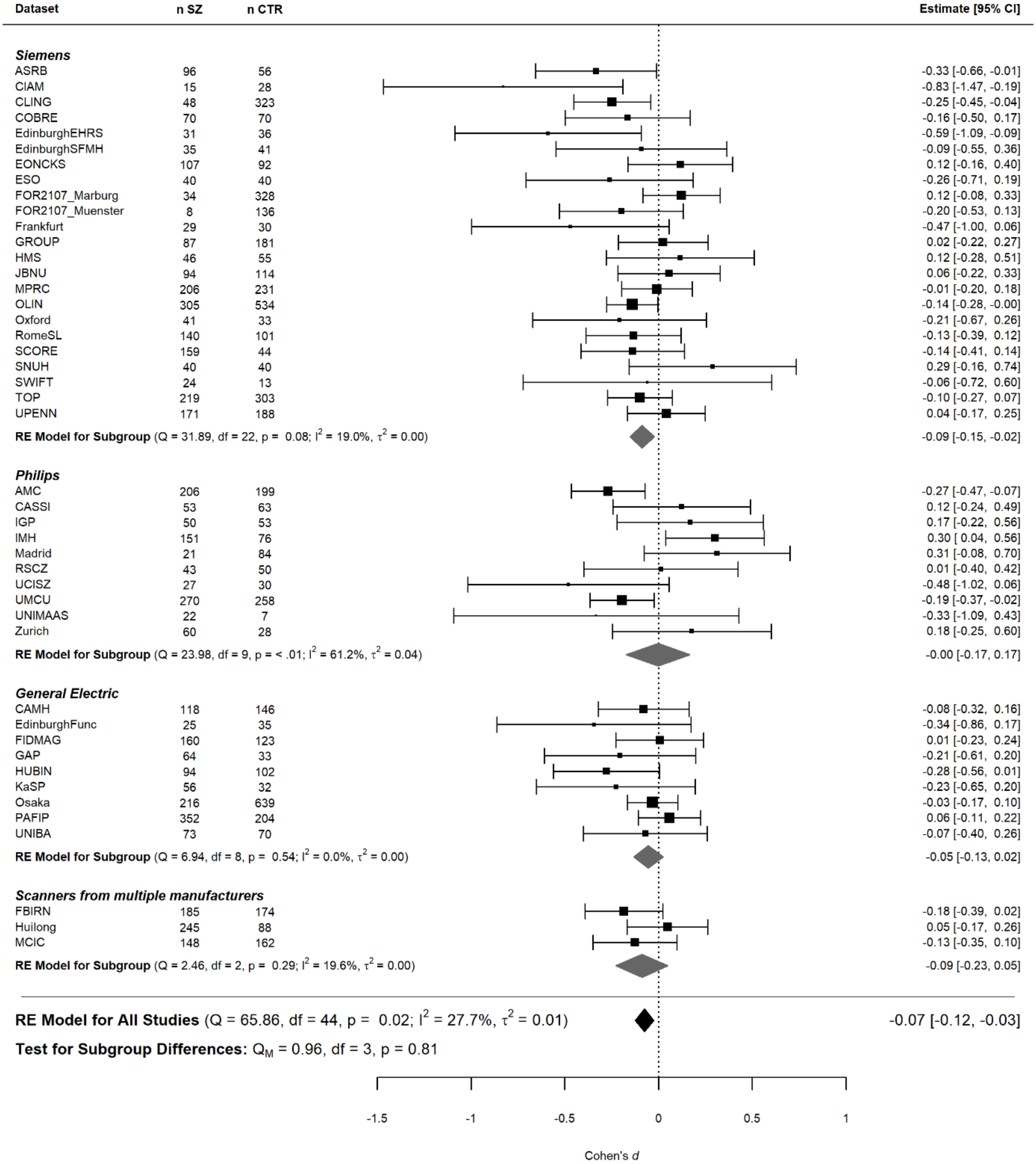

**Fig. S12. Forest plot grouped by scanner manufacturer, for random effects meta-analysis of middle temporal gyrus asymmetry differences between schizophrenia individuals and unaffected controls.** Shown are the per-dataset effect sizes, including confidence intervals, and the numbers of affected individuals (n SZ) and unaffected controls (n CTR). Cohen's *d* dot sizes represent relative dataset sizes. Meta-analyzed effect sizes are shown at a group level (gray diamonds) and across all studies (black diamonds), as well as between-dataset heterogeneity statistics (Cochran's Q test statistics). Scanner manufacturer group differences were tested using an omnibus test for heterogeneity. Groups are ordered from largest to smallest.

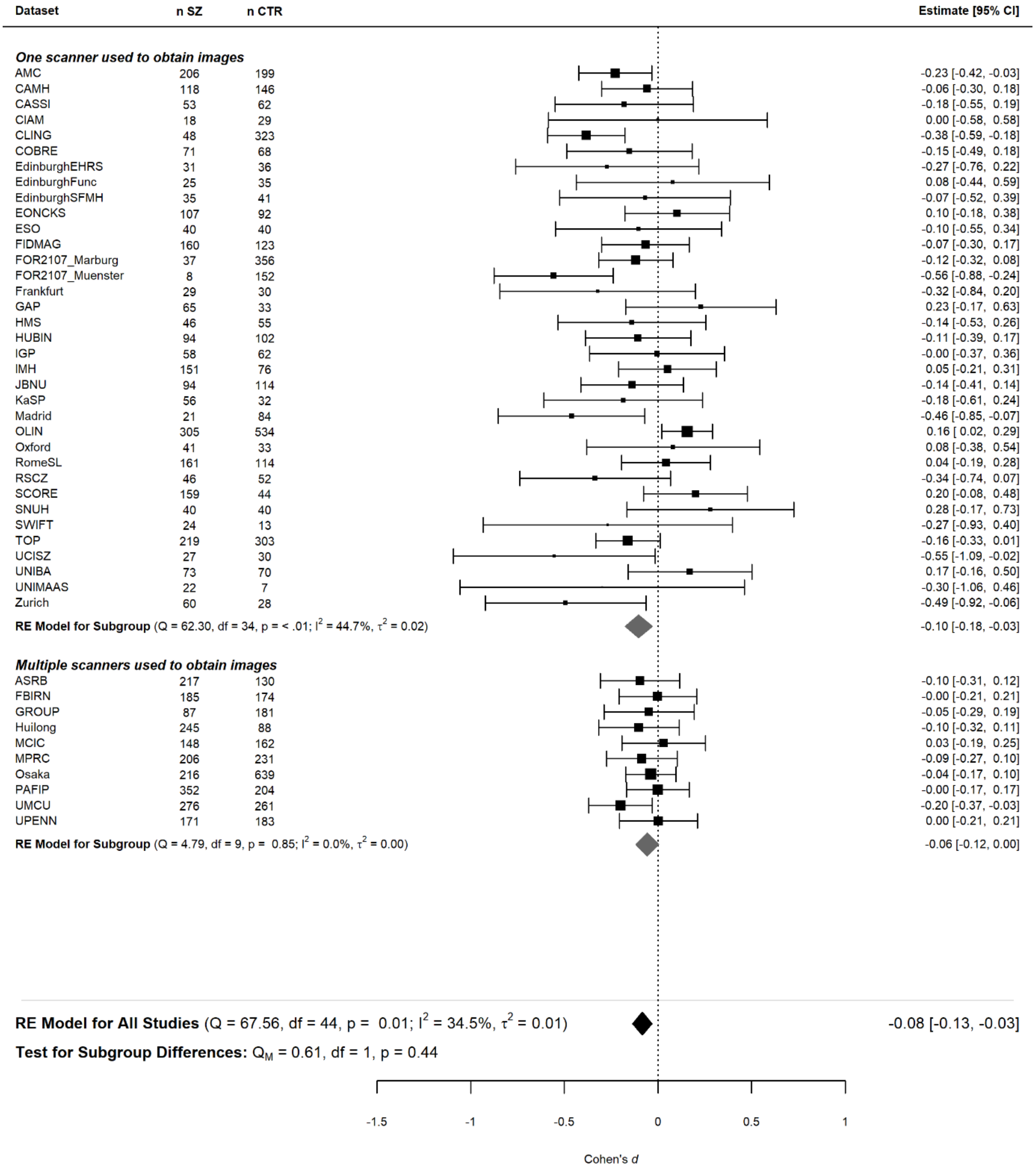

**Fig. S13. Forest plot where datasets are grouped by use of a single scanner versus multiple scanners, for random effects meta-analysis of rostral anterior cingulate thickness asymmetry differences between schizophrenia individuals and unaffected controls.** Shown are the per-dataset effect sizes, including confidence intervals, and the numbers of affected individuals (n SZ) and unaffected controls (n CTR). Cohen's *d* dot sizes represent relative dataset sizes. Meta-analyzed effect sizes are shown at a group level (gray diamonds) and across all studies (black diamonds), as well as between-dataset heterogeneity statistics (Cochran's Q test statistics). Single scanner versus multiple scanner group differences were tested using an omnibus test for heterogeneity. Groups are ordered from largest to smallest.

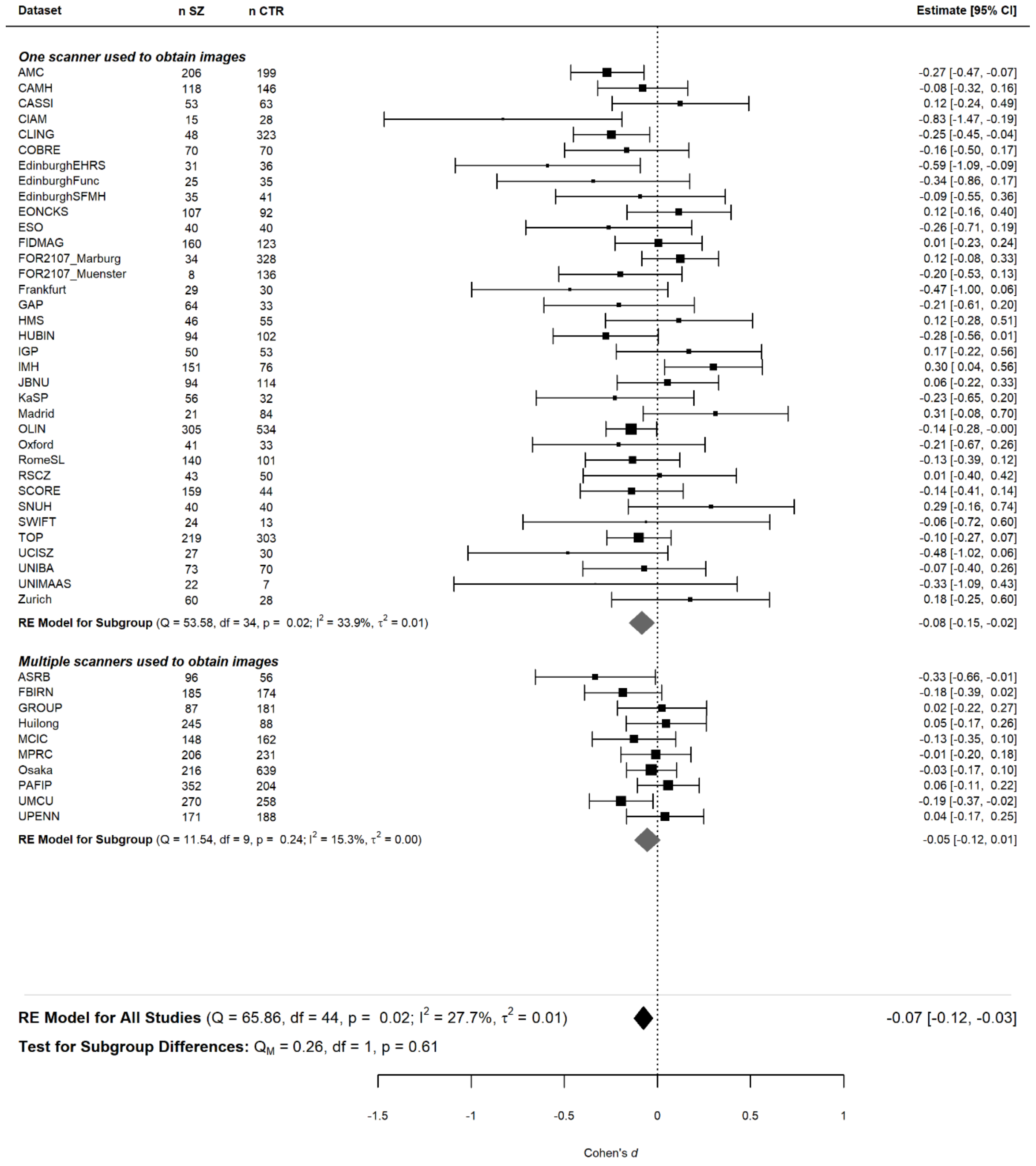

**Fig. S14. Forest plot where datasets are grouped by the use of a single scanner versus multiple scanners, for random effects meta-analysis of middle temporal gyrus thickness asymmetry differences between schizophrenia individuals and unaffected controls.** Shown are the per-dataset effect sizes, including confidence intervals, and the numbers of affected individuals (n SZ) and unaffected controls (n CTR). Cohen's *d* dot sizes represent relative dataset sizes. Meta-analyzed effect sizes are shown at a group level (gray diamonds) and across all studies (black diamonds), as well as between-dataset heterogeneity statistics (Cochran's Q test statistics). Single scanner versus multiple scanner group differences were tested using an omnibus test for heterogeneity. Groups are ordered from largest to smallest.

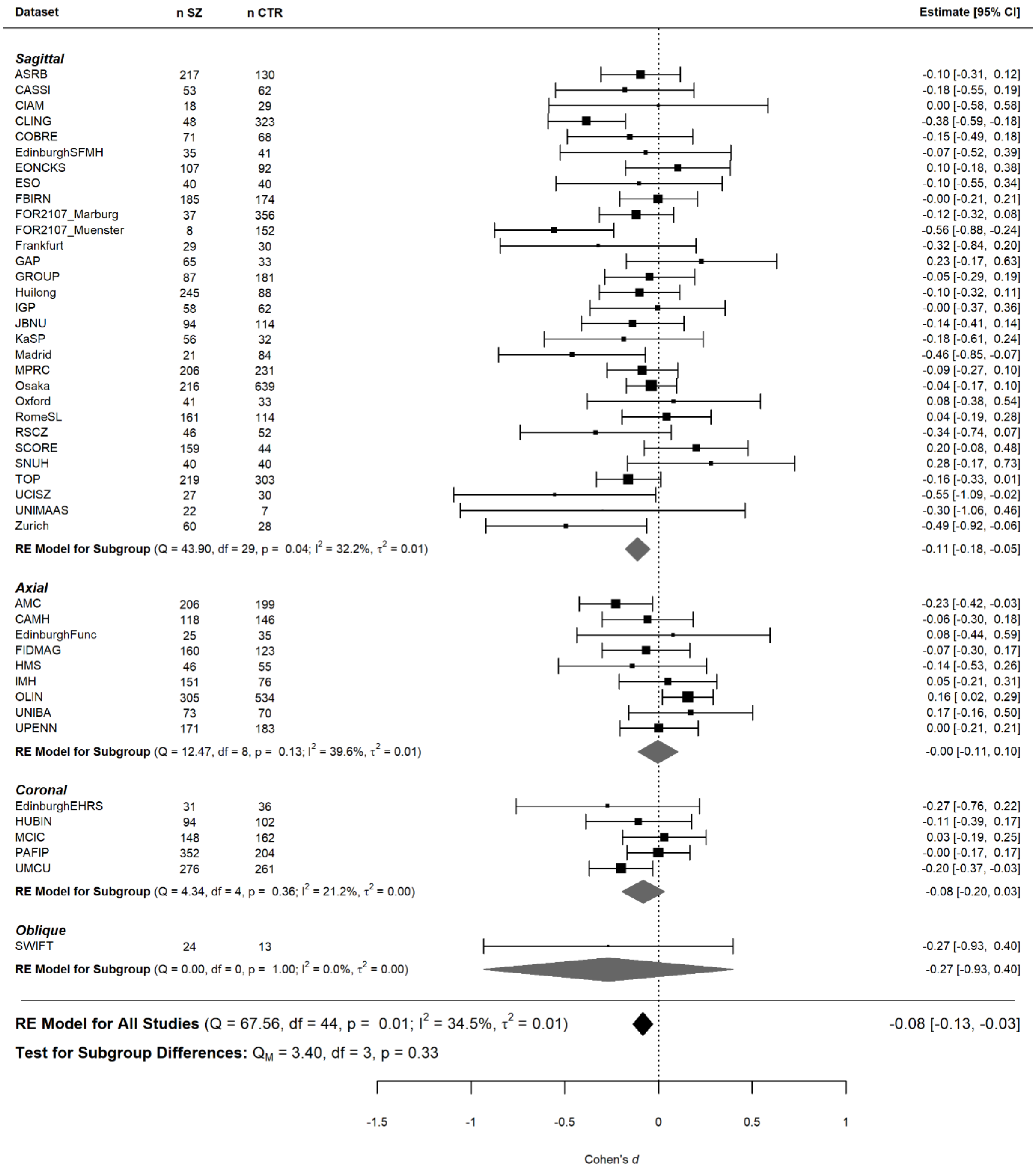

**Fig. S15. Forest plot grouped by image slice orientation, for random effects meta-analysis of rostral anterior cingulate thickness asymmetry differences between schizophrenia individuals and unaffected controls.** Shown are the per-dataset effect sizes, including confidence intervals, and the numbers of affected individuals (n SZ) and unaffected controls (n CTR). Cohen's *d* dot sizes represent relative dataset sizes. Meta-analyzed effect sizes are shown at a group level (gray diamonds) and across all studies (black diamonds), as well as between-dataset heterogeneity statistics (Cochran's Q test statistics). Image slice orientation group differences were tested using an omnibus test for heterogeneity. Groups are ordered from largest to smallest.

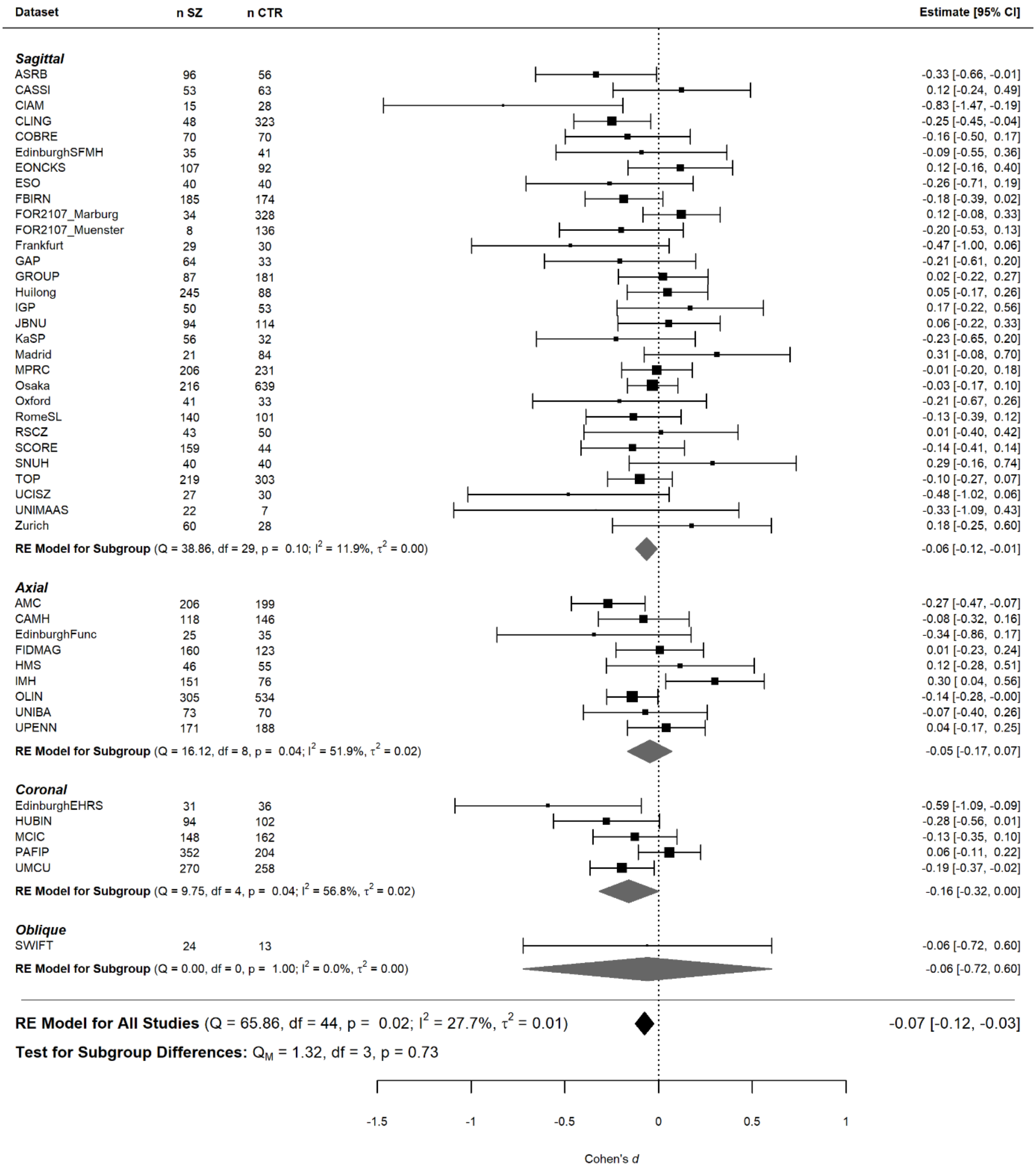

**Fig. S16. Forest plot grouped by image slice orientation, for random effects meta-analysis of middle temporal gyrus thickness asymmetry differences between schizophrenia individuals and unaffected controls.** Shown are the per-dataset effect sizes, including confidence intervals, and the numbers of affected individuals (n SZ) and unaffected controls (n CTR). Cohen's *d* dot sizes represent relative dataset sizes. Meta-analyzed effect sizes are shown at a group level (gray diamonds) and across all studies (black diamonds), as well as between-dataset heterogeneity statistics (Cochran's Q test statistics). Image slice orientation group differences were tested using an omnibus test for heterogeneity. Groups are ordered from largest to smallest.

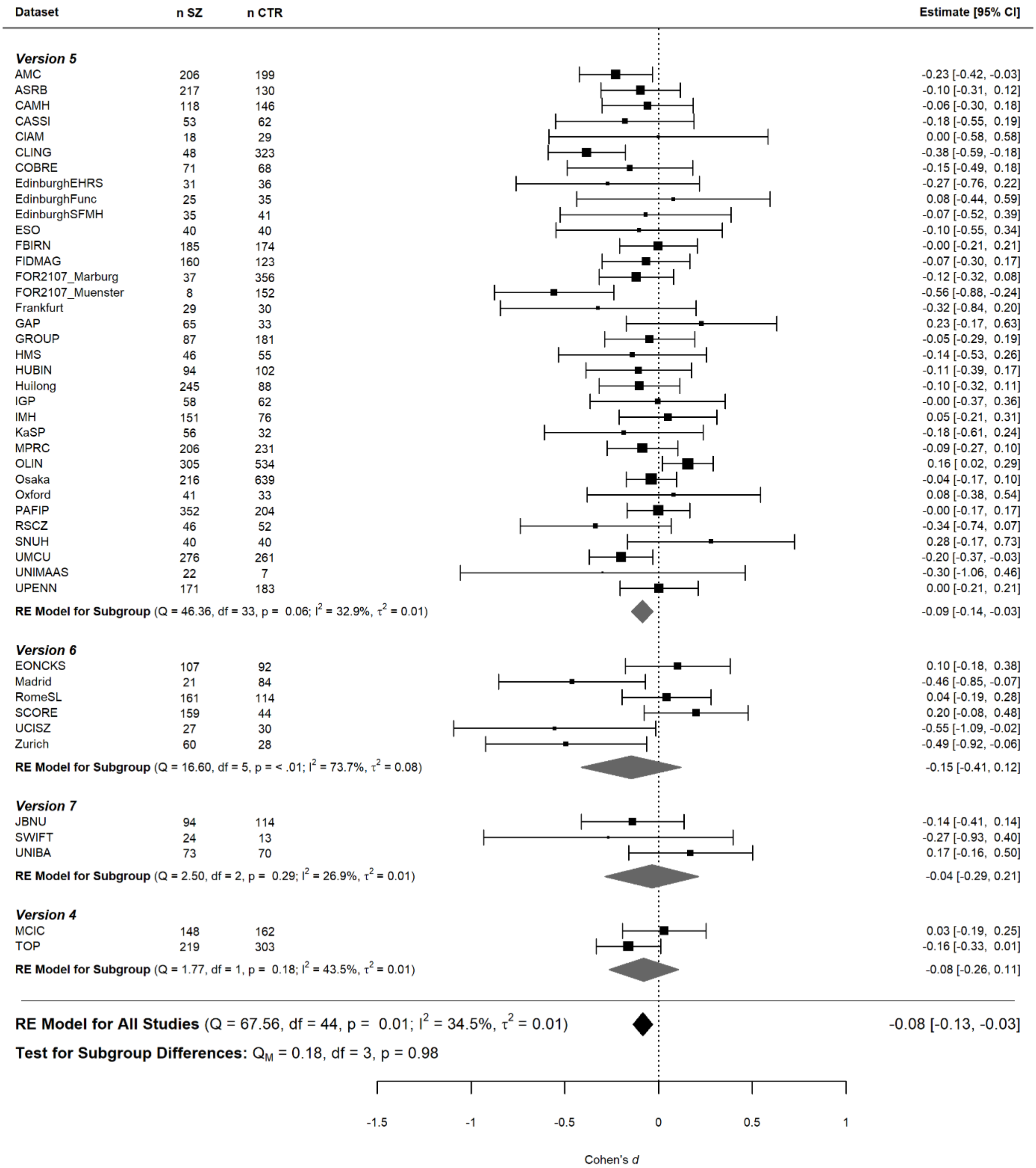

**Fig. S17. Forest plot grouped by Freesurfer version, for random effects meta-analysis of rostral anterior cingulate thickness asymmetry differences between schizophrenia individuals and unaffected controls.** Shown are the per-dataset effect sizes, including confidence intervals, and the numbers of affected individuals (n SZ) and unaffected controls (n CTR). Cohen's *d* dot sizes represent relative dataset sizes. Meta-analyzed effect sizes are shown at a group level (gray diamonds) and across all studies (black diamonds), as well as between-dataset heterogeneity statistics (Cochran's Q test statistics). Freesurfer version group differences were tested using an omnibus test for heterogeneity. Groups are ordered from largest to smallest.

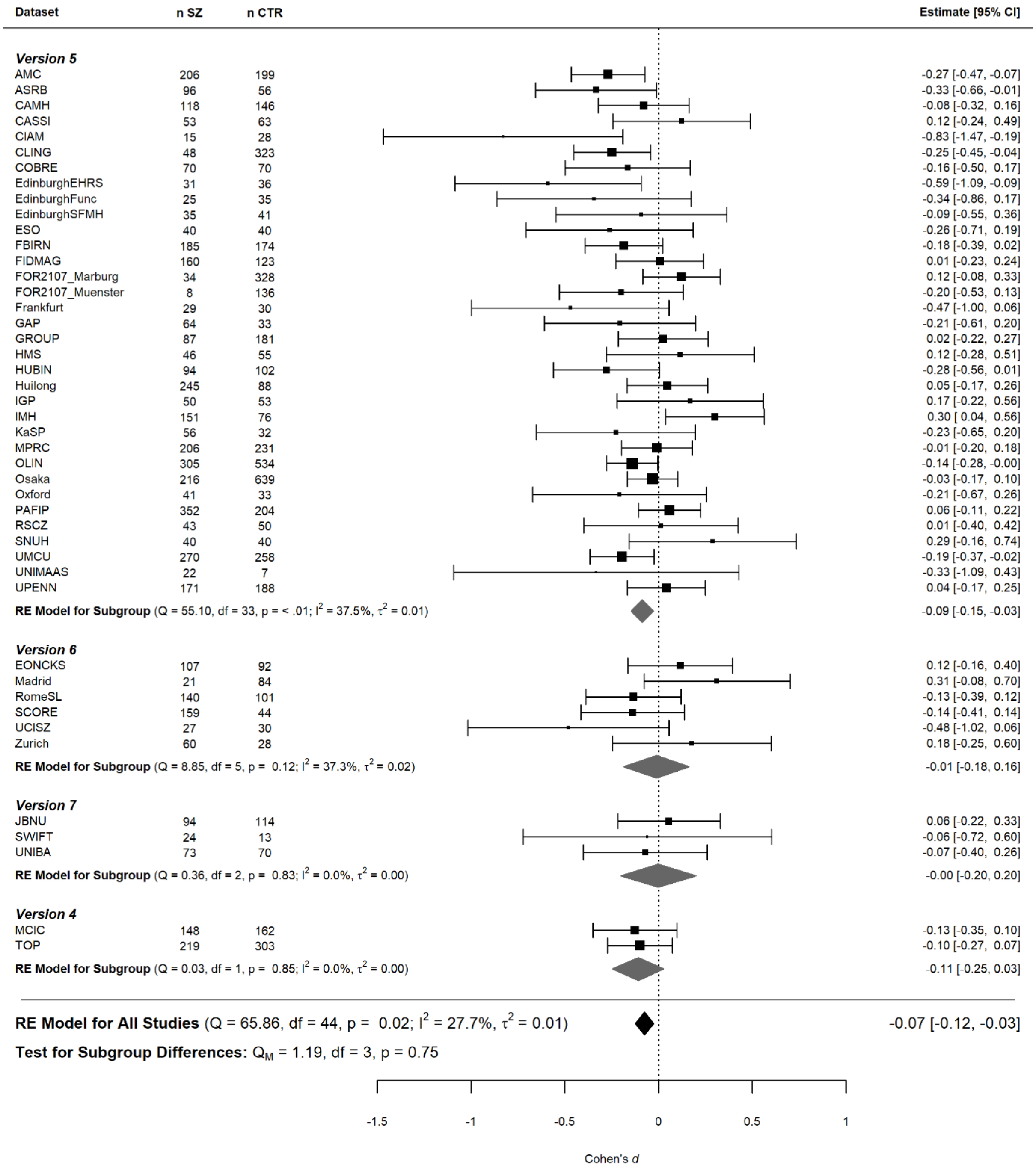

**Fig. S18. Forest plot grouped by Freesurfer version, for random effects meta-analysis of middle temporal gyrus thickness asymmetry differences between schizophrenia individuals and unaffected controls.** Shown are the per-dataset effect sizes, including confidence intervals, and the numbers of affected individuals (n SZ) and unaffected controls (n CTR). Cohen's *d* dot sizes represent relative dataset sizes. Meta-analyzed effect sizes are shown at a group level (gray diamonds) and across all studies (black diamonds), as well as between-dataset heterogeneity statistics (Cochran's Q test statistics). Freesurfer version group differences were tested using an omnibus test for heterogeneity. Groups are ordered from largest to smallest.

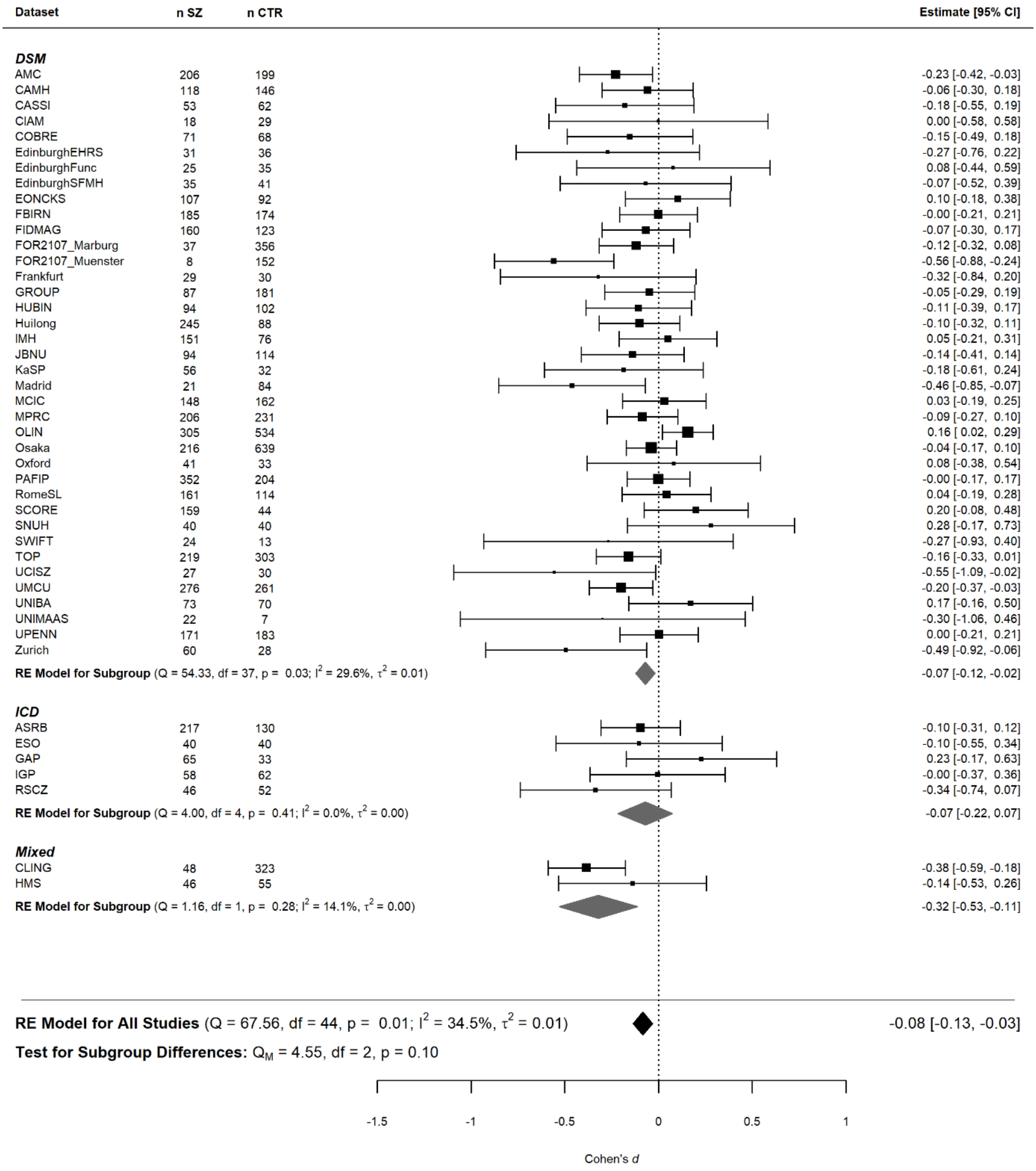

**Fig. S19. Forest plot grouped by diagnostic tool, for random effects meta-analysis of rostral anterior cingulate thickness asymmetry differences between schizophrenia individuals and unaffected controls.** Shown are the per-dataset effect sizes, including confidence intervals, and the numbers of affected individuals (n SZ) and unaffected controls (n CTR). Cohen's *d* dot sizes represent relative dataset sizes. Meta-analyzed effect sizes are shown at a group level (gray diamonds) and across all studies (black diamonds), as well as between-dataset heterogeneity statistics (Cochran's Q test statistics). Diagnostic tool group differences were tested using an omnibus test for heterogeneity. Groups are ordered from largest to smallest.

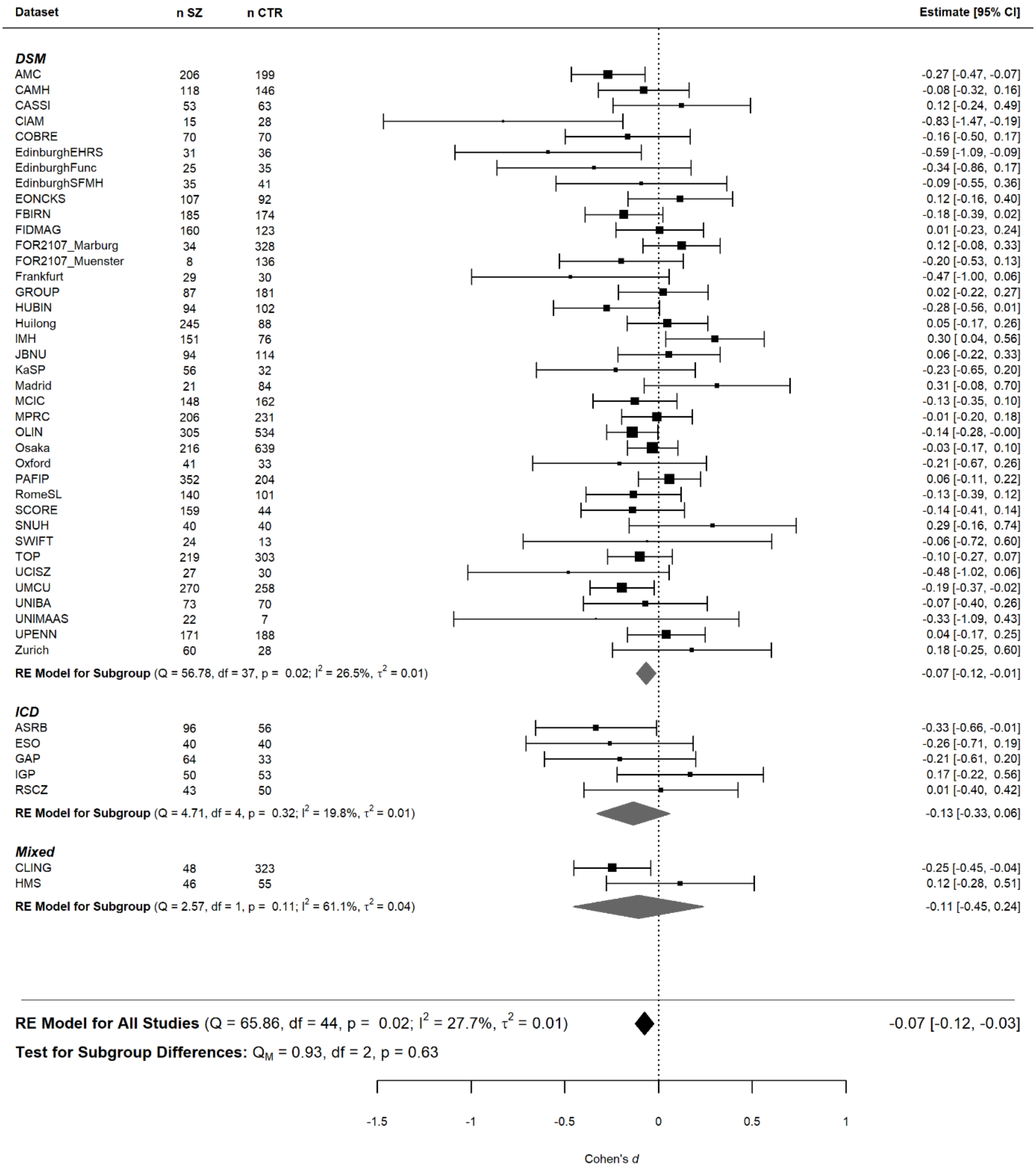

**Fig. S20. Forest plot grouped by diagnostic tool, for random effects meta-analysis of middle temporal gyrus thickness asymmetry differences between schizophrenia individuals and unaffected controls.** Shown are the per-dataset effect sizes, including confidence intervals, and the numbers of affected individuals (n SZ) and unaffected controls (n CTR). Cohen's *d* dot sizes represent relative dataset sizes. Meta-analyzed effect sizes are shown at a group level (gray diamonds) and across all studies (black diamonds), as well as between-dataset heterogeneity statistics (Cochran's Q test statistics). Diagnostic tool group differences were tested using an omnibus test for heterogeneity. Groups are ordered from largest to smallest.

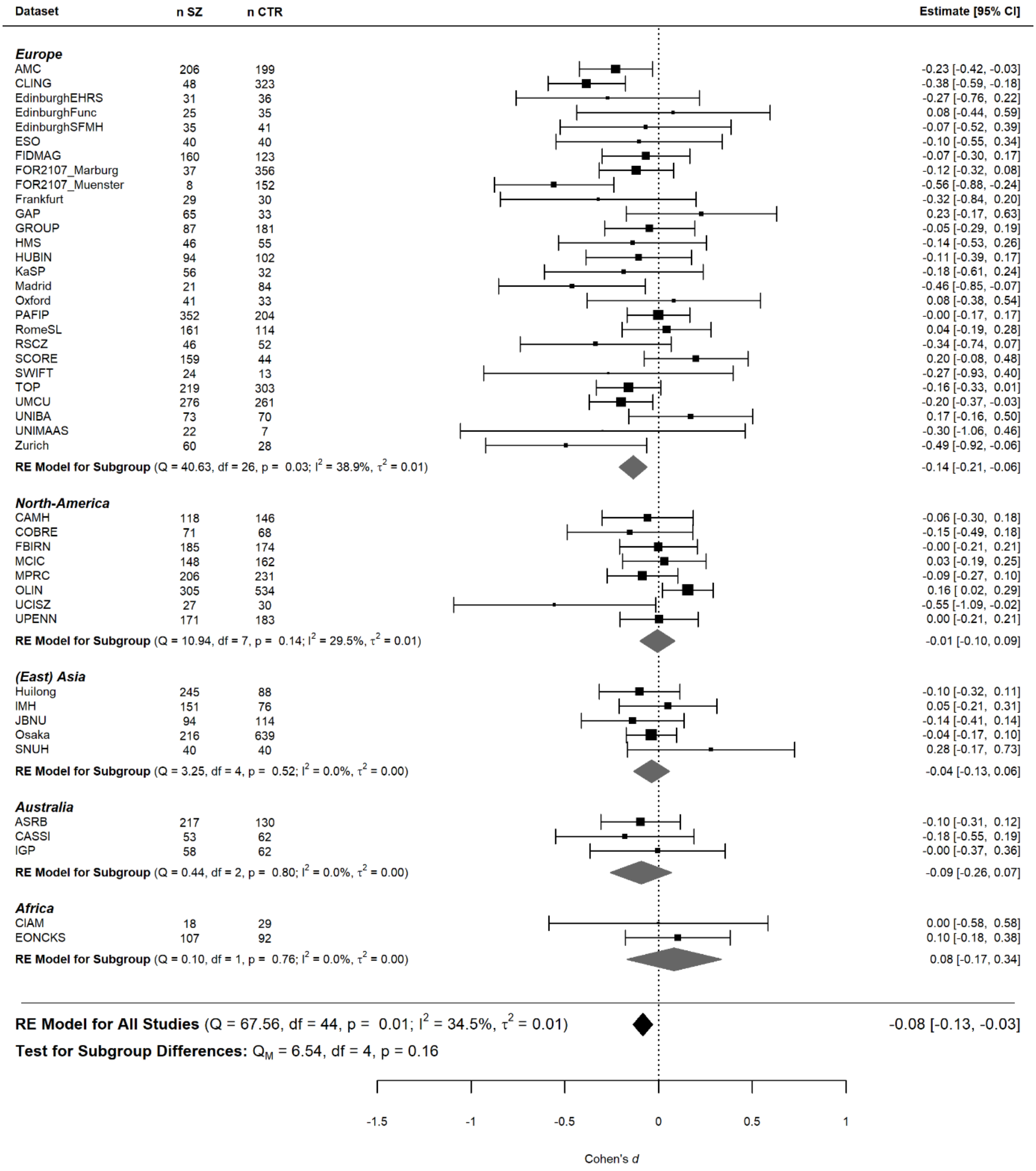

**Fig. S21. Forest plot grouped by geographic origin, for random effects meta-analysis of rostral anterior cingulate thickness asymmetry differences between schizophrenia individuals and unaffected controls.** Shown are the per-dataset effect sizes, including confidence intervals, and the numbers of affected individuals (n SZ) and unaffected controls (n CTR). Cohen's *d* dot sizes represent relative dataset sizes. Meta-analyzed effect sizes are shown at a group level (gray diamonds) and across all studies (black diamonds), as well as between-dataset heterogeneity statistics (Cochran's Q test statistics). Image slice orientation group differences were tested using an omnibus test for heterogeneity. Geographic origin groups are ordered from largest to smallest.

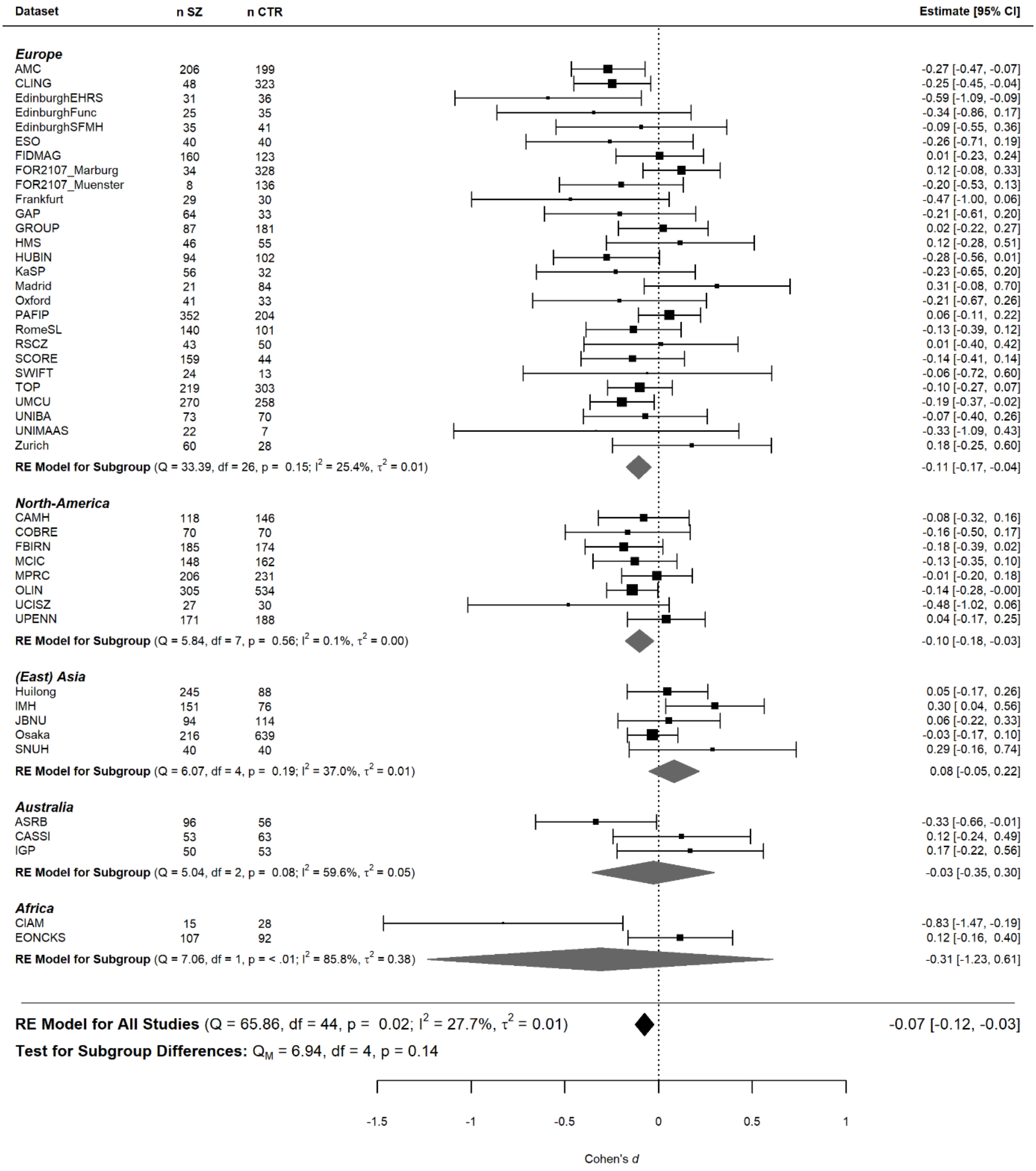

**Fig. S22. Forest plot grouped by geographic origin, for random effects meta-analysis of middle temporal gyrus thickness asymmetry differences between schizophrenia individuals and unaffected controls.** Shown are the per-dataset effect sizes, including confidence intervals, and the numbers of affected individuals (n SZ) and unaffected controls (n CTR). Cohen's *d* dot sizes represent relative dataset sizes. Meta-analyzed effect sizes are shown at a group level (gray diamonds) and across all studies (black diamonds), as well as between-dataset heterogeneity statistics (Cochran's Q test statistics). Geographic origin group differences were tested using an omnibus test for heterogeneity. Groups are ordered from largest to smallest.

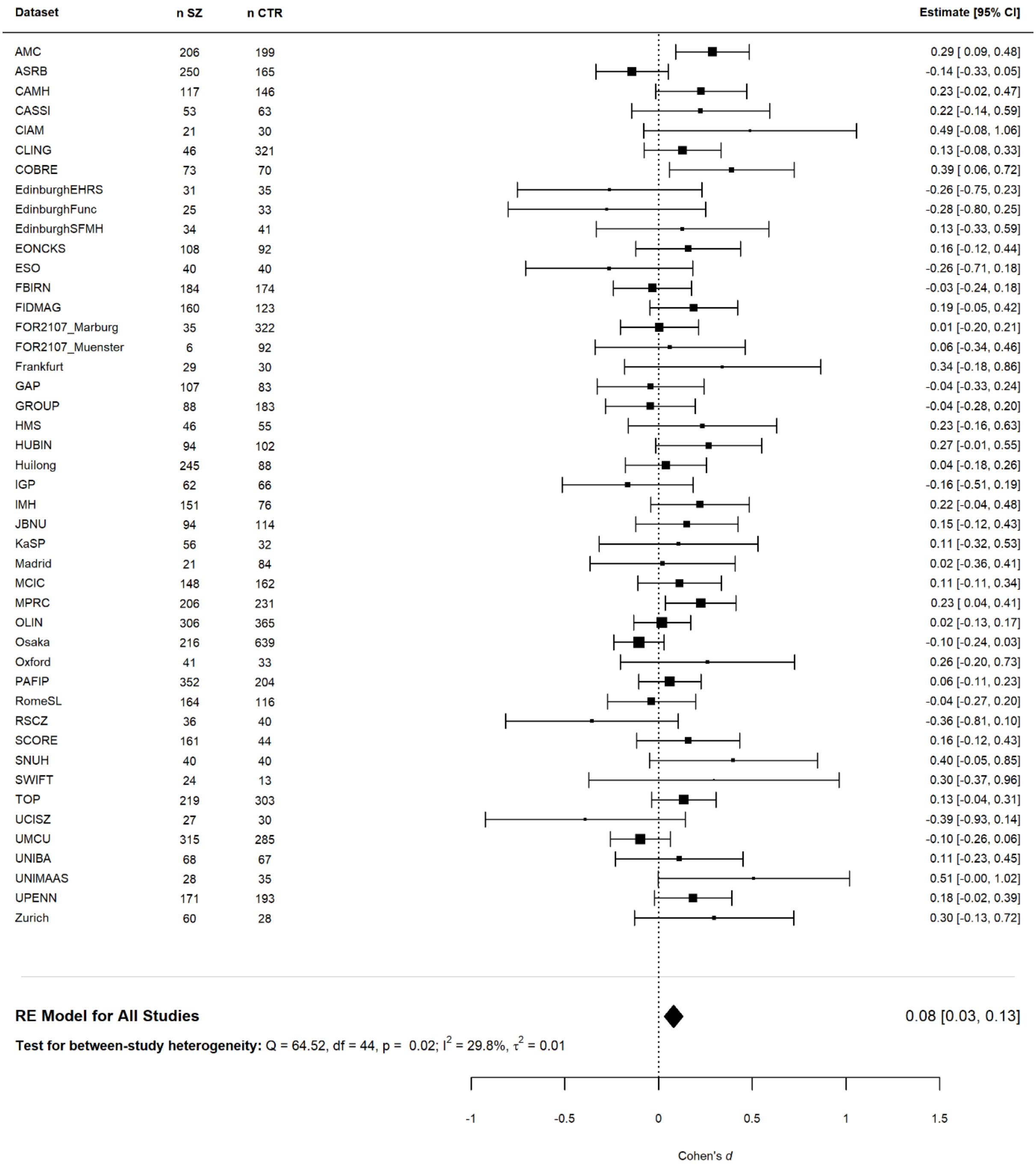

**Fig. S23. Forest plot for random effects meta-analysis of pallidum volume asymmetry differences between schizophrenia individuals and unaffected controls, with age interaction.** Shown are the per-dataset effect sizes, including confidence intervals, of affected individuals (n SZ) and unaffected controls (n CTR). Cohen's *d* dot sizes represent relative dataset sizes. The meta-analyzed effect sizes across all studies is shown (black diamond), as well as between-dataset heterogeneity statistics (Cochran's Q test statistics).

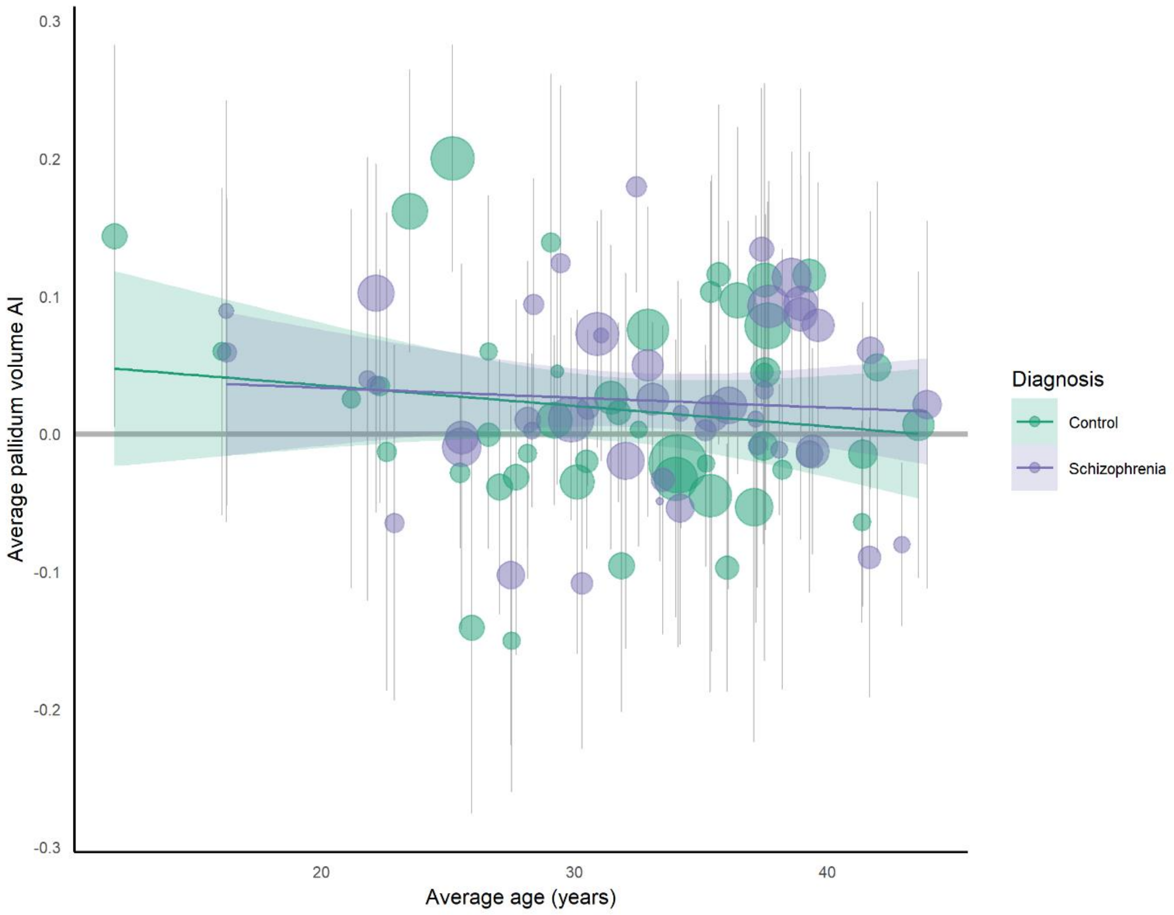

**Fig. S24. Average pallidum volume asymmetry against average age per dataset.** The average pallidum volume asymmetry index is plotted separately per dataset and for controls (green) and individuals with schizophrenia (purple). Point size indicates the relative sample size of each group per dataset, error bars show standard deviations of the average AI. The regression lines and their shaded confidence intervals show the linear relationships between pallidum volume AIs and age separately in cases and controls – revealing a possible, small diagnosis-by-age interaction effect.

### Acknowledgments per dataset

Acknowledgments per dataset are as follows:

**AMC:** The AMC study was supported by grants from ZonMW (grant numbers: 3160007, 91676084, 31160003, 31180002, 31000056, 2812412, 100001002, 100002034), NWO (grant numbers: 90461193, 40007080, 48004004, 40003330), and grants from the Amsterdam Brain Imaging Platform, Neuroscience Campus Amsterdam and the Dutch Brain foundation. The processing with Freesurfer was performed on the Dutch e-Science Grid through BiG Grid project and COMMIT project "e-Biobanking with imaging for healthcare", which are funded by the Netherlands Organization for Scientific Research (NWO).

**ASRB:** The Australian Schizophrenia Research Bank (ASRB), was supported by the National Health and Medical Research Council of Australia (NHMRC) (Enabling Grant, ID 386500), the Pratt Foundation, Ramsay Health Care, the Viertel Charitable Foundation and the Schizophrenia Research Institute. Chief Investigators for ASRB were Carr, V., Schall, U., Scott, R., Jablensky, A., Mowry, B., Michie, P., Catts, S., Henskens, F., Pantelis, C. We thank Loughland, C., the ASRB Manager, and acknowledge the help of Jason Bridge for ASRB database queries.

**CAMH:** The CAMH datasets were collected and shared with support from the CAMH Foundation and the Canadian Institutes of Health Research.

**CASSI:** The CASSI data set was supported by the University of New South Wales School of Psychiatry, the National Health and Medical Research Council (NHMRC) of Australia Project Grant no. 568807, Neuroscience Research Australia, the Schizophrenia Research Institute utilizing infrastructure funding from NSW Ministry of Health and the Macquarie Group Foundation and the Australian Schizophrenia Research Bank, which was supported by the NHMRC of Australia, the Pratt Foundation, Ramsay Health Care and the Viertel Charitable Foundation.

**CIAM:** The CIAM group (PI: FMH) was supported by the University Research Committee, University of CapeTown; South African National Research Foundation (NRF); South African Medical Research Council (SA MRC)

**CLING:** Sample data collection of the CliNG/KFO sample was partially supported by a grant of the Deutsche Forschungsgemeinschaft (DFG) to OG (grant number GR1950/5-1).

**COBRE:** The COBRE dataset and investigators were supported by NIH grants R01EB006841 & P20GM103472, as well as NSF grant 1539067. JAT (senior author) and VDC are supported by 5R01MH094524. JMS is supported by R01 AA021771 and P50 AA022534.

**EdinburghEHRS:** Funded by the Medical Research Council (MRC) [Grant Numbers: G9226254 & G9825423] and the Dr. Mortimer and Theresa Sackler Foundation.

**EdinburghFunc:** Functional Psychosis – Funded by the MRC Clinical Training Fellowship (Ref G84/5699) and Health Foundation Clinician Scientist Fellowship (Ref: 2268/4295).

**EdinburghSFMH:** Funded by an award from the Translational Medicine Research Collaboration (NS_EU_166) Scottish Enterprise and Pfizer and the Dr Mortimer and Theresa Sackler Foundation. The University of Edinburgh is a charitable body, registered in Scotland, with registration number SC005336. Is e buidheann carthannais a th’ ann an Oilthigh Dhùn Èideann, clàraichte an Alba, àireamh clàraidh SC005336.

**EONCKS:** This study was supported by the Medical Research Council of South Africa and the New Partnership for Africa's Development (NEPAD) initiative through the Department of Science and Technology of South Africa (Grant number #65174).

**ESO:** The ESO study was supported by the Ministry of Health of the Czech Republic, grant nr. NU20-04-00393.

**FBIRN**: The FBIRN study was supported by the National Center for Research Resources at the National Institutes of Health (NIH 1 U24 RR021992 (Function Biomedical Informatics Research Network) and NIH 1 U24 RR025736-01 (Biomedical Informatics Research Network Coordinating Center; http://www.birncommunity.org). FBIRN data was processed by the UCI High Performance Computing cluster supported by Joseph Farran, Harry Mangalam, and Adam Brenner and the National Center for Research Resources and the National Center for Advancing Translational Sciences, National Institutes of Health, through Grant UL1 TR000153. Adam Brenner was also supported by NIH grants 5R01 MH61603, and 2R01MH058251; Judith M. Ford by NIMH (R01 MH-58262). FBIRN thanks Mrs. Liv McMillan for overall study coordination.

**FIDMAG:** No relevant information to report.

**FOR2107 Marburg:** This work was funded by the German Research Foundation (DFG), Tilo Kircher (speaker FOR2107; DFG grant numbers KI 588/14-1, KI 588/14-2), Axel Krug (KR 3822/5-1, KR 3822/7-2), Igor Nenadić (NE 2254/1-2, NE2254/3-1, NE2254/4-1), Carsten Konrad (KO 4291/3-1), and Andreas Jansen (Grant No. FOR 2107, JA, 1890/7-1, 1890/7-2).

**FOR2107 Muenster:** The FOR2107 Muenster study was funded by the German Research Foundation (DFG, grant FOR2107 DA1151/5-1 and DA1151/5-2 to UD; SFB-TRR58, Projects C09 and Z02 to UD) and the Interdisciplinary Center for Clinical Research (IZKF) of the medical faculty of Münster (grant Dan3/012/17 to UD).

**Frankfurt:** MRI was performed at the Frankfurt Brain Imaging Centre, supported by the German Research Council (DFG) and the German Ministry for Education and Research (BMBF; Brain Imaging Center Frankfurt/Main, DLR 01GO0203).

**GAP:** The GAP dataset represents independent research funded by the NIHR Biomedical Research Centre at South London and Maudsley NHS Foundation Trust and King’s College London. The views expressed are those of the author(s) and not necessarily those of the NHS, the NIHR or the Department of Health.

**GIPSI:** This project was supported by “PRISMA U.T" Colciencias Invitación 990 del 3 de Agosto de 2017, Código 111577757629, Contrato 781 de 2017.

**GROUP:** We thank Truda Driesen and Inge Crolla for their coordinating roles in the data collection, as well as the G.R.O.U.P. investigators: René S. Kahn, Don H. Linszen, Jim van Os, Durk Wiersma; Richard Bruggeman, Wiepke Cahn, Lieuwe de Haan, Lydia Krabbendam, Inez Myin-Germeys. The infrastructure for the GROUP study is funded through the Geestkracht programme of the Dutch Health Research Council (Zon-Mw, grant number 10-000-1001), and matching funds from participating pharmaceutical companies (Lundbeck, AstraZeneca, Eli Lilly, Janssen Cilag) and universities and mental health care organizations (Amsterdam: Academic Psychiatric Centre of the Academic Medical Center and the mental health institutions: GGZ Ingeest, Arkin, Dijk en Duin, GGZ Rivierduinen, Erasmus Medical Centre, GGZ Noord Holland Noord. Groningen: University Medical Center Groningen and the mental health institutions: Lentis, GGZ Friesland, GGZ Drenthe, Dimence, Mediant, GGNet Warnsveld, Yulius Dordrecht and Parnassia psycho-medical center The Hague. Maastricht: Maastricht University Medical Centre and the mental health institutions: GGzE, GGZ Breburg, GGZ Oost-Brabant, Vincent van Gogh voor Geestelijke Gezondheid, Mondriaan, Virenze riagg, Zuyderland GGZ, MET ggz, Universitair Centrum Sint-Jozef Kortenberg, CAPRI University of Antwerp, PC Ziekeren Sint-Truiden, PZ Sancta Maria Sint-Truiden, GGZ Overpelt, OPZ Rekem. Utrecht: University Medical Center Utrecht and the mental health institutions Altrecht, GGZ Centraal and Delta).

**HMS:** Sample data collection of the HMS sample was supported by a grant of the Competence Network Schizophrenia to OG.

**HUBIN:** The HUBIN study was supported by the Swedish Research Council (K2015-62X-15077-12-3), 2017-00949, the regional agreement on medical training and clinical research between Stockholm County Council and the Karolinska Institutet, the Knut and Alice Wallenberg Foundation.

**Huilong:** This study was funded by the National Natural Science Foundation of China (81761128021; 31671145; 81401115; 81401133), Beijing Municipal Science & Technology Commission grant (Z141107002514016) and Beijing Natural Science Foundation (7162087, Beijing Municipal

Administration of Hospitals Clinical medicine Development of special funding (XMLX201609; zylx201409).

**IGP:** The Imaging Genetics in Psychosis (IGP) study was funded by Project Grants from the Australian National Health and Medical Research Council (NHMRC; APP630471 and APP1081603), and the Macquarie University's ARC Centre of Excellence in Cognition and its Disorders (CE110001021). This project used participants from the Australian Schizophrenia Research Bank (ASRB), funded by the NHMRC Enabling Grant (386500 held by V. Carr, U. Schall, R. Scott, A. Jablensky, B. Mowry, P. Michie, S. Catts, F. Henskens and C. Pantelis; Chief Investigators), and the Pratt Foundation, Ramsay Health Care, the Viertel Charitable Foundation, as well the Schizophrenia Research Institute, using an infrastructure grant from the NSW Ministry of Health.

**IMH:** This work was supported by research grants from the National Healthcare Group, Singapore (SIG/05004; SIG/05028), and the Singapore Bioimaging Consortium (RP C009/2006) research grants awarded to KS.

**JBNU:** Y-CC was supported by a grant of the Korean Mental Health Technology R&D Project, Ministry of Health & Welfare, Republic of Korea (HL19C0015) and a grant of the Korea Health Technology R&D Project through the Korea Health Industry Development Institute (KHIDI), funded by the Ministry of Health & Welfare, Republic of Korea (HI18C2383).

**KaSP:** KaSP was supported by the Swedish Research Council (K2015-62X-15077-12-3), and by by grants from the Swedish Medical Research Council (SE: 2009-7053; 2013-2838; SC: 523- 2014-3467), the Swedish Brain Foundation, Åhlén-siftelsen, Svenska Läkaresällskapet, Petrus och Augusta Hedlunds Stiftelse, Torsten Söderbergs Stiftelse, the AstraZeneca-Karolinska Institutet Joint Research Program in Translational Science, Söderbergs Königska Stiftelse, Professor Bror Gadelius Minne, Knut och Alice Wallenbergs stiftelse, the Swedish Federal Government under the LUA/ALF agreement (CMS, SCe), Centre for Psychiatry Research, KID-funding from the Karolinska Institutet.

**Madrid:** This work was supported by the Spanish Ministry of Science and Innovation. Instituto de Salud Carlos III (SAM16PE07CP1, PI16/02012, PI19/024), co-financed by ERDF Funds from the European Commission, "A way of making Europe", CIBERSAM. Madrid Regional Government (B2017/BMD-3740 AGES-CM-2), European Union Structural Funds. European Union Seventh Framework Program under grant agreements FP7-4-HEALTH-2009-2.2.1-2-241909 (Project EU-GEI), FP7- HEALTH-2013-2.2.1-2-603196 (Project PSYSCAN) and FP7- HEALTH-2013-2.2.1-2-602478 (Project METSY); and European Union H2020 Program under the Innovative Medicines Initiative 2 Joint Undertaking (grant agreement No 115916, Project PRISM, and grant agreement No 777394, Project AIMS-2-TRIALS), Fundación Familia Alonso and Fundación Alicia Koplowitz.

**MCIC:** The MCIC study was supported by the National Institutes of Health (NIH/NCRR P41RR14075 and R01EB005846 to Vince D. Calhoun), the Department of Energy (DE-FG02-99ER62764), the Mind Research Network, the Morphometry BIRN (1U24, RR021382A), the Function BIRN (U24RR021992-01, NIH.NCRR MO1 RR025758-01, NIMH 1RC1MH089257 to Vince D. Calhoun), the Deutsche Forschungsgemeinschaft (research fellowship to Stefan Ehrlich), and a NARSAD Young Investigator Award (to Stefan Ehrlich).

**MPRC:** Support was received from NIH grants U01MH108148, 2R01EB015611, R01MH112180, R01DA027680, R01MH085646, P50MH103222 and T32MH067533, a State of Maryland contract (M00B6400091) and NSF grant (1620457).

**OLIN:** The Olin study was supported by NIH grants R01MH106324 and R01MH077945.

**Osaka:** The Osaka study was partially supported by AMED under Grant Number JP21dm0307002, JP21dm0207069, JP21dk0307103, JP21uk1024002 and JP21wm0425012, JSPS KAKENHI under Grant Number JP20H03611 and JP20K06920, and Intramural Research Grant (3-1) for Neurological and Psychiatric Disorders of NCNP. The computations were performed using Research Center for Computational Science, Okazaki, Japan.

**Oxford:** We would like to thank the participants and their families, referring psychiatrists and the Donnington Health Centre, Oxford. This study is supported by the MRC, OHSRC, UK EPSRC, BBSRC and Wellcome Trust.

**PAFIP:** The authors wish to thank all PAFIP research teams and all patients and family members who participated in the study. PAFIP has been funded by Instituto de Salud Carlos III through the projects PI14/00639, PI14/00918 and PI17/01056 (Co-funded by European Regional Development Fund / European Social Fund "Investing in your future") and Health Research Institute Marques de Valdecilla NTC0235832 and NCT02534363. The PAFIP study was further supported by Instituto de Salud Carlos III, FIS 00/3095, 01/3129, PI020499, PI060507, PI10/00183, the SENY Fundació Research Grant CI 2005-0308007, and the Fundación Marqués de Valdecilla API07/011. The PAFIP study was also funded by Instituto de Salud Carlos III, MINECOSAF2013-46292-R, PSYSCAN (Exp.: HEALTH.2013.2.2.1-2_Grant agreement no. 603196). We want to particularly acknowledge the patients and the BioBankValdecilla (PT13/0010/0024) integrated in the Spanish National Biobanks Network for its collaboration. We thank IDIVAL Neuroimaging Unit for its help in the technical execution of this work.

**RomeSL:** No relevant information to report.

**RSCZ:** RSCZ was supported in part by the RFBR grant 20-013-00748.

**SCORE:** This study was supported in part by grant 3232BO_119382 from the Swiss National Science Foundation. We thank the FePsy (Frueherkennung von Psychosen; early detection of psychosis) Study Group from the University of Basel, Department of Psychiatry, Switzerland, for the recruitment of the study participants. The FePsy Study was supported in part by grant No. SNF 3200-057216/1, ext./2, ext./3.

**SNUH:** This work was supported by the Basic Science Research Program through the National Research Foundation of Korea (NRF) and the Korea Brain Research Institute (KBRI) basic research program through the KBRI, funded by the Ministry of Science, ICT & Future Planning (grant nos. 2019R1C1C1002457, 2020M3E5D9079910 and 21-BR-03–01).

**SWIFT:** This research was supported in part by the Swiss National Science Foundation (grant no. 320030_146789).

**TOP:** The TOP study was supported by the Research Council of Norway (#160181, 190311, 223273, 213837, 249711), the South-East Norway Health Authority (2014114, 2014097, 2017- 112), and the Kristian Gerhard Jebsen Stiftelsen (SKGJ-MED-008) and the European Community's Seventh Framework Programme (FP7/2007–2013), grant agreement no. 602450 (IMAGEMEND).

**UCISZ:** The UCISZ study was supported by the National Institutes of Mental Health grant number R21MH097196 to TGMvE. UCISZ data was processed by the UCI High Performance Computing cluster supported by Joseph Farran, Harry Mangalam, and Adam Brenner and the National Center for Research Resources and the National Center for Advancing Translational Sciences, National Institutes of Health, through Grant UL1 TR000153. Adam Brenner was also supported by NIH grants 5R01 MH61603, and 2R01MH058251; JF by NIMH (R01 MH-58262).

**UMCU:** The UMCU study was supported by the Netherlands Organization for Health Research and Development Zon-Mw grants 90802123 and 91746370 (to Hilleke E. Hulshoff Pol) and 10-000-1001 (to René S. Kahn).

**UNIBA:** The UNIBA study was supported by grant funding from the Italian Ministry of Research (2017M7SZM8_004, PI Bertolino; 2017K2NEF4, PI Pergola).

**UNIMAAS:** This work was supported by the Dutch Organization for Health Research and Development (ZonMw 91112002) and by a personal grant to Thérèse van Amelsvoort (ZonMw-VIDI: 91712394). This clinical trial was registered in the Dutch clinical trial registry under ID: NTR5094 (http://www.trialregister.nl).

**UPENN:** No relevant information to report.

**Zurich:** The Zurich dataset was funded by the Swiss National Science Foundation.
